# Cardiometabolic drugs and kidney function – a drug target Mendelian randomisation study

**DOI:** 10.1101/2025.06.16.25329403

**Authors:** Aristomo Andries, Schyler Marie Bennett, Venexia Walker, Iris M. Heid, Anna Köttgen, Mathias Gorski, Artemis Briasouli, Jie Zheng, Kristian Hveem, Stein Ivar Hallan, Bjørn Olav Åsvold, Tom Gaunt, Ben Michael Brumpton, Humaira Rasheed

**Affiliations:** HUNT Center for Molecular and Clinical Epidemiology (MCE), Norwegian University of Science and Technology (NTNU); Department of Public Health and Nursing, NTNU; Medical Research Council Integrative Epidemiology Unit, University of Bristol; Department of Surgery, Perelman School of Medicine at the University of Pennsylvania; Department of Genetic Epidemiology, University of Regensburg, Regensburg, Germany; Institute of Genetic Epidemiology, Faculty of Medicine and Medical Center - University of Freiburg, Freiburg, Germany; Department of Epidemiology, Johns Hopkins Bloomberg School of Public Health, Baltimore, MD, USA; Department of Nephrology, St. Olav’s University Hospital; Department of Clinical and Molecular Medicine, NTNU; Department of Endocrinology, St. Olav’s University Hospital; Institute of Clinical Medicine, University of Oslo; Nordic RWE, Oslo, Norway

**Author notes:** Corresponding author: Aristomo Andries, Department of Public Health and Nursing, Faculty of Medicine and Health Sciences, NTNU Norwegian University of Science and Technology, P.O. Box 8905, MTFS, NO-7491, Trondheim, Norway.

**Keywords:** chronic kidney disease, cardiometabolic, mendelian randomization, drug target, genome-wide association study

## Abstract

**Background:** Chronic kidney disease (CKD) is a growing global public health concern. Using drug target Mendelian randomization (MR) approach, we assessed the effects of cardiometabolic drug classes on kidney function indices in the general population.

**Method:** We leveraged 93 single nucleotide polymorphisms (SNPs) as genetically proxies for targets of antihypertensives (Ca-channel blockers/CCB, beta-blockers/BB, ACE inhibitors/ACEI), lipid-lowering drugs (PCSK9 inhibitors/PCSK9I, statins, NPC1L1 inhibitors/NPC1L1I), and antidiabetics (metformin, SGLT2 inhibitors/SGLT2I, GLP1R agonist/GLP1RA) drugs. Genetic instruments reflected lowering of systolic blood pressure (SBP), LDL cholesterol, and glycated haemoglobin (HbA1c). We used summary statistics from genome-wide association study (GWAS) of log natural annual change of estimate glomerular filtration rate (eGFR) (n=334,339), cross-sectional creatinine- and cystatin-based eGFR, and blood urea nitrogen (BUN) (n=1,201,909) as kidney function outcome. Two-sample MR and sensitivity methods (simple mode, weighted mode, weighted median, and MR Egger) were applied.

**Results:** The annual change of eGFR was attenuated by genetically proxied PCSK9I (β=0.066 per mmol/L LDL lowering; 95%CI 0.008,0.125). Cross-sectional eGFR_crea_ was increased by CCBs (β=0.006 per 10 mmHg SBP lowering; 95%CI 0.001,0.011), SGLT2I (β=0.024-0.028), MC1 (β=0.022), and GDF15 (β=0.094–0.100) per 6.7 mmol/mol HbA1c lowering. Statins and NPC1L1I may reduce eGFR and SGLT2I may decrease annual eGFR change.

**Conclusion:** Our findings provided genetic evidence that specific cardiometabolic drug targets influence kidney function in the general population. These insights may guide personalize drug selection to preserve renal function.

## Background

Chronic kidney disease (CKD) affects approximately 10% of the global population and remains a significant non-communicable disease (NCD) burden [1, 2]. A recent study in developed nations, including Germany, Italy, and France, reported that 62-96% of people with stage 3 kidney disease were undiagnosed [3], suggesting that the true prevalence of CKD might be higher. CKD is projected to become the fifth leading cause of death globally by 2040, rising from the sixteenth position in 2016 [4].

CKD is characterised by a sustained loss of kidney function and structures over a period of more than three months [5–7]. CKD is staged by evaluating estimated glomerular filtration rate (eGFR) and the urine albumin-to-creatinine ratio [7–10]. Both serum creatinine and cystatin C are used to calculate eGFR [7]. As the earlier stages of CKD are asymptomatic, patients are often unaware that their kidney function is deteriorating [11]. This condition usually affects a patient’s health for years and often remains undetectable until later stages [12]. At this point, significant damage to the kidney has already occurred. These later stages of kidney disease contribute to increased treatment complexity, kidney failure, cost, and mortality [11]. Patients with end stage kidney disease (ESKD) require dialysis or kidney transplant which is available in high resource settings only [13].

Unlike in the past two decades where lack of strong evidence due to small clinical trials with unclear clinical benefit hindered novel drug discovery [14, 15], drug development for CKD has recently gain more interest. An antidiabetic drug sodium-glucose cotransporter-2 (SGLT2) inhibitor has shown promising results for use in CKD patients [16, 17], highlighting the success of repurposing strategy. Many CKD patients have comorbidities that may contribute to deteriorating kidney function, such as type 2 diabetes mellitus (T2D), hypertension, and dyslipidaemia [18]. Thus, treatment that targets both CKD and its comorbidities may be beneficial for the kidney. One drug repurposing strategy is to explore potential kidney benefits from drugs that are mainly used to treat CKD comorbidities. This strategy is attractive since it can optimize the usage of existing treatments and save money compared with new drug development [19]. Drug target Mendelian randomization (MR) can be used to improve the likelihood of selecting a useful drug [20]. If results from an MR study support the findings from randomised control trials (RCTs), this will strengthen our confidence that a drug effect is likely to be true.

One approach in drug target MR is to estimate the effect of genetic variants (single nucleotide polymorphisms/SNPs) in or near drug target genes of an intermediate phenotype, such as systolic blood pressure (SBP), mimicking the effect of a drug [20, 21]. Subsequently, the effect of this drug on CKD is estimated. Similar with other uses of MR, drug target MR is based on the Mendelian principles that variants are randomly segregated and independently assorted at conception [22]. This process is unrelated to exposures after conception, making these variants less susceptible to confounders than the intermediate phenotypes they instrument. In drug target MR, subjects with higher and lower genetically-predicted levels of the intermediate phenotype are randomised in a way analogous to control and intervention groups in an RCT [23, 24].

An earlier study reported that two classes of antihypertensives, angiotensin-converting enzyme (ACE) inhibitor and calcium channel blocker (CCB), were associated with higher cross-sectional eGFR [25]. While this indicated a favourable outcome, the effect of these drugs on annual change in eGFR needs to be investigated. The first-line antidiabetic metformin has multiple markers. For example, a marker of metformin, AMP-activated protein kinase (AMPK), has been associated with positive renal outcomes [26]. As a frequently used medicine in CKD patients with cardiovascular disease, statins have shown potential in preventing eGFR decline [27]. Furthermore, there is a need to explore the potential effects of these cardiometabolic drugs on kidney function due to frequent coexistence of these comorbidities with CKD. Given the shared pathophysiology between CKD and cardiometabolic conditions, we hypothesize that cardiometabolic drugs may influence kidney function. Using a drug target two-sample MR approach, we estimated the effects of genetically proxied antihypertensive, antidiabetic, and lipid-lowering drugs on changes in eGFR, cross-sectional eGFR, and blood urea nitrogen (BUN) in the general population.

## Method

### Study design

This study applied a drug target MR using genetic summary statistics for the effects of antihypertensives, antidiabetic, and lipid-lowering drugs as exposures [28]. The outcomes were the annual change in creatinine-based eGFR [29], cross-sectional eGFR and BUN [30] (**Figure 1**).

**Figure 1.**
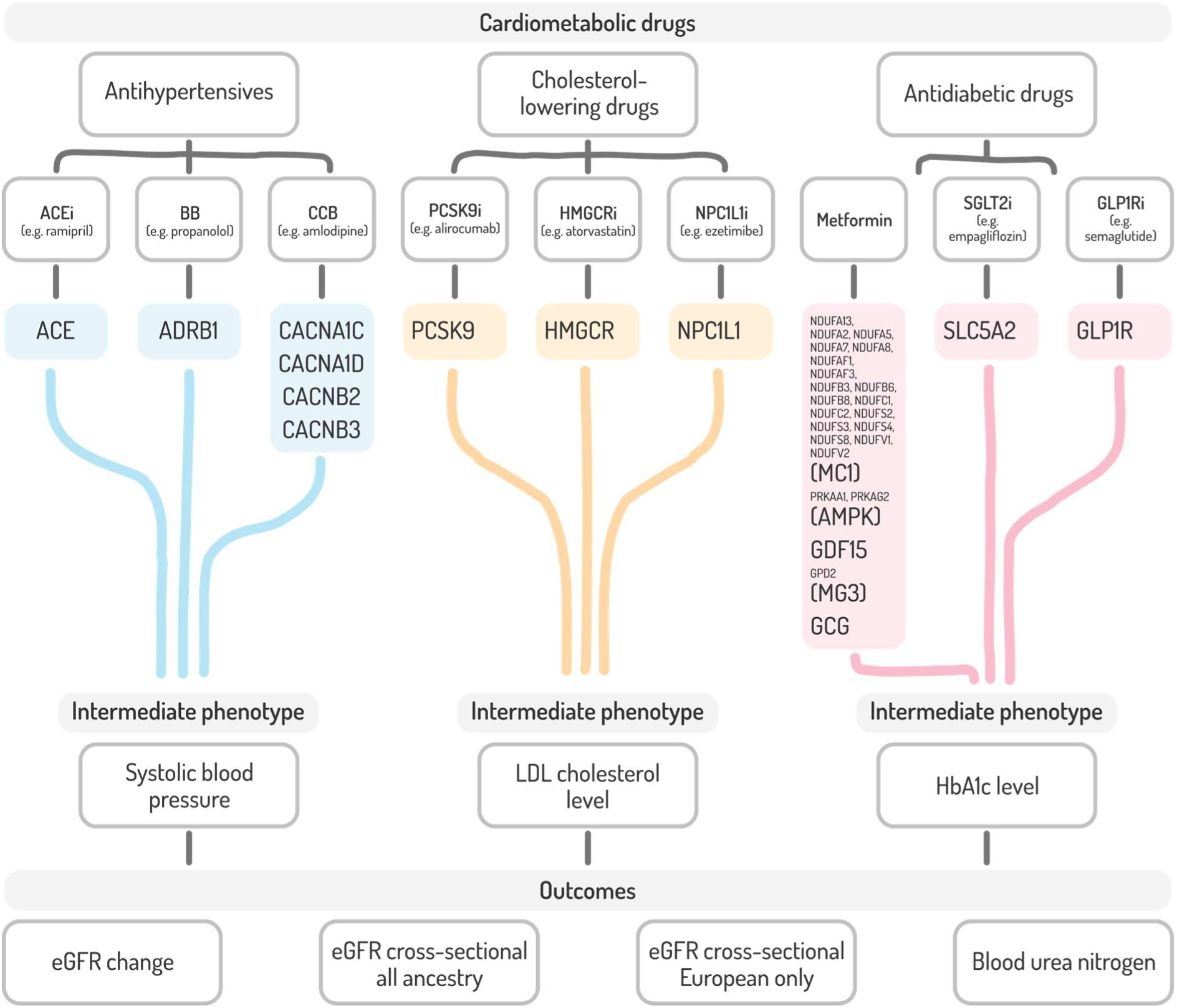
Study design illustrates the three cardiometabolic drugs selected for this study. For each drug class, drug target genes and intermediate phenotype-associated SNPs in or near the targets were retrieved from published literature. The drug target MR was applied for each cardiometabolic drug to intermediate phenotype to mimic the effect of each drug and estimate the effect on kidney function outcomes: eGFR change, eGFR cross-sectional (both all ancestry and European only), and blood urea nitrogen. ACE, angiotensin-converting enzyme; BB, beta-blocker; ADRB1, adrenoceptor beta 1; CCB, calcium channel blocker; CACNA1C, calcium voltage-gated channel subunit alpha1 C; CACNA1D, calcium voltage-gated channel subunit alpha1 D; CACNB2, calcium voltage-gated channel auxiliary subunit beta 2; CACNB3, calcium voltage-gated channel auxiliary subunit beta 3; PCSK9, proprotein convertase subtilisin/kexin type 9; HMGCR, 3-hydroxy-3-methyl-glutaryl-coenzyme A reductase; NPC1L1, Niemann-Pick C1-Like 1; SGLT2, sodium-glucose cotransporter-2; SLC5A2, solute carrier family 5, member 2; MC1, mitochondrial complex I; AMPK, AMP-activated protein kinase; GDF15, growth/differentiation factor 15; MG3, mitochondrial glycerol 3; GCG, glucagon; GLP1R, glucagon-like peptide 1 receptor

### Instrument selection

The SNPs for drug target genes of antihypertensive, antidiabetic, and lipid-lowering drugs and their effects for downstream cardiometabolic phenotypes were obtained from published literature [28, 31–37]. For antihypertensive drugs, three commonly used antihypertensive in outpatient setting, namely ACE inhibitor, beta-blocker (BB), and CCB, were explored. For lipid-lowering drug, we explored statin as a first line drug and two classes for advanced use: NPC1L1 inhibitor (e.g. ezetimibe) and PCSK9 inhibitor (e.g. evolo-/alirocumab). For antidiabetic, we explored a first-line drug metformin and more recent drugs sodium-glucose cotransporter-2 (SGLT2) inhibitor, which has undergone clinical studies in cardiac and kidney patients [16, 17], and glucagon-like peptide 1 receptor (GLP1R) agonist [38]. For each type of cardiometabolic drugs, SNPs were clumped using linkage disequilibrium threshold (LD) and p-value (**Table 1**).

**Table 1.** Exposure instruments selection criteria.

| Cardiometabolic drug type | Source of study | Source of summary statistics | Intermediate phenotypes | SNPs selection criteria | Genes and SNPs retrieved |
| --- | --- | --- | --- | --- | --- |
| Antihypertensives | Gill, et.al. (2019) [28] | GWAS meta-analysis of 757,601 individuals on SBP of European ancestry in UK Biobank. The GWAS was adjusted for body mass index [28, 39] | Systolic blood pressure (SBP) | Clumping was made using 1000G reference panel at LD $r^2 < 0.1$ and p-value $< 5 \times 10^{-8}$. | ACE inhibitor: ACE (1 SNP)<br>BB: ADRB1 (5 SNPs)<br>CCB: CACNA1C<br>CACNA1D, CACNB2, and CACNB3 (24 SNPs) |
| Lipid-lowering drugs | Liu, et.al. (2021) [31]<br>Zheng, et.al. (2020) [32] | GWAS summary statistics of 1.32 million individuals on LDL-C with European descent from Global Lipids Genetics Consortium (GLGC) [40]. | Low-density lipoprotein cholesterol (LDL-C) | Significant threshold at $< 5 \times 10^{-8}$. Clumping using 1000G reference panel with physical distance $\pm 250$ kb and LD $r^2 \leq 0.2$ (if $\geq 3$ SNPs were available) or $r^2 \leq 0.4$ (if $< 3$ SNPs were available) [31]. In Zheng, et.al. (2020) study, LD $r^2$ were $< 0.2$ for statin and PCSK9 inhibitor and $< 0.3$ for NPC1L1 inhibitor [32]. | PCSK9 inhibitor: PCSK9 (11 SNPs)<br>Statin: HMGCR (5 SNPs)<br>NPC1L1 inhibitor: NPC1L1 (6 SNPs) |
| Antidiabetic drugs | Zheng, et.al. (2022) [33]<br>Xu, et.al. (2022) [34]<br>Li, et.al. (2023) [35]<br>Daghlas, et.al. (2021) [36]<br>Yang, et.al. (2025) [37] | GWAS of 344,182 unrelated individuals on HbA1c of European ancestry without diabetes from UK Biobank [33-37]. | Glycated haemoglobin (HbA1c) | For SGLT2 inhibitor, SNPs were selected based on colocalization probability > 70% between SLC5A2 expression and HbA1c and LD $r^2 < 0.8$, using 1000G reference panel [34, 35]. For SNPs of metformin makers, colocalization probability was $\geq 80\%$ and LD $r^2 < 0.8$ [33]. For GLP1R agonist, LD $r^2 < 0.1$ [36] and 0.001 [37] were used. | SGLT2 inhibitor: SLC5A2 (6 SNPs)<br>Metformin: MC1 (25 SNPs), AMPK (3 SNPs), GDF15 (1 SNP), MG3 (1 SNP), and GCG (1 SNP)<br>GLP1R agonist: GLP1R (4 SNPs) |

ACE, angiotensin-converting enzyme; BB, beta-blocker; ADRB1, adrenoceptor beta 1; CCB, calcium channel blocker; CACNA1C, calcium voltage-gated channel subunit alpha1 C; CACNA1D, calcium voltage-gated channel subunit alpha1 D; CACNB2, calcium voltage-gated channel auxiliary subunit beta 2; CACNB3, calcium voltage-gated channel auxiliary subunit beta 3; PCSK9, proprotein convertase subtilisin/kexin type 9; HMGCR, 3-hydroxy-3-methyl-glutaryl-coenzyme A reductase; NPC1L1, Niemann-Pick C1-Like 1; SGLT2, sodium-glucose cotransporter-2; SLC5A2, solute carrier family 5, member 2; MC1, mitochondrial complex I; AMPK, AMP-activated protein kinase; GDF15, growth/differentiation factor 15; MG3, mitochondrial glycerol 3; GCG, glucagon; GLP1R, glucagon-like peptide 1 receptor; GWAS, genome-wide association study; LD, linkage disequilibrium; HbA1c, glycated haemoglobin; SNP, single nucleotide polymorphisms

After retrieving all SNPs from various summary statistics, we clumped the SNPs for each drug target gene using ± 250 kb clumping window and LD threshold r^2^ < 0.4 (**Supplementary table ST1**). This threshold has been used in the previous study to account for instrument availability [31]. We also performed a sensitivity analysis of choosing stricter and more liberal thresholds to show how they affect our instruments (**Supplementary figure S1**). We coded the effect of the genetic instrument to mimic the drug effect on intermediate phenotype, for example, the effect of lowering systolic blood pressure from ACE inhibitor drugs. We assumed that causal pathways existed from drug target genes to estimated eGFR through SBP, the level of LDL-C, and HbA1c as common primary endpoints for intermediate cardiometabolic phenotypes.

### The outcome variable

eGFR was calculated using the CKD-Epidemiology Collaboration (CKD-Epi) equation [41]. We used GWAS summary statistics from Gorski, et.al. [29] which provided annual decline in kidney function measured using creatinine-based eGFR. The annual decline in eGFR GWAS was obtained from 334,339 individuals (74% European descent) by the CKDGen Consortium [42]. To aid interpretation, we presented the annual eGFR change as a positive value, indicating improved kidney function over time. The formula of eGFR change is 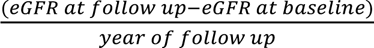. For cross-sectional eGFR and BUN, we used summary statistics from Stanzick, et.al. [30]. A total of 1,201,909 individuals with predominantly European ancestry from both UK Biobank and CKDGen consortium were selected. In cross-sectional eGFR and BUN, positive effect estimates indicate an increase. For eGFR change and eGFR cross-sectional, beta estimates were in log natural (ln). The beta estimates of BUN were obtained by multiplying BUN values by 2.8 and log-transforming them using log natural. The measurement unit for eGFR was ml/min/1.73 m^2^ and for BUN was in mg/dl.

### Drug target Mendelian randomization analysis

We used a two-sample drug target MR approach to analyse the effect of genetically proxied drug target equivalent to reduction in intermediate cardiometabolic phenotypes on the outcome [20]. We estimated the effect of genetically proxied antihypertensive (for example, ACE inhibitor) corresponding to a reduction of 10 mmHg in SBP on annual eGFR change, cross-sectional eGFR, and BUN. Similarly, genetically proxied lipid-lowering drugs (such as statin) and genetically proxied antidiabetic drugs (such as markers for metformin) corresponding to a reduction of 1 mmol/l in LDL-C and 6.7 mmol/mol HbA1c respectively on the outcomes were estimated.

For instruments consisting of only one SNP, we reported the Wald ratio estimate. For instruments consisting of multiple SNPs, we reported the inverse variance weighted (IVW) estimate, which is a meta-analysis of Wald ratio from each SNP, weighted by the inverse variance of association between SNP and the outcome [24]. The instrument variables (IVs) must fulfil three assumptions: 1) the IV must be strongly associated with the exposure, 2) the IV must have no association with confounders of the exposure and the outcome, and 3) there must be no association from the IV to the outcome other than through the exposure (no horizontal pleiotropy) [23, 24]. The first assumption was tested using F-statistics.

Instruments with F-statistics < 10 were dropped from the analysis. The second and third assumptions cannot be proven, albeit several sensitivity analyses were employed to strengthen the causal inference from the MR analysis [24]. The SNPs from exposure variables were harmonized with the outcome summary statistics using information about effect and other alleles and effect allele frequency.

### Sensitivity analyses

We performed several sensitivity analyses using alternative MR methods. These were the simple mode, weighted mode, weighted median, and MR Egger methods. We assessed if heterogeneity between the instruments affected the association by using Cochran’s Q-test statistics [43]. To detect violation of the third MR assumption, we assessed the MR Egger intercept and its p-value [44]. We used positive controls of SBP on hypertension, LDL-C on coronary heart disease (CHD), HbA1c on T2D, annual eGFR decline and cross-sectional eGFR on CKD to validate the direction of effects of the intermediate phenotypes and the outcomes (**Supplementary figure S2**).

The analysis was performed using RStudio version 2023.12.1 Build 402 (R version 4.3.2). For two-sample MR, we used TwoSampleMR package version 0.6.15 [45, 46]. We followed the Strengthening the Reporting of Observational Studies in Epidemiology–Mendelian randomization (STROBE-MR) guideline in reporting our results to ensure reproducibility [47]. The code was made available on Github: <u>remotesuo/drug-target-cardiometabolic-kidney</u>

## Results

### The effect of genetically proxied antihypertensive drugs

In the two-sample MR analyses, we found little evidence of an effect from genetically proxied antihypertensive drugs on annual eGFR change (**Figure 2a**). When we restricted to the European population sample, we observed a small increase on cross-sectional eGFR-creat from the variants proxying the effect of CCB with ln(ml/min/1.73 m^2^) β-estimate 0.006 (95% CI 0.001, 0.011; p-value 0.02) per 10 mmHg decrease of SBP (**Figure 2b.1 to 2b.4**).

**Figure 2.**
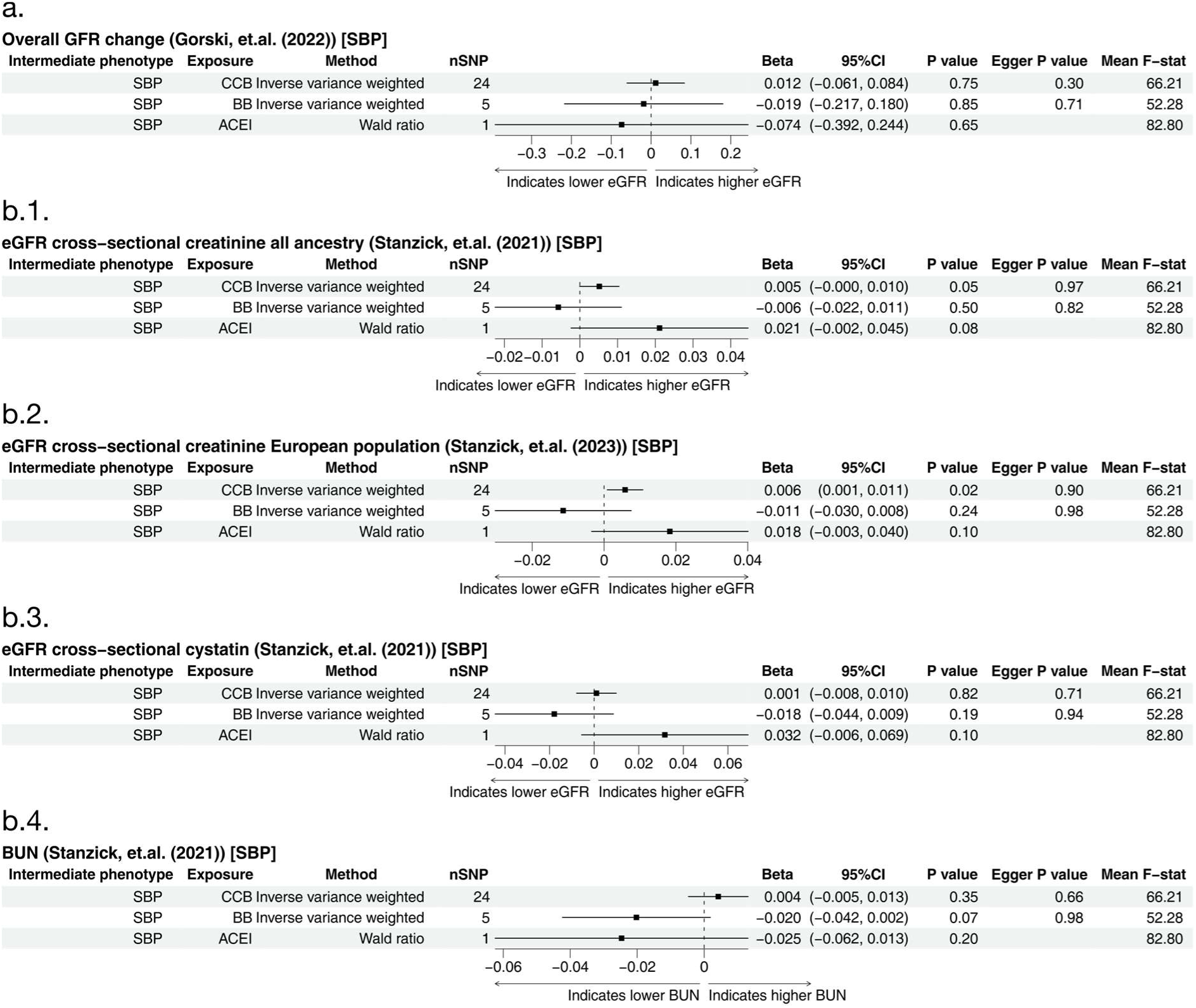
Forest plot for results of genetically proxied antihypertensive drug treatment on kidney function. The estimates show the effect on ln(ml/min/1.73 m^2^) annual eGFR change, ln(ml/min/1.73 m^2^) cross-sectional eGFR, and ln(mg/dl) BUN for each 10 mmHg lowering in SBP. SBP, systolic blood pressure; BUN, blood urea nitrogen; CCB, calcium channel blocker; BB, beta blocker; ACEI, angiotensin-converting enzyme inhibitor; 95% CI, 95% confidence interval; Egger P value, P value for MR Egger intercept. Cystatin-based eGFR and BUN are based in European only populations.

### The effect of genetically proxied lipid-lowering drugs

For genetically proxied lipid-lowering drugs, we found that the variants proxying the effect of PCSK9 inhibitor improved annual eGFR change with ln(ml/min/1.73 m^2^) β-estimate 0.066; 95% CI 0.008, 0.125; p-value 0.03) per 1 mmol/l decrease of LDL-C. In sensitivity analyses genetically proxied PCSK9 inhibitor showed deviation of effect directions (using simple mode, weighted mode, and weighted median) (**Supplementary figures S4-S8**) and heterogeneity (Cochrane Q p-value < 0.05) (**Supplementary figures S3**) on cross-sectional eGFR and BUN. The effect estimates for the other genetically proxied lipid-lowering drugs, statins and NPC1L1 inhibitors, had large CIs with little evidence for an effect on annual eGFR change (**Figure 3a**). Genetically proxied statins reduced cross-sectional eGFR-creat in the all ancestry analysis (ln β-estimate −0.012; 95% CI −0.018, −0.006; p-value < 0.01), eGFR-creat in the European only analysis (ln β-estimate −0.013; 95% CI −0.019, −0.007; p-value < 0.01), and in eGFR-cys (ln β-estimate −0.013; 95% CI −0.023, −0.004; p-value 0.01). Genetically proxied NPC1L1 inhibitors decreased cross-sectional eGFR-creat with ln β-estimate −0.015 (95% CI −0.029, −0.002; p-value 0.03) in all ancestry, and ln β-estimate −0.020 (95% CI −0.033, −0.008; p-value < 0.01) in European only analysis (**Figure 3b.1 to 3b.4**). Potential pleiotropy effects were observed for genetically proxied PCSK9 inhibitors on cross-sectional eGFR-creat and BUN.

**Figure 3.**
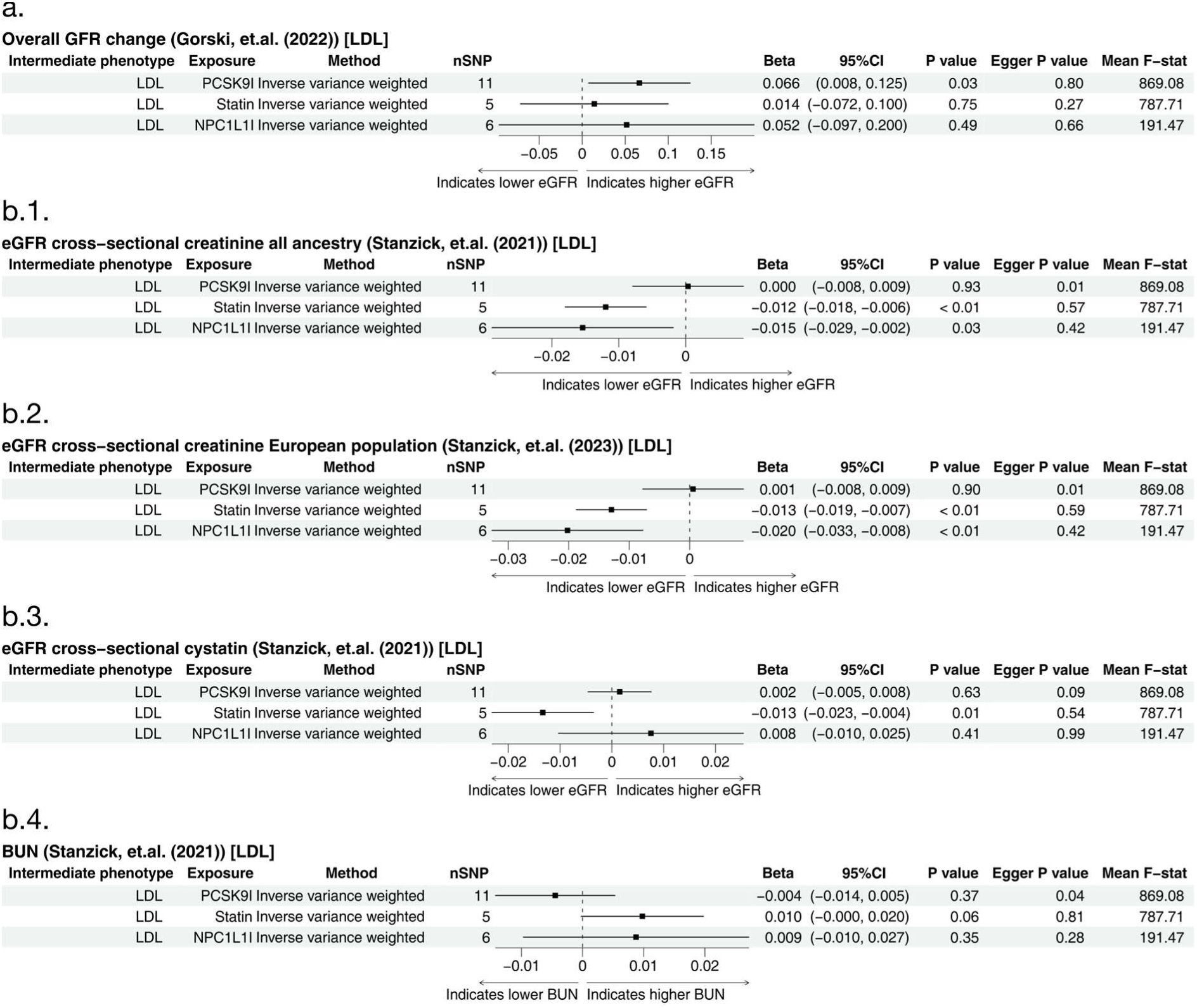
Forest plot for results of genetically proxied lipid-lowering drug treatment on kidney function. The estimates show the effect on ln(ml/min/1.73 m^2^) annual eGFR change, ln(ml/min/1.73 m^2^) cross-sectional eGFR, and ln(mg/dl) BUN for each 1 mmol/l lowering in LDL-C. LDL-C, low-density lipoprotein cholesterol; BUN, blood urea nitrogen HMGCR, 3-Hydroxy-3-Methylglutaryl-CoA Reductase; PCSK9, Proprotein convertase subtilisin/kexin type 9; NPC1L1, Niemann-Pick C1-Like 1; 95% CI, 95% confidence interval; Egger P value, P value for MR Egger intercept. Cystatin-based eGFR and BUN are based in European only populations.

### The effect of genetically proxied antidiabetic drugs

In the group of antidiabetic drugs, genetically proxied SGLT2 inhibitors showed a decrease in annual eGFR change with ln(ml/min/1.73m^2^) β-estimate −0.410 (95% CI −0.698, −0.123; p-value 0.01) per 6.7 mmol/mol decrease of HbA1c (**Figure 4a**). Genetically proxied SGLT2 inhibitors increased eGFR-creat in both the all ancestry and European only analyses with ln β-estimate 0.024 (95% CI 0.004, 0.045; p-value 0.02) and 0.028 (95% CI 0.008, 0.048; p-value < 0.01) respectively. The effect directions for genetically proxied SGLT2 inhibitors were consistent for eGFR-cys (increase effect) and BUN (decrease effect), albeit with CIs crossing the null (**Figure 4b.1 to 4b.4**). For markers of metformin, genetically proxied MC1 increased cross-sectional eGFR-creat with ln β-estimate 0.022 (95% CI 0.001, 0.043 p-value 0.04) in both all ancestry and European only analyses. On cross-sectional eGFR, genetically proxied MC1 showed heterogeneity (**Supplementary figure S3**). GDF15 also increased cross-sectional eGFR-creat in both all ancestry and European only with ln β-estimates 0.094 (95% CI 0.031, 0.156; p-value < 0.01) and 0.100 (95% CI 0.029, 0.172; p-value 0.01) respectively (**Figure 4b.1 to 4b.4**). A marker of metformin, GCG, has F-statistic < 10 and was removed (**Supplementary tables ST2-ST6**). Using Steiger test, GCG also showed a reverse causal direction on annual eGFR change (**Supplementary table ST7**). No conclusive effect on annual eGFR change, cross-sectional eGFR, and BUN was shown by GLP1R agonist (**Figure 4a, and 4b.1 to 4b.4**).

**Figure 4.**
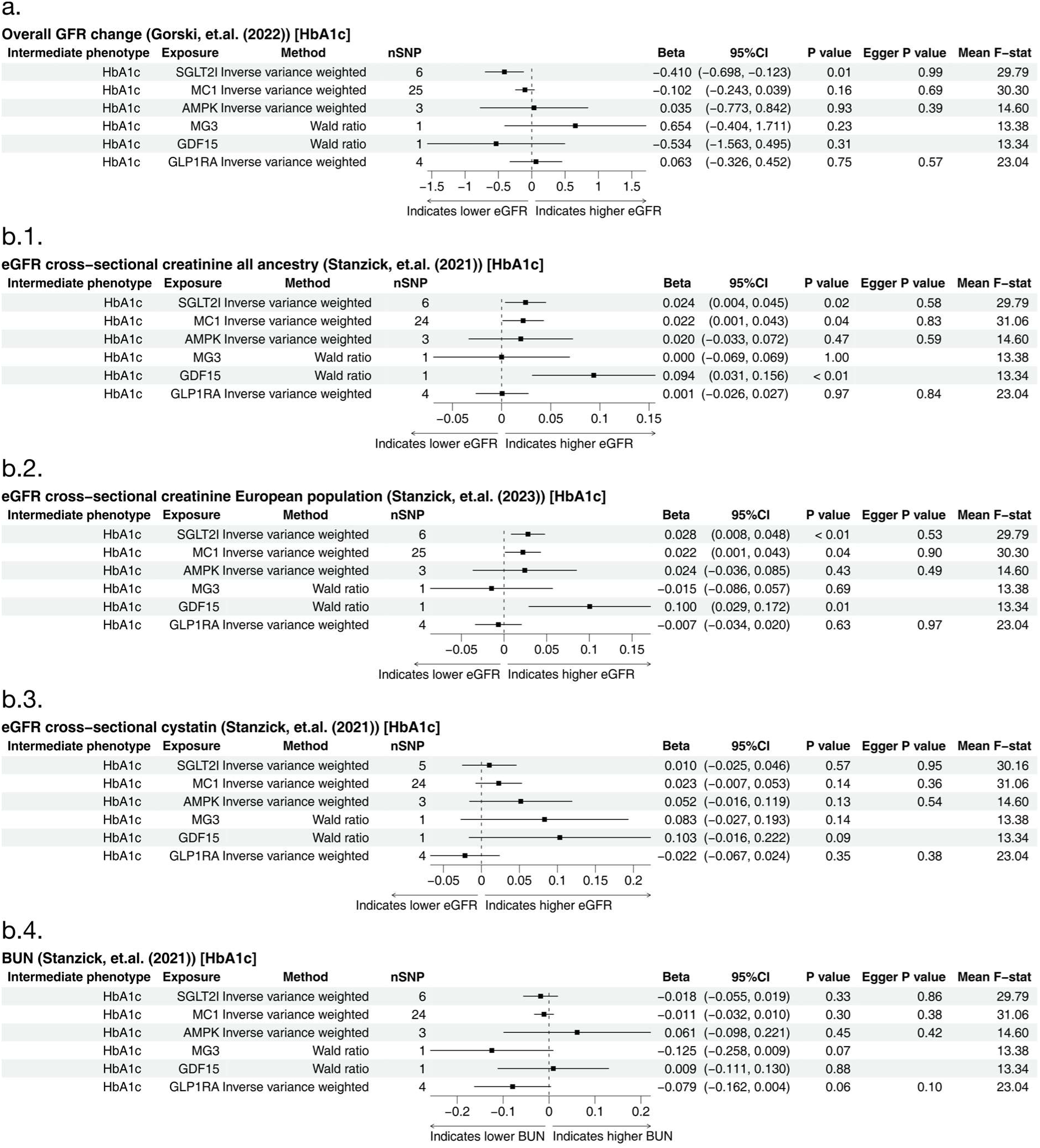
Forest plot for results of genetically proxied antidiabetic drug treatment on kidney function. The estimates show the effect on ln(ml/min/1.73 m^2^) annual eGFR change, ln(ml/min/1.73 m^2^) cross-sectional eGFR, and ln(mg/dl) BUN for each 6.7 mmol/mol lowering in HbA1c. HbA1c, glycated haemoglobin; BUN, blood urea nitrogen; AMPK, AMP-activated protein kinase; MC1, mitochondrial complex 1; MG3, mitochondrial glycerol 3; GDF15, growth differentiation factor 15; GCG, glucagon; GLP1R, glucagon-like peptide 1 receptor; 95% CI, 95% confidence interval; Egger P value, P value for MR Egger intercept. Cystatin-based eGFR and BUN are based in European only populations.

## Discussion

In this study, we have investigated the effect of genetically proxied antihypertensive, antidiabetic, and lipid-lowering drugs on the annual eGFR change, cross-sectional eGFR and BUN in the general population. Our study found that genetically proxied SGLT2 inhibitors, corresponding to reduction in HbA1c, have a favourable effect on cross-sectional eGFR-crea but had limited evidence for an effect on eGFR-cys or BUN and led to a decline in annual eGFR. The effect from markers of metformin were quite heterogenous, with genetically proxied MC1 largely driven a positive effect on cross-sectional eGFR. Genetically proxied GLP1R agonist showed inconclusive effects on the kidney outcome. Overall, there was limited evidence for an effect of SBP on kidney function. However, genetically proxied CCB were found to have a small positive effect on cross-sectional eGFR. The genetically proxied effect from two lipid-lowering drugs: statins and NPC1L1 inhibitor had a negative effect on cross-sectional eGFR-crea, supported by consistent effect direction on BUN. In contrast, genetically proxied PCSK9 inhibitors did not adversely affect eGFR and might even improve annual eGFR change.

We observed conflicting results for genetically proxied SGLT2 inhibitors, as they were estimated to improve cross-sectional eGFR but worsen annual eGFR change. The discrepancy might be due to differences in how eGFR was obtained in these two outcomes with the annual eGFR change consisted of serial eGFR measurements instead of a single timepoint. A decline in eGFR at the beginning of treatment [6] has been reported for patients on SGLT2 inhibitors as compared to those on standard therapy [16, 17, 48]. However, this decline became milder as comparison with the decline in control group over longer period of observation, which indicated favourable kidney function preservation [48, 49]. This might also suggest a non-linear relationship underlying the effect of SGLT2 inhibitors on kidney function. As SGLT2 inhibitors reduce the overall fluid volume by increasing osmolarity inside the kidney tubules due to high glucose concentration, they also improve cardiovascular performance and kidney function [50–52]. This benefit might not be captured in our results as we only used HbA1c to proxy the effect on kidney outcomes.

There was evidence for heterogeneity among the instruments proxying the effect of metformin on cross-sectional eGFR and BUN. The effects of metformin on the kidney are not fully understood and might be independent from glucose-lowering effect [53]. We presented the proxied effect of metformin using only five known markers, of which only genetically proxied MC1 and GDF15 suggested significant effect on cross-sectional eGFR. The MC1 inhibition plays a role in reducing gluconeogenesis which subsequently reduces blood glucose level [54]. The effect of GDF15 might contribute to weight loss and possibly beneficial for the kidney through immunoinflammatory regulation [55, 56]. It is likely that the other metformin markers might also play role in the kidney [57]. Considering its safety for the kidney and cost effectiveness, metformin can be used as first-line antidiabetic drug in patients with reduced kidney function [58, 59]. GLP1R agonist has shown a potential protective effect on the kidney both through indirect effect, such as glycaemic control and direct effect such as by promoting kidney haemodynamic [26]. Our study did not show conclusive effect for this drug target, which suggested the benefit might be through pathways other than HbA1c lowering on eGFR.

The genetically proxied CCB and ACE inhibitors might benefit the kidney indices, albeit the confidence interval contained the null for the latter. An earlier MR study by Zhao et al [25] reported that ACE inhibitors increased, and BBs decreased cross-sectional eGFR. Their study also found favourable effects of CCB on UACR but not on eGFR. Albeit using different instrument selection criteria, their results were consistent with ours [25]. In meta-analyses, antihypertensives were reported to have mixed of positive results [60] and no effect [27]. In a recent clinical trial of CKD patients with eGFR < 30 ml/min/1.73 m^2^, continuing ACE inhibitors was favourable in comparison with stopping the drugs [61]. Our study retrieved only one SNP for ACE inhibitors. It is also plausible that worsening kidney function leads to elevated blood pressure rather than the opposite [62]. However, our directionality test (**Supplementary table ST7**) indicated a correct causal direction of antihypertensives on the kidney function.

Statins and NPC1L1 inhibitors showed reduced effect on cross-sectional eGFR. This was also supported by direction of increase in BUN. A systematic review has previously reported that statins improve renal function, including in those with cardiovascular comorbidities [27]. However, it might not slow CKD progression in patients with renal failure [63]. Statins may benefit the kidney through mechanisms other than reducing eGFR, such as lowering albumin or increasing creatinine clearance [64]. Our results show that PCSK9 inhibitors increase annual eGFR change. Although the point estimates for PCSK9 inhibitors were consistent with a positive effect on cross-sectional eGFR and a decrease in BUN, the confidence intervals contained the null and there was evidence of potential pleiotropy and heterogeneity. A recent clinical trial of alirocumab, a PCSK9 inhibitor, concluded that eGFR was stable for the 3-year trial period and there was no difference in eGFR between treatment and control groups. In this trial, those with eGFR < 30 ml/min/1.73 m^2^ were excluded [65]. A previous meta-analysis has also shown that alirocumab does not affect eGFR [66].

The use of GWAS summary statistics with a large sample size for the kidney function phenotypes is one of the strengths of our study. These data represent the effect of genetically proxied cardiometabolic drugs on kidney function in the general population of mainly European descent. The IVs that we used have also been validated in earlier studies. Only instruments with strong evidence of causality to the intermediate phenotype were used, which met the first assumption of MR. Furthermore, we performed various sensitivity analyses to support our interpretation and help to explain the second and third MR assumptions [24]. Drug target MR may overcome bias that commonly occurred in observational studies [23, 24]. Using drug target MR also limits the IVs specific to the drug target of interest, thus might elude horizontal pleiotropy [20]. The analysis using general population may also avoid bias from conditioning on case-only subjects (a collider bias) [24].

Our study also has limitations. There is a partial sample overlap between the exposure and the outcome GWAS, which may have led to exaggerated effect estimates [67]. However, this bias is unlikely to change the direction of the our effects [68]. Another limitation is that drug target MR is unable to distinguish between drug subclasses and different doses [69]. As the IVs were obtained from different studies, there was variation in selection criteria. However, we performed various clumping thresholds as sensitivity analyses to show how they affected the IVs. The IVs can only explain genetically proxied drug target effect on the kidney that corresponds to the effect of intermediate phenotypes. We also cannot exclude possible survival bias as cardiovascular disease may cause mortality before CKD occurs [23]. While analysing general population may avoid bias from using case-only subjects, the effect may not reflect the experience of CKD patients. Future studies may need to explore the effect of cardiometabolic drugs in CKD patients.

### Conclusions

We found some evidence of a positive but weak effect of genetically proxied SGLT2 inhibitors on cross-sectional eGFR and a reduction in annual eGFR change. This might be postulated as a non-linear effect of the drug on the kidney. Positive effect from genetically proxied MC1 might explain specific pathway of metformin on the kidney. Kidney benefit from GLP1R agonists might need to be explored using more kidney indices than what we currently have. The lack of evidence for an effect of antihypertensive targets on kidney function indices indicates that they are likely safe to use for the kidney. Genetically proxied statins and NPC1L1 inhibitors may decrease cross-sectional eGFR while our findings suggest that genetically proxied PCSK9 inhibitors did not. Genetically proxied PCSK9 inhibitors also showed some improvement in annual eGFR change. Lastly, our study provided insight about potential effect of cardiometabolic drugs in generally healthy population.

## Data Availability

All data produced in the present study are available upon reasonable request to the authors

## Acknowledgements

We thank to the participants of the genetic study and population surveys. We thank to the team in the Chronic Kidney Disease Genetics Consortium, Global Lipids Genetics Consortium, and Medical Research Council Integrative Epidemiology Unit at the University of Bristol for access to the GWAS summary statistics used in this study.

## Author contributions

HR, AA, SB, and BB conceived and design the study. AA and SB analysed the data. AA wrote the first draft of the manuscript. All authors contributed in the interpretation and revision of the manuscript.

## Ethical standards

We used publicly available summary statistics for this study.

