## Supplemental figures and tables for "Cardiometabolic drugs and kidney function – a drug target Mendelian randomisation study"

### Figure S1.a. Various clumping $R^2$ thresholds for exposures on annual eGFR change

**Overall GFR change (Gorski, et.al. (2022)) [SBP]**

| Intermediate phenotype | Exposure | Method | nSNP | Beta | 95%CI | P value | Egger P value | Mean F−stat |
|---|---|---|---|---|---|---|---|---|
| SBP | CCB | Inverse variance weighted | 24 | 0.012 | (−0.061, 0.084) | 0.75 | 0.30 | 66.21 |
| SBP | BB | Inverse variance weighted | 5 | −0.019 | (−0.217, 0.180) | 0.85 | 0.71 | 52.28 |
| SBP | ACEI | Wald ratio | 1 | −0.074 | (−0.392, 0.244) | 0.65 | | 82.80 |

**Overall GFR change (Gorski, et.al. (2022)) [LDL]**

| Intermediate phenotype | Exposure | Method | nSNP | Beta | 95%CI | P value | Egger P value | Mean F−stat |
|---|---|---|---|---|---|---|---|---|
| LDL | PCSK9I | Inverse variance weighted | 12 | 0.066 | (0.009, 0.123) | 0.02 | 0.81 | 841.56 |
| LDL | Statin | Inverse variance weighted | 8 | −0.003 | (−0.080, 0.075) | 0.95 | 0.04 | 654.90 |
| LDL | NPC1L1I | Inverse variance weighted | 6 | 0.052 | (−0.097, 0.200) | 0.49 | 0.66 | 191.47 |

**Overall GFR change (Gorski, et.al. (2022)) [HbA1c]**

| Intermediate phenotype | Exposure | Method | nSNP | Beta | 95%CI | P value | Egger P value | Mean F−stat |
|---|---|---|---|---|---|---|---|---|
| HbA1c | SGLT2I | Inverse variance weighted | 13 | −0.413 | (−0.615, −0.210) | < 0.01 | 0.44 | 28.01 |
| HbA1c | MC1 | Inverse variance weighted | 25 | −0.102 | (−0.243, 0.039) | 0.16 | 0.69 | 30.30 |
| HbA1c | AMPK | Inverse variance weighted | 3 | 0.035 | (−0.773, 0.842) | 0.93 | 0.39 | 14.60 |
| HbA1c | MG3 | Wald ratio | 1 | 0.654 | (−0.404, 1.711) | 0.23 | | 13.38 |
| HbA1c | GDF15 | Wald ratio | 1 | −0.534 | (−1.563, 0.495) | 0.31 | | 13.34 |
| HbA1c | GLP1RA | Inverse variance weighted | 4 | 0.063 | (−0.326, 0.452) | 0.75 | 0.57 | 23.04 |

Clumping 250 kb, $R^2$ 0.8

**Overall GFR change (Gorski, et.al. (2022)) [SBP]**

| Intermediate phenotype | Exposure | Method | nSNP | Beta | 95%CI | P value | Egger P value | Mean F−stat |
|---|---|---|---|---|---|---|---|---|
| SBP | CCB | Inverse variance weighted | 24 | 0.012 | (−0.061, 0.084) | 0.75 | 0.30 | 66.21 |
| SBP | BB | Inverse variance weighted | 5 | −0.019 | (−0.217, 0.180) | 0.85 | 0.71 | 52.28 |
| SBP | ACEI | Wald ratio | 1 | −0.074 | (−0.392, 0.244) | 0.65 | | 82.80 |

**Overall GFR change (Gorski, et.al. (2022)) [LDL]**

| Intermediate phenotype | Exposure | Method | nSNP | Beta | 95%CI | P value | Egger P value | Mean F−stat |
|---|---|---|---|---|---|---|---|---|
| LDL | PCSK9I | Inverse variance weighted | 11 | 0.066 | (0.008, 0.125) | 0.03 | 0.80 | 869.08 |
| LDL | Statin | Inverse variance weighted | 5 | 0.014 | (−0.072, 0.100) | 0.75 | 0.27 | 787.71 |
| LDL | NPC1L1I | Inverse variance weighted | 6 | 0.052 | (−0.097, 0.200) | 0.49 | 0.66 | 191.47 |

**Overall GFR change (Gorski, et.al. (2022)) [HbA1c]**

| Intermediate phenotype | Exposure | Method | nSNP | Beta | 95%CI | P value | Egger P value | Mean F−stat |
|---|---|---|---|---|---|---|---|---|
| HbA1c | SGLT2I | Inverse variance weighted | 6 | −0.410 | (−0.698, −0.123) | 0.01 | 0.99 | 29.79 |
| HbA1c | MC1 | Inverse variance weighted | 25 | −0.102 | (−0.243, 0.039) | 0.16 | 0.69 | 30.30 |
| HbA1c | AMPK | Inverse variance weighted | 3 | 0.035 | (−0.773, 0.842) | 0.93 | 0.39 | 14.60 |
| HbA1c | MG3 | Wald ratio | 1 | 0.654 | (−0.404, 1.711) | 0.23 | | 13.38 |
| HbA1c | GDF15 | Wald ratio | 1 | −0.534 | (−1.563, 0.495) | 0.31 | | 13.34 |
| HbA1c | GLP1RA | Inverse variance weighted | 4 | 0.063 | (−0.326, 0.452) | 0.75 | 0.57 | 23.04 |

Clumping 250 kb, $R^2$ 0.4

**Overall GFR change (Gorski, et.al. (2022)) [SBP]**

| Intermediate phenotype | Exposure | Method | nSNP | Beta | 95%CI | P value | Egger P value | Mean F−stat |
|---|---|---|---|---|---|---|---|---|
| SBP | CCB | Inverse variance weighted | 24 | 0.012 | (−0.061, 0.084) | 0.75 | 0.30 | 66.21 |
| SBP | BB | Inverse variance weighted | 5 | −0.019 | (−0.217, 0.180) | 0.85 | 0.71 | 52.28 |
| SBP | ACEI | Wald ratio | 1 | −0.074 | (−0.392, 0.244) | 0.65 | | 82.80 |

**Overall GFR change (Gorski, et.al. (2022)) [LDL]**

| Intermediate phenotype | Exposure | Method | nSNP | Beta | 95%CI | P value | Egger P value | Mean F−stat |
|---|---|---|---|---|---|---|---|---|
| LDL | PCSK9I | Inverse variance weighted | 11 | 0.066 | (0.008, 0.125) | 0.03 | 0.80 | 869.08 |
| LDL | Statin | Inverse variance weighted | 5 | −0.001 | (−0.117, 0.114) | 0.98 | 0.11 | 659.99 |
| LDL | NPC1L1I | Inverse variance weighted | 4 | 0.052 | (−0.117, 0.220) | 0.55 | 0.60 | 218.35 |

**Overall GFR change (Gorski, et.al. (2022)) [HbA1c]**

| Intermediate phenotype | Exposure | Method | nSNP | Beta | 95%CI | P value | Egger P value | Mean F−stat |
|---|---|---|---|---|---|---|---|---|
| HbA1c | SGLT2I | Inverse variance weighted | 4 | −0.442 | (−0.779, −0.106) | 0.01 | 0.83 | 33.37 |
| HbA1c | MC1 | Inverse variance weighted | 25 | −0.102 | (−0.243, 0.039) | 0.16 | 0.69 | 30.30 |
| HbA1c | AMPK | Inverse variance weighted | 3 | 0.035 | (−0.773, 0.842) | 0.93 | 0.39 | 14.60 |
| HbA1c | MG3 | Wald ratio | 1 | 0.654 | (−0.404, 1.711) | 0.23 | | 13.38 |
| HbA1c | GDF15 | Wald ratio | 1 | −0.534 | (−1.563, 0.495) | 0.31 | | 13.34 |
| HbA1c | GLP1RA | Inverse variance weighted | 4 | 0.063 | (−0.326, 0.452) | 0.75 | 0.57 | 23.04 |

Clumping 250 kb, $R^2$ 0.2

**Overall GFR change (Gorski, et.al. (2022)) [SBP]**

| Intermediate phenotype | Exposure | Method | nSNP | Beta | 95%CI | P value | Egger P value | Mean F−stat |
|---|---|---|---|---|---|---|---|---|
| SBP | CCB | Inverse variance weighted | 24 | 0.012 | (−0.061, 0.084) | 0.75 | 0.30 | 66.21 |
| SBP | BB | Inverse variance weighted | 5 | −0.019 | (−0.217, 0.180) | 0.85 | 0.71 | 52.28 |
| SBP | ACEI | Wald ratio | 1 | −0.074 | (−0.392, 0.244) | 0.65 | | 82.80 |

**Overall GFR change (Gorski, et.al. (2022)) [LDL]**

| Intermediate phenotype | Exposure | Method | nSNP | Beta | 95%CI | P value | Egger P value | Mean F−stat |
|---|---|---|---|---|---|---|---|---|
| LDL | PCSK9I | Inverse variance weighted | 8 | 0.047 | (−0.016, 0.111) | 0.14 | 0.40 | 1054.56 |
| LDL | Statin | Inverse variance weighted | 2 | 0.003 | (−0.234, 0.241) | 0.98 | | 1022.88 |
| LDL | NPC1L1I | Inverse variance weighted | 2 | 0.108 | (−0.118, 0.334) | 0.35 | | 236.71 |

**Overall GFR change (Gorski, et.al. (2022)) [HbA1c]**

| Intermediate phenotype | Exposure | Method | nSNP | Beta | 95%CI | P value | Egger P value | Mean F−stat |
|---|---|---|---|---|---|---|---|---|
| HbA1c | SGLT2I | Inverse variance weighted | 4 | −0.442 | (−0.779, −0.106) | 0.01 | 0.83 | 33.37 |
| HbA1c | MC1 | Inverse variance weighted | 25 | −0.102 | (−0.243, 0.039) | 0.16 | 0.69 | 30.30 |
| HbA1c | AMPK | Inverse variance weighted | 3 | 0.035 | (−0.773, 0.842) | 0.93 | 0.39 | 14.60 |
| HbA1c | MG3 | Wald ratio | 1 | 0.654 | (−0.404, 1.711) | 0.23 | | 13.38 |
| HbA1c | GDF15 | Wald ratio | 1 | −0.534 | (−1.563, 0.495) | 0.31 | | 13.34 |
| HbA1c | GLP1RA | Inverse variance weighted | 3 | 0.051 | (−0.392, 0.494) | 0.82 | 0.63 | 22.90 |

Clumping 250 kb, $R^2$ 0.1

**Overall GFR change (Gorski, et.al. (2022)) [SBP]**

| Intermediate phenotype | Exposure | Method | nSNP | Beta | 95%CI | P value | Egger P value | Mean F−stat |
|---|---|---|---|---|---|---|---|---|
| SBP | CCB | Inverse variance weighted | 7 | 0.026 | (−0.076, 0.128) | 0.62 | 0.24 | 116.66 |
| SBP | BB | Wald ratio | 1 | 0.018 | (−0.322, 0.358) | 0.92 | | 72.78 |
| SBP | ACEI | Wald ratio | 1 | −0.074 | (−0.392, 0.244) | 0.65 | | 82.80 |

**Overall GFR change (Gorski, et.al. (2022)) [LDL]**

| Intermediate phenotype | Exposure | Method | nSNP | Beta | 95%CI | P value | Egger P value | Mean F−stat |
|---|---|---|---|---|---|---|---|---|
| LDL | PCSK9I | Inverse variance weighted | 4 | 0.053 | (−0.034, 0.141) | 0.23 | 0.48 | 1480.35 |
| LDL | Statin | Wald ratio | 1 | 0.047 | (−0.087, 0.182) | 0.49 | | 1849.06 |
| LDL | NPC1L1I | Wald ratio | 1 | 0.122 | (−0.122, 0.365) | 0.33 | | 409.21 |

**Overall GFR change (Gorski, et.al. (2022)) [HbA1c]**

| Intermediate phenotype | Exposure | Method | nSNP | Beta | 95%CI | P value | Egger P value | Mean F−stat |
|---|---|---|---|---|---|---|---|---|
| HbA1c | SGLT2I | Inverse variance weighted | 2 | −0.356 | (−0.806, 0.095) | 0.12 | | 38.88 |
| HbA1c | MC1 | Inverse variance weighted | 25 | −0.102 | (−0.243, 0.039) | 0.16 | 0.69 | 30.30 |
| HbA1c | AMPK | Inverse variance weighted | 3 | 0.035 | (−0.773, 0.842) | 0.93 | 0.39 | 14.60 |
| HbA1c | MG3 | Wald ratio | 1 | 0.654 | (−0.404, 1.711) | 0.23 | | 13.38 |
| HbA1c | GDF15 | Wald ratio | 1 | −0.534 | (−1.563, 0.495) | 0.31 | | 13.34 |
| HbA1c | GLP1RA | Inverse variance weighted | 3 | 0.051 | (−0.392, 0.494) | 0.82 | 0.63 | 22.90 |

Clumping 250 kb, $R^2$ 0.01

### Figure S1.b. Various clumping $R^2$ thresholds for exposures on cross-sectional eGFR all ancestry

**eGFR cross-sectional creatinine all ancestry (Stanzick, et.al. (2021)) [SBP]**

| Intermediate phenotype | Exposure | Method | nSNP | Beta | 95%CI | P value | Egger P value | Mean F-stat |
|---|---|---|---|---|---|---|---|---|
| SBP | CCB | Inverse variance weighted | 24 | 0.005 | (−0.000, 0.010) | 0.05 | 0.97 | 66.21 |
| SBP | BB | Inverse variance weighted | 5 | −0.006 | (−0.022, 0.011) | 0.50 | 0.82 | 52.28 |
| SBP | ACEI | Wald ratio | 1 | 0.021 | (−0.002, 0.045) | 0.08 | | 82.80 |

**eGFR cross-sectional creatinine all ancestry (Stanzick, et.al. (2021)) [LDL]**

| Intermediate phenotype | Exposure | Method | nSNP | Beta | 95%CI | P value | Egger P value | Mean F-stat |
|---|---|---|---|---|---|---|---|---|
| LDL | PCSK9I | Inverse variance weighted | 12 | 0.000 | (−0.007, 0.008) | 0.91 | 0.01 | 841.56 |
| LDL | Statin | Inverse variance weighted | 8 | −0.011 | (−0.017, −0.006) | < 0.01 | 0.23 | 654.90 |
| LDL | NPC1L1I | Inverse variance weighted | 6 | −0.015 | (−0.029, −0.002) | 0.03 | 0.42 | 191.47 |

**eGFR cross-sectional creatinine all ancestry (Stanzick, et.al. (2021)) [HbA1c]**

| Intermediate phenotype | Exposure | Method | nSNP | Beta | 95%CI | P value | Egger P value | Mean F-stat |
|---|---|---|---|---|---|---|---|---|
| HbA1c | SGLT2I | Inverse variance weighted | 12 | 0.026 | (0.011, 0.042) | < 0.01 | 0.36 | 28.16 |
| HbA1c | MC1 | Inverse variance weighted | 24 | 0.022 | (0.001, 0.043) | 0.04 | 0.83 | 31.06 |
| HbA1c | AMPK | Inverse variance weighted | 3 | 0.020 | (−0.033, 0.072) | 0.47 | 0.59 | 14.60 |
| HbA1c | MG3 | Wald ratio | 1 | 0.000 | (−0.069, 0.069) | 1.00 | | 13.38 |
| HbA1c | GDF15 | Wald ratio | 1 | 0.094 | (0.031, 0.156) | < 0.01 | | 13.34 |
| HbA1c | GLP1RA | Inverse variance weighted | 4 | 0.001 | (−0.026, 0.027) | 0.97 | 0.84 | 23.04 |

Clumping 250 kb, $R^2$ 0.8

**eGFR cross-sectional creatinine all ancestry (Stanzick, et.al. (2021)) [SBP]**

| Intermediate phenotype | Exposure | Method | nSNP | Beta | 95%CI | P value | Egger P value | Mean F-stat |
|---|---|---|---|---|---|---|---|---|
| SBP | CCB | Inverse variance weighted | 24 | 0.005 | (−0.000, 0.010) | 0.05 | 0.97 | 66.21 |
| SBP | BB | Inverse variance weighted | 5 | −0.006 | (−0.022, 0.011) | 0.50 | 0.82 | 52.28 |
| SBP | ACEI | Wald ratio | 1 | 0.021 | (−0.002, 0.045) | 0.08 | | 82.80 |

**eGFR cross-sectional creatinine all ancestry (Stanzick, et.al. (2021)) [LDL]**

| Intermediate phenotype | Exposure | Method | nSNP | Beta | 95%CI | P value | Egger P value | Mean F-stat |
|---|---|---|---|---|---|---|---|---|
| LDL | PCSK9I | Inverse variance weighted | 11 | 0.000 | (−0.008, 0.009) | 0.93 | 0.01 | 869.08 |
| LDL | Statin | Inverse variance weighted | 5 | −0.012 | (−0.018, −0.006) | < 0.01 | 0.57 | 787.71 |
| LDL | NPC1L1I | Inverse variance weighted | 6 | −0.015 | (−0.029, −0.002) | 0.03 | 0.42 | 191.47 |

**eGFR cross-sectional creatinine all ancestry (Stanzick, et.al. (2021)) [HbA1c]**

| Intermediate phenotype | Exposure | Method | nSNP | Beta | 95%CI | P value | Egger P value | Mean F-stat |
|---|---|---|---|---|---|---|---|---|
| HbA1c | SGLT2I | Inverse variance weighted | 6 | 0.024 | (0.004, 0.045) | 0.02 | 0.58 | 29.79 |
| HbA1c | MC1 | Inverse variance weighted | 24 | 0.022 | (0.001, 0.043) | 0.04 | 0.83 | 31.06 |
| HbA1c | AMPK | Inverse variance weighted | 3 | 0.020 | (−0.033, 0.072) | 0.47 | 0.59 | 14.60 |
| HbA1c | MG3 | Wald ratio | 1 | 0.000 | (−0.069, 0.069) | 1.00 | | 13.38 |
| HbA1c | GDF15 | Wald ratio | 1 | 0.094 | (0.031, 0.156) | < 0.01 | | 13.34 |
| HbA1c | GLP1RA | Inverse variance weighted | 4 | 0.001 | (−0.026, 0.027) | 0.97 | 0.84 | 23.04 |

Clumping 250 kb, $R^2$ 0.4

**eGFR cross-sectional creatinine all ancestry (Stanzick, et.al. (2021)) [SBP]**

| Intermediate phenotype | Exposure | Method | nSNP | Beta | 95%CI | P value | Egger P value | Mean F-stat |
|---|---|---|---|---|---|---|---|---|
| SBP | CCB | Inverse variance weighted | 24 | 0.005 | (−0.000, 0.010) | 0.05 | 0.97 | 66.21 |
| SBP | BB | Inverse variance weighted | 5 | −0.006 | (−0.022, 0.011) | 0.50 | 0.82 | 52.28 |
| SBP | ACEI | Wald ratio | 1 | 0.021 | (−0.002, 0.045) | 0.08 | | 82.80 |

**eGFR cross-sectional creatinine all ancestry (Stanzick, et.al. (2021)) [LDL]**

| Intermediate phenotype | Exposure | Method | nSNP | Beta | 95%CI | P value | Egger P value | Mean F-stat |
|---|---|---|---|---|---|---|---|---|
| LDL | PCSK9I | Inverse variance weighted | 11 | 0.000 | (−0.008, 0.009) | 0.93 | 0.01 | 869.08 |
| LDL | Statin | Inverse variance weighted | 5 | −0.012 | (−0.019, −0.006) | < 0.01 | 0.30 | 659.99 |
| LDL | NPC1L1I | Inverse variance weighted | 4 | −0.017 | (−0.034, 0.001) | 0.07 | 0.64 | 218.35 |

**eGFR cross-sectional creatinine all ancestry (Stanzick, et.al. (2021)) [HbA1c]**

| Intermediate phenotype | Exposure | Method | nSNP | Beta | 95%CI | P value | Egger P value | Mean F-stat |
|---|---|---|---|---|---|---|---|---|
| HbA1c | SGLT2I | Inverse variance weighted | 4 | 0.025 | (0.000, 0.049) | 0.05 | 0.56 | 33.37 |
| HbA1c | MC1 | Inverse variance weighted | 24 | 0.022 | (0.001, 0.043) | 0.04 | 0.83 | 31.06 |
| HbA1c | AMPK | Inverse variance weighted | 3 | 0.020 | (−0.033, 0.072) | 0.47 | 0.59 | 14.60 |
| HbA1c | MG3 | Wald ratio | 1 | 0.000 | (−0.069, 0.069) | 1.00 | | 13.38 |
| HbA1c | GDF15 | Wald ratio | 1 | 0.094 | (0.031, 0.156) | < 0.01 | | 13.34 |
| HbA1c | GLP1RA | Inverse variance weighted | 4 | 0.001 | (−0.026, 0.027) | 0.97 | 0.84 | 23.04 |

Clumping 250 kb, $R^2$ 0.2

**eGFR cross-sectional creatinine all ancestry (Stanzick, et.al. (2021)) [SBP]**

| Intermediate phenotype | Exposure | Method | nSNP | Beta | 95%CI | P value | Egger P value | Mean F-stat |
|---|---|---|---|---|---|---|---|---|
| SBP | CCB | Inverse variance weighted | 24 | 0.005 | (−0.000, 0.010) | 0.05 | 0.97 | 66.21 |
| SBP | BB | Inverse variance weighted | 5 | −0.006 | (−0.022, 0.011) | 0.50 | 0.82 | 52.28 |
| SBP | ACEI | Wald ratio | 1 | 0.021 | (−0.002, 0.045) | 0.08 | | 82.80 |

**eGFR cross-sectional creatinine all ancestry (Stanzick, et.al. (2021)) [LDL]**

| Intermediate phenotype | Exposure | Method | nSNP | Beta | 95%CI | P value | Egger P value | Mean F-stat |
|---|---|---|---|---|---|---|---|---|
| LDL | PCSK9I | Inverse variance weighted | 8 | −0.004 | (−0.009, 0.002) | 0.19 | 0.03 | 1054.56 |
| LDL | Statin | Inverse variance weighted | 2 | −0.012 | (−0.023, 0.000) | 0.05 | | 1022.88 |
| LDL | NPC1L1I | Inverse variance weighted | 2 | −0.008 | (−0.030, 0.014) | 0.48 | | 236.71 |

**eGFR cross-sectional creatinine all ancestry (Stanzick, et.al. (2021)) [HbA1c]**

| Intermediate phenotype | Exposure | Method | nSNP | Beta | 95%CI | P value | Egger P value | Mean F-stat |
|---|---|---|---|---|---|---|---|---|
| HbA1c | SGLT2I | Inverse variance weighted | 4 | 0.025 | (0.000, 0.049) | 0.05 | 0.56 | 33.37 |
| HbA1c | MC1 | Inverse variance weighted | 24 | 0.022 | (0.001, 0.043) | 0.04 | 0.83 | 31.06 |
| HbA1c | AMPK | Inverse variance weighted | 3 | 0.020 | (−0.033, 0.072) | 0.47 | 0.59 | 14.60 |
| HbA1c | MG3 | Wald ratio | 1 | 0.000 | (−0.069, 0.069) | 1.00 | | 13.38 |
| HbA1c | GDF15 | Wald ratio | 1 | 0.094 | (0.031, 0.156) | < 0.01 | | 13.34 |
| HbA1c | GLP1RA | Inverse variance weighted | 3 | 0.006 | (−0.024, 0.036) | 0.68 | 0.80 | 22.90 |

Clumping 250 kb, $R^2$ 0.1

**eGFR cross-sectional creatinine all ancestry (Stanzick, et.al. (2021)) [SBP]**

| Intermediate phenotype | Exposure | Method | nSNP | Beta | 95%CI | P value | Egger P value | Mean F-stat |
|---|---|---|---|---|---|---|---|---|
| SBP | CCB | Inverse variance weighted | 7 | 0.009 | (−0.001, 0.018) | 0.07 | 0.99 | 116.66 |
| SBP | BB | Wald ratio | 1 | −0.022 | (−0.046, 0.002) | 0.08 | | 72.78 |
| SBP | ACEI | Wald ratio | 1 | 0.021 | (−0.002, 0.045) | 0.08 | | 82.80 |

**eGFR cross-sectional creatinine all ancestry (Stanzick, et.al. (2021)) [LDL]**

| Intermediate phenotype | Exposure | Method | nSNP | Beta | 95%CI | P value | Egger P value | Mean F-stat |
|---|---|---|---|---|---|---|---|---|
| LDL | PCSK9I | Inverse variance weighted | 4 | −0.005 | (−0.012, 0.002) | 0.18 | 0.15 | 1480.35 |
| LDL | Statin | Wald ratio | 1 | −0.014 | (−0.023, −0.005) | < 0.01 | | 1849.06 |
| LDL | NPC1L1I | Wald ratio | 1 | −0.012 | (−0.027, 0.004) | 0.14 | | 409.21 |

**eGFR cross-sectional creatinine all ancestry (Stanzick, et.al. (2021)) [HbA1c]**

| Intermediate phenotype | Exposure | Method | nSNP | Beta | 95%CI | P value | Egger P value | Mean F-stat |
|---|---|---|---|---|---|---|---|---|
| HbA1c | SGLT2I | Inverse variance weighted | 2 | 0.019 | (−0.012, 0.050) | 0.24 | | 38.88 |
| HbA1c | MC1 | Inverse variance weighted | 24 | 0.022 | (0.001, 0.043) | 0.04 | 0.83 | 31.06 |
| HbA1c | AMPK | Inverse variance weighted | 3 | 0.020 | (−0.033, 0.072) | 0.47 | 0.59 | 14.60 |
| HbA1c | MG3 | Wald ratio | 1 | 0.000 | (−0.069, 0.069) | 1.00 | | 13.38 |
| HbA1c | GDF15 | Wald ratio | 1 | 0.094 | (0.031, 0.156) | < 0.01 | | 13.34 |
| HbA1c | GLP1RA | Inverse variance weighted | 3 | 0.006 | (−0.024, 0.036) | 0.68 | 0.80 | 22.90 |

Clumping 250 kb, $R^2$ 0.01

### Figure S2. Positive controls to validate the causal effects of exposures on intermediate phenotypes.

#### SBP on hypertension

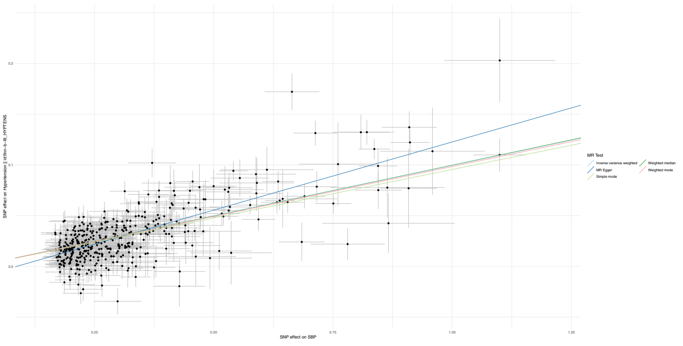


#### LDL cholesterol on coronary heart disease

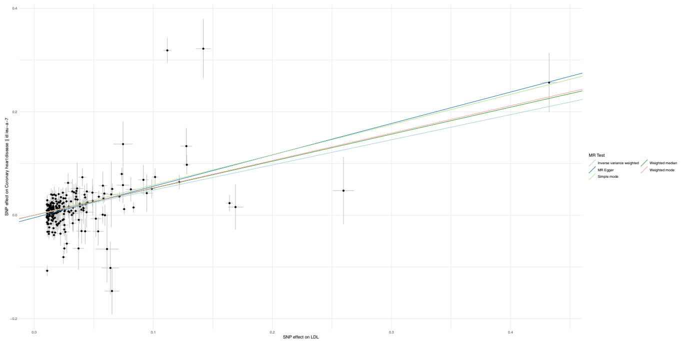


#### HbA1c on type 2 diabetes

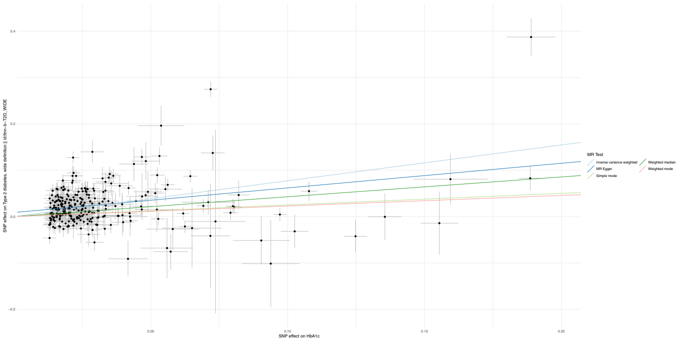


#### Annual eGFR <u>decline</u> on chronic kidney disease

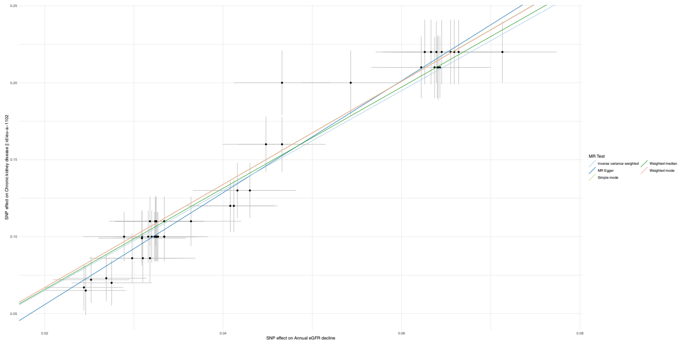


* eGFR decline <u>positive</u> value = a <u>decline</u> in annual eGFR

#### Cross-sectional eGFR on chronic kidney disease

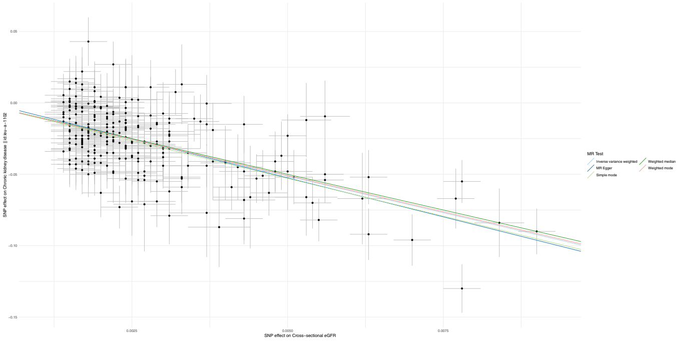


* Cross-sectional eGFR increase = an increase in eGFR

### Figure S3. Relationships between all intermediate phenotype instruments and kidney outcomes.

**Overall GFR change (Gorski, et.al. (2022))**

| IntermediatePheno | exposure | method | nsnp | Beta | 95%CI | pval | RSSobs | Phet | Q | Q_pval | egger_intercept | egger_pval | MeanFstat |
|---|---|---|---|---|---|---|---|---|---|---|---|---|---|
| HbA1c | Lowering HbA1c | Inverse variance weighted | 287 | 0.016 | (−0.005, 0.037) | 0.13 | | | 291.76 | 0.39 | 0 | 0.02 | 123.20 |
| HbA1c | MC1 | Inverse variance weighted | 25 | −0.102 | (−0.243, 0.039) | 0.16 | | | 24.76 | 0.42 | 0 | 0.69 | 30.30 |
| HbA1c | SGLT2I | Inverse variance weighted | 6 | −0.410 | (−0.698, −0.123) | 0.01 | | | 0.55 | 0.99 | 0 | 0.99 | 29.79 |
| HbA1c | GLP1RA | Inverse variance weighted | 4 | 0.063 | (−0.326, 0.452) | 0.75 | | | 0.45 | 0.93 | 0 | 0.57 | 23.04 |
| HbA1c | AMPK | Inverse variance weighted | 3 | 0.035 | (−0.773, 0.842) | 0.93 | | | 3.89 | 0.14 | 0.02 | 0.39 | 14.60 |
| HbA1c | MG3 | Wald ratio | 1 | 0.654 | (−0.404, 1.711) | 0.23 | | | | | | | 13.38 |
| HbA1c | GCG | Wald ratio | 1 | −1.749 | (−3.299, −0.199) | 0.03 | | | | | | | 4.12 |
| HbA1c | GDF15 | Wald ratio | 1 | −0.534 | (−1.563, 0.495) | 0.31 | | | | | | | 13.34 |
| LDL | Lowering LDL | Inverse variance weighted | 229 | −0.002 | (−0.025, 0.022) | 0.88 | | | 243.72 | 0.23 | 0 | 0.01 | 240.65 |
| LDL | PCSK9I | Inverse variance weighted | 11 | 0.066 | (0.008, 0.125) | 0.03 | | | 8.97 | 0.54 | 0 | 0.8 | 869.08 |
| LDL | NPC1L1I | Inverse variance weighted | 6 | 0.052 | (−0.097, 0.200) | 0.49 | | | 0.63 | 0.99 | 0 | 0.66 | 191.47 |
| LDL | Statin | Inverse variance weighted | 5 | 0.014 | (−0.072, 0.100) | 0.75 | | | 2.78 | 0.6 | −0.02 | 0.27 | 787.71 |
| SBP | Lowering SBP | Inverse variance weighted | 440 | 0.045 | (0.025, 0.064) | 0.00 | | | 676.52 | 0 | 0.02 | 0 | 75.13 |
| SBP | CCB | Inverse variance weighted | 24 | 0.012 | (−0.061, 0.084) | 0.75 | | | 21.48 | 0.55 | −0.03 | 0.3 | 66.21 |
| SBP | BB | Inverse variance weighted | 5 | −0.019 | (−0.217, 0.180) | 0.85 | | | 4.75 | 0.31 | 0.06 | 0.71 | 52.28 |
| SBP | ACEI | Wald ratio | 1 | −0.074 | (−0.392, 0.244) | 0.65 | | | | | | | 82.80 |

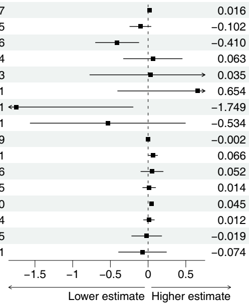


**eGFR cross-sectional creatinine all ancestry (Stanzick, et.al. (2021))**

| IntermediatePheno | exposure | method | nsnp | Beta | 95%CI | pval | RSSobs | Phet | Q | Q_pval | egger_intercept | egger_pval | MeanFstat |
|---|---|---|---|---|---|---|---|---|---|---|---|---|---|
| HbA1c | Lowering HbA1c | Inverse variance weighted | 276 | 0.000 | (−0.003, 0.004) | 0.80 | | | 1819.19 | 0 | 0 | 0.76 | 123.88 |
| HbA1c | MC1 | Inverse variance weighted | 24 | 0.022 | (0.001, 0.043) | 0.04 | | | 114.36 | 0 | 0 | 0.83 | 31.06 |
| HbA1c | SGLT2I | Inverse variance weighted | 6 | 0.024 | (0.004, 0.045) | 0.02 | | | 0.99 | 0.96 | 0 | 0.58 | 29.79 |
| HbA1c | GLP1RA | Inverse variance weighted | 4 | 0.001 | (−0.026, 0.027) | 0.97 | | | 0.78 | 0.86 | 0 | 0.84 | 23.04 |
| HbA1c | AMPK | Inverse variance weighted | 3 | 0.020 | (−0.033, 0.072) | 0.47 | | | 3.74 | 0.15 | 0 | 0.59 | 14.60 |
| HbA1c | MG3 | Wald ratio | 1 | 0.000 | (−0.069, 0.069) | 1.00 | | | | | | | 13.38 |
| HbA1c | GCG | Wald ratio | 1 | −0.092 | (−0.222, 0.038) | 0.17 | | | | | | | 4.12 |
| HbA1c | GDF15 | Wald ratio | 1 | 0.094 | (0.031, 0.156) | 0.00 | | | | | | | 13.34 |
| LDL | Lowering LDL | Inverse variance weighted | 224 | −0.006 | (−0.010, −0.002) | 0.00 | | | 1451.12 | 0 | 0 | 0.95 | 243.54 |
| LDL | PCSK9I | Inverse variance weighted | 11 | 0.000 | (−0.008, 0.009) | 0.93 | | | 51.67 | 0 | 0 | 0.01 | 869.08 |
| LDL | NPC1L1I | Inverse variance weighted | 6 | −0.015 | (−0.029, −0.002) | 0.03 | | | 8.94 | 0.11 | 0 | 0.42 | 191.47 |
| LDL | Statin | Inverse variance weighted | 5 | −0.012 | (−0.018, −0.006) | 0.00 | | | 1.3 | 0.86 | 0 | 0.57 | 787.71 |
| SBP | Lowering SBP | Inverse variance weighted | 436 | 0.003 | (−0.001, 0.006) | 0.16 | | | 5015.1 | 0 | 0 | 0.42 | 75.13 |
| SBP | CCB | Inverse variance weighted | 24 | 0.005 | (−0.000, 0.010) | 0.05 | | | 25.49 | 0.33 | 0 | 0.97 | 66.21 |
| SBP | BB | Inverse variance weighted | 5 | −0.006 | (−0.022, 0.011) | 0.50 | | | 7.49 | 0.11 | 0 | 0.82 | 52.28 |
| SBP | ACEI | Wald ratio | 1 | 0.021 | (−0.002, 0.045) | 0.08 | | | | | | | 82.80 |

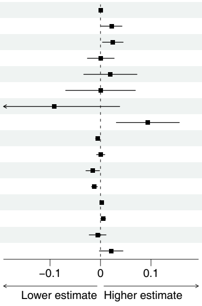


**eGFR cross-sectional creatinine European population (Stanzick, et.al. (2023))**

| IntermediatePheno | exposure | method | nsnp | Beta | 95%CI | pval | RSSobs | Phet | Q | Q_pval | egger_intercept | egger_pval | MeanFstat |
|---|---|---|---|---|---|---|---|---|---|---|---|---|---|
| HbA1c | Lowering HbA1c | Inverse variance weighted | 281 | 0.000 | (−0.003, 0.004) | 0.86 | | | 2010.88 | 0 | 0 | 0.98 | 124.60 |
| HbA1c | MC1 | Inverse variance weighted | 25 | 0.022 | (0.001, 0.043) | 0.04 | | | 110.52 | 0 | 0 | 0.9 | 30.30 |
| HbA1c | SGLT2I | Inverse variance weighted | 6 | 0.028 | (0.008, 0.048) | 0.00 | | | 1.25 | 0.94 | 0 | 0.53 | 29.79 |
| HbA1c | GLP1RA | Inverse variance weighted | 4 | −0.007 | (−0.034, 0.020) | 0.63 | | | 0.42 | 0.94 | 0 | 0.97 | 23.04 |
| HbA1c | AMPK | Inverse variance weighted | 3 | 0.024 | (−0.036, 0.085) | 0.43 | | | 4.78 | 0.09 | 0 | 0.49 | 14.60 |
| HbA1c | MG3 | Wald ratio | 1 | −0.015 | (−0.086, 0.057) | 0.69 | | | | | | | 13.38 |
| HbA1c | GCG | Wald ratio | 1 | −0.095 | (−0.225, 0.035) | 0.15 | | | | | | | 4.12 |
| HbA1c | GDF15 | Wald ratio | 1 | 0.100 | (0.029, 0.172) | 0.01 | | | | | | | 13.34 |
| LDL | Lowering LDL | Inverse variance weighted | 228 | −0.007 | (−0.011, −0.003) | 0.00 | | | 1650.25 | 0 | 0 | 0.9 | 241.48 |
| LDL | PCSK9I | Inverse variance weighted | 11 | 0.001 | (−0.008, 0.009) | 0.90 | | | 51.3 | 0 | 0 | 0.01 | 869.08 |
| LDL | NPC1L1I | Inverse variance weighted | 6 | −0.020 | (−0.033, −0.008) | 0.00 | | | 6.95 | 0.22 | 0 | 0.42 | 191.47 |
| LDL | Statin | Inverse variance weighted | 5 | −0.013 | (−0.019, −0.007) | 0.00 | | | 1.07 | 0.9 | 0 | 0.59 | 787.71 |
| SBP | Lowering SBP | Inverse variance weighted | 441 | 0.001 | (−0.003, 0.005) | 0.67 | | | 5342.3 | 0 | 0 | 0.42 | 75.13 |
| SBP | CCB | Inverse variance weighted | 24 | 0.006 | (0.001, 0.011) | 0.02 | | | 23.4 | 0.44 | 0 | 0.9 | 66.21 |
| SBP | BB | Inverse variance weighted | 5 | −0.011 | (−0.030, 0.008) | 0.24 | | | 9.22 | 0.06 | 0 | 0.98 | 52.28 |
| SBP | ACEI | Wald ratio | 1 | 0.018 | (−0.003, 0.040) | 0.10 | | | | | | | 82.80 |

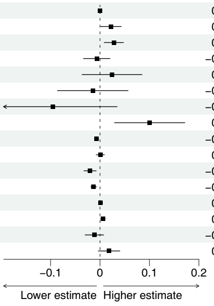


**eGFR cross-sectional cystatin (Stanzick, et.al. (2021))**

| IntermediatePheno | exposure | method | nsnp | Beta | 95%CI | pval | RSSobs | Phet | Q | Q_pval | egger_intercept | egger_pval | MeanFstat |
|---|---|---|---|---|---|---|---|---|---|---|---|---|---|
| HbA1c | Lowering HbA1c | Inverse variance weighted | 273 | −0.003 | (−0.008, 0.002) | 0.19 | | | 1270.6 | 0 | 0 | 0.34 | 124.76 |
| HbA1c | MC1 | Inverse variance weighted | 24 | 0.023 | (−0.007, 0.053) | 0.14 | | | 90.68 | 0 | 0 | 0.36 | 31.06 |
| HbA1c | SGLT2I | Inverse variance weighted | 5 | 0.010 | (−0.025, 0.046) | 0.57 | | | 0.46 | 0.98 | 0 | 0.95 | 30.16 |
| HbA1c | GLP1RA | Inverse variance weighted | 4 | −0.022 | (−0.067, 0.024) | 0.35 | | | 2.42 | 0.49 | 0 | 0.38 | 23.04 |
| HbA1c | AMPK | Inverse variance weighted | 3 | 0.052 | (−0.016, 0.119) | 0.13 | | | 1.22 | 0.54 | 0 | 0.54 | 14.60 |
| HbA1c | MG3 | Wald ratio | 1 | 0.083 | (−0.027, 0.193) | 0.14 | | | | | | | 13.38 |
| HbA1c | GCG | Wald ratio | 1 | 0.080 | (−0.125, 0.284) | 0.45 | | | | | | | 4.12 |
| HbA1c | GDF15 | Wald ratio | 1 | 0.103 | (−0.016, 0.222) | 0.09 | | | | | | | 13.34 |
| LDL | Lowering LDL | Inverse variance weighted | 221 | −0.006 | (−0.013, 0.001) | 0.07 | | | 1481.24 | 0 | 0 | 0.32 | 242.87 |
| LDL | PCSK9I | Inverse variance weighted | 11 | 0.002 | (−0.005, 0.008) | 0.63 | | | 9.34 | 0.5 | 0 | 0.09 | 869.08 |
| LDL | NPC1L1I | Inverse variance weighted | 6 | 0.008 | (−0.010, 0.025) | 0.41 | | | 0.3 | 1 | 0 | 0.99 | 191.47 |
| LDL | Statin | Inverse variance weighted | 5 | −0.013 | (−0.023, −0.004) | 0.01 | | | 2.71 | 0.61 | 0 | 0.54 | 787.71 |
| SBP | Lowering SBP | Inverse variance weighted | 439 | 0.001 | (−0.004, 0.006) | 0.70 | | | 2925.73 | 0 | 0 | 0.92 | 74.78 |
| SBP | CCB | Inverse variance weighted | 24 | 0.001 | (−0.008, 0.010) | 0.82 | | | 28.03 | 0.21 | 0 | 0.71 | 66.21 |
| SBP | BB | Inverse variance weighted | 5 | −0.018 | (−0.044, 0.009) | 0.19 | | | 6.78 | 0.15 | 0 | 0.94 | 52.28 |
| SBP | ACEI | Wald ratio | 1 | 0.032 | (−0.006, 0.069) | 0.10 | | | | | | | 82.80 |

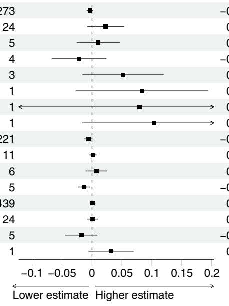


**BUN (Stanzick, et.al. (2021))**

| IntermediatePheno | exposure | method | nsnp | Beta | 95%CI | pval | RSSobs | Phet | Q | Q_pval | egger_intercept | egger_pval | MeanFstat |
|---|---|---|---|---|---|---|---|---|---|---|---|---|---|
| HbA1c | Lowering HbA1c | Inverse variance weighted | 274 | −0.002 | (−0.007, 0.004) | 0.53 | | | 1356.61 | 0 | 0 | 0.09 | 124.42 |
| HbA1c | MC1 | Inverse variance weighted | 24 | −0.011 | (−0.032, 0.010) | 0.30 | | | 34.13 | 0.06 | 0 | 0.38 | 31.06 |
| HbA1c | SGLT2I | Inverse variance weighted | 6 | −0.018 | (−0.055, 0.019) | 0.33 | | | 5.96 | 0.31 | 0 | 0.86 | 29.79 |
| HbA1c | GLP1RA | Inverse variance weighted | 4 | −0.079 | (−0.162, 0.004) | 0.06 | | | 9.31 | 0.03 | 0 | 0.1 | 23.04 |
| HbA1c | AMPK | Inverse variance weighted | 3 | 0.061 | (−0.098, 0.221) | 0.45 | | | 11.37 | 0 | 0 | 0.42 | 14.60 |
| HbA1c | MG3 | Wald ratio | 1 | −0.125 | (−0.258, 0.009) | 0.07 | | | | | | | 13.38 |
| HbA1c | GCG | Wald ratio | 1 | 0.046 | (−0.196, 0.288) | 0.71 | | | | | | | 4.12 |
| HbA1c | GDF15 | Wald ratio | 1 | 0.009 | (−0.111, 0.130) | 0.88 | | | | | | | 13.34 |
| LDL | Lowering LDL | Inverse variance weighted | 220 | 0.001 | (−0.005, 0.008) | 0.67 | | | 1242.92 | 0 | 0 | 0.6 | 241.13 |
| LDL | PCSK9I | Inverse variance weighted | 11 | −0.004 | (−0.014, 0.005) | 0.37 | | | 20.12 | 0.03 | 0 | 0.04 | 869.08 |
| LDL | NPC1L1I | Inverse variance weighted | 6 | 0.009 | (−0.010, 0.027) | 0.35 | | | 4.1 | 0.53 | 0 | 0.28 | 191.47 |
| LDL | Statin | Inverse variance weighted | 5 | 0.010 | (−0.000, 0.020) | 0.06 | | | 1.14 | 0.89 | 0 | 0.81 | 787.71 |
| SBP | Lowering SBP | Inverse variance weighted | 436 | −0.008 | (−0.013, −0.003) | 0.00 | | | 2916.01 | 0 | 0 | 0.11 | 75.13 |
| SBP | CCB | Inverse variance weighted | 24 | 0.004 | (−0.005, 0.013) | 0.35 | | | 16.16 | 0.85 | 0 | 0.66 | 66.21 |
| SBP | BB | Inverse variance weighted | 5 | −0.020 | (−0.042, 0.002) | 0.07 | | | 4.18 | 0.38 | 0 | 0.98 | 52.28 |
| SBP | ACEI | Wald ratio | 1 | −0.025 | (−0.062, 0.013) | 0.20 | | | | | | | 82.80 |

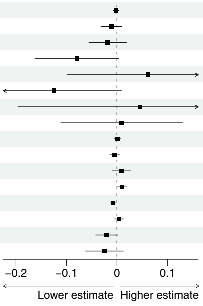

Figure S4. Scatter plots of exposures and outcome eGFR change.

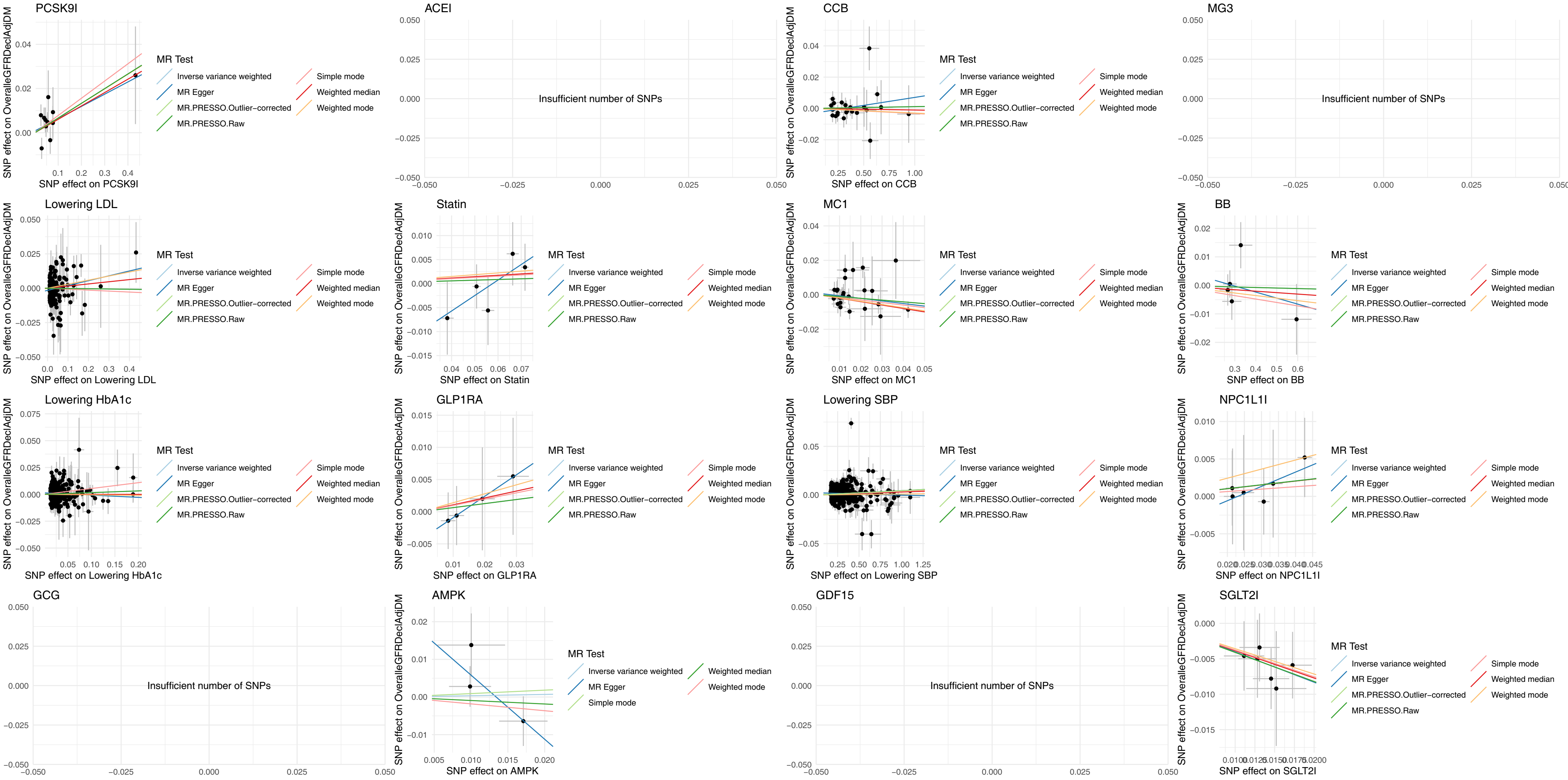

### Figure S5. Scatter plots of exposures and outcome cross-sectional eGFR-creat in all ancestry.

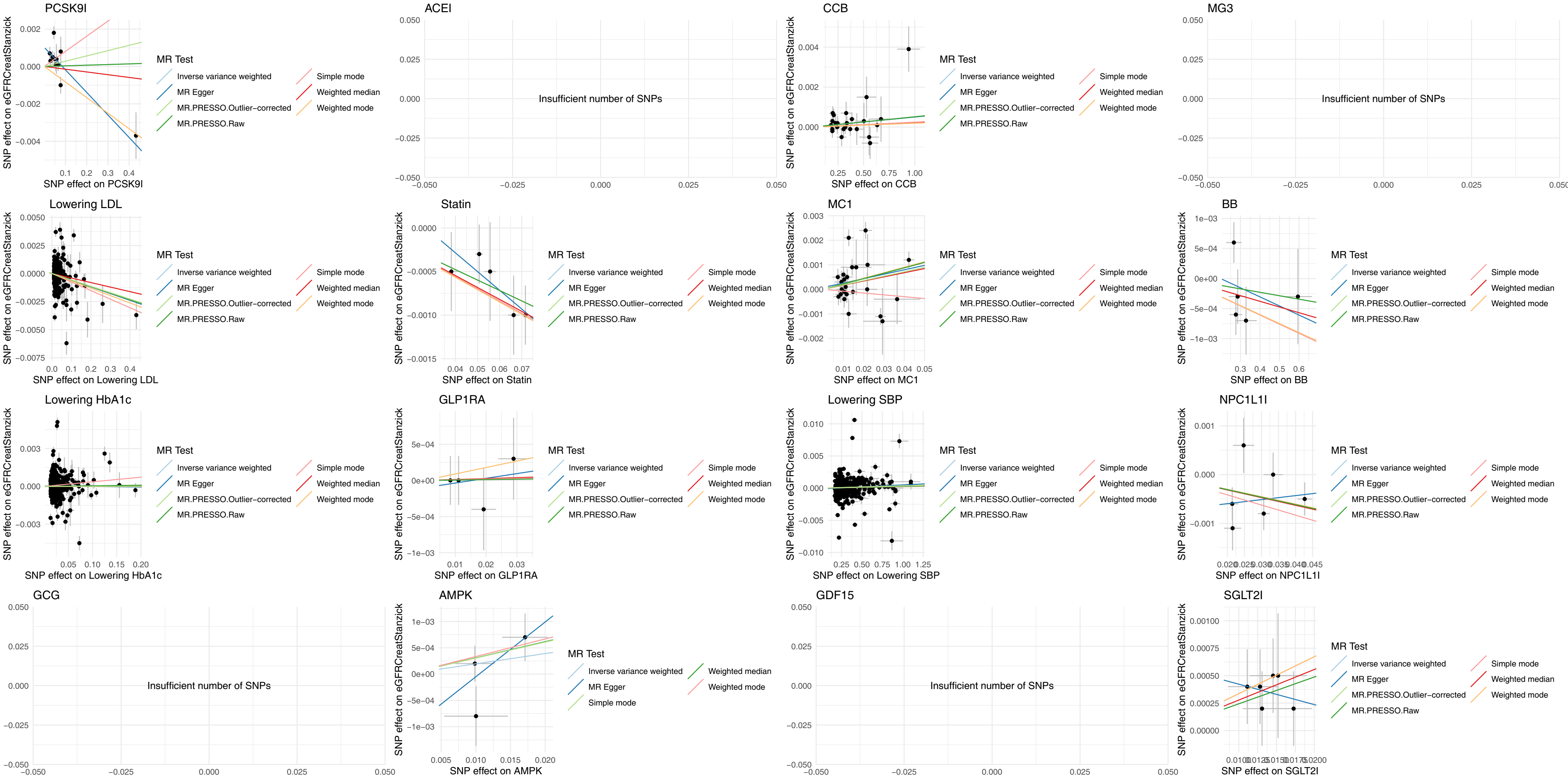

Figure S6. Scatter plots of exposures and outcome cross-sectional eGFR-creat in European only.

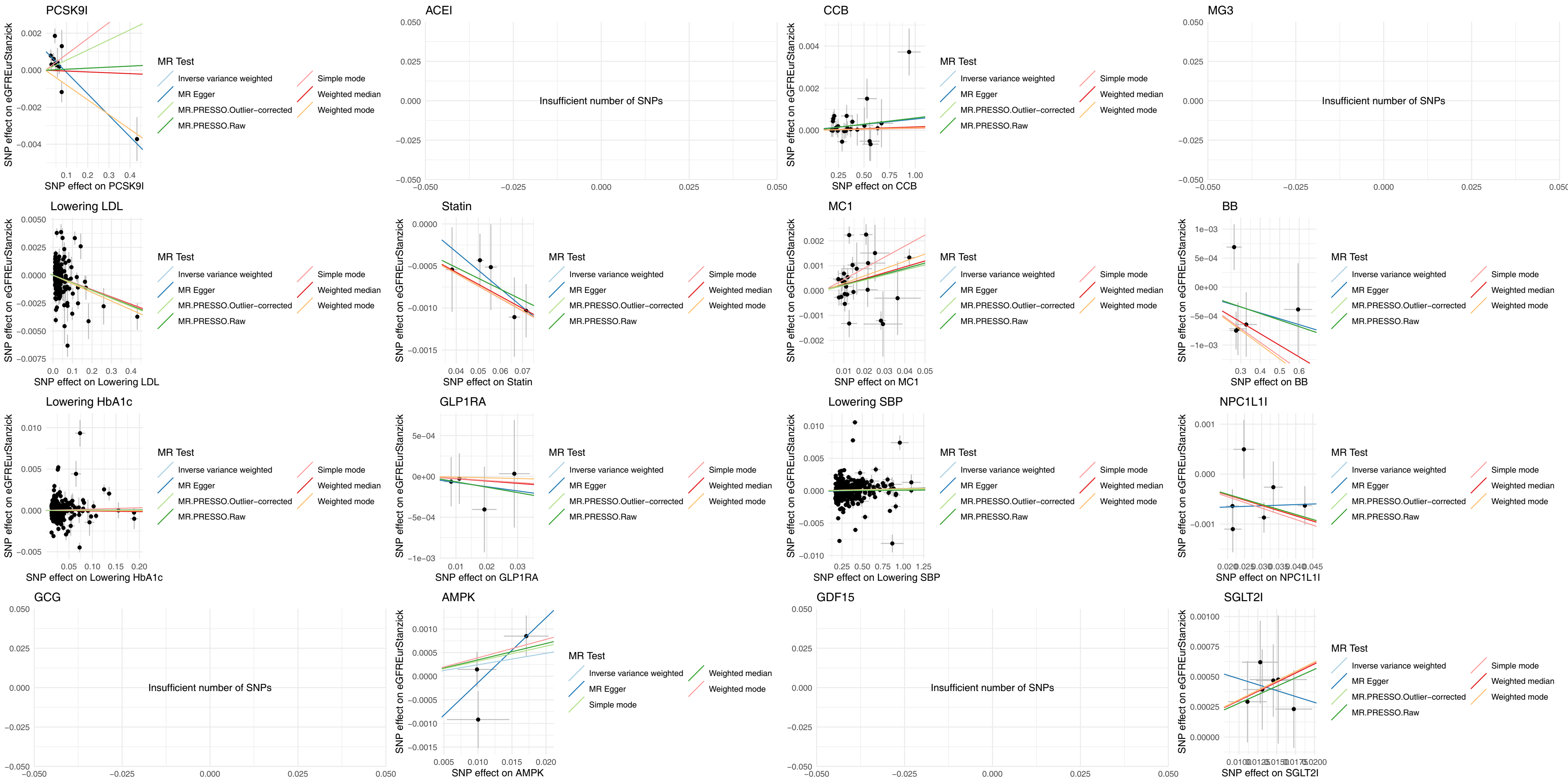

Figure S7. Scatter plots of exposures and outcome cross-sectional eGFR-cys (European only).

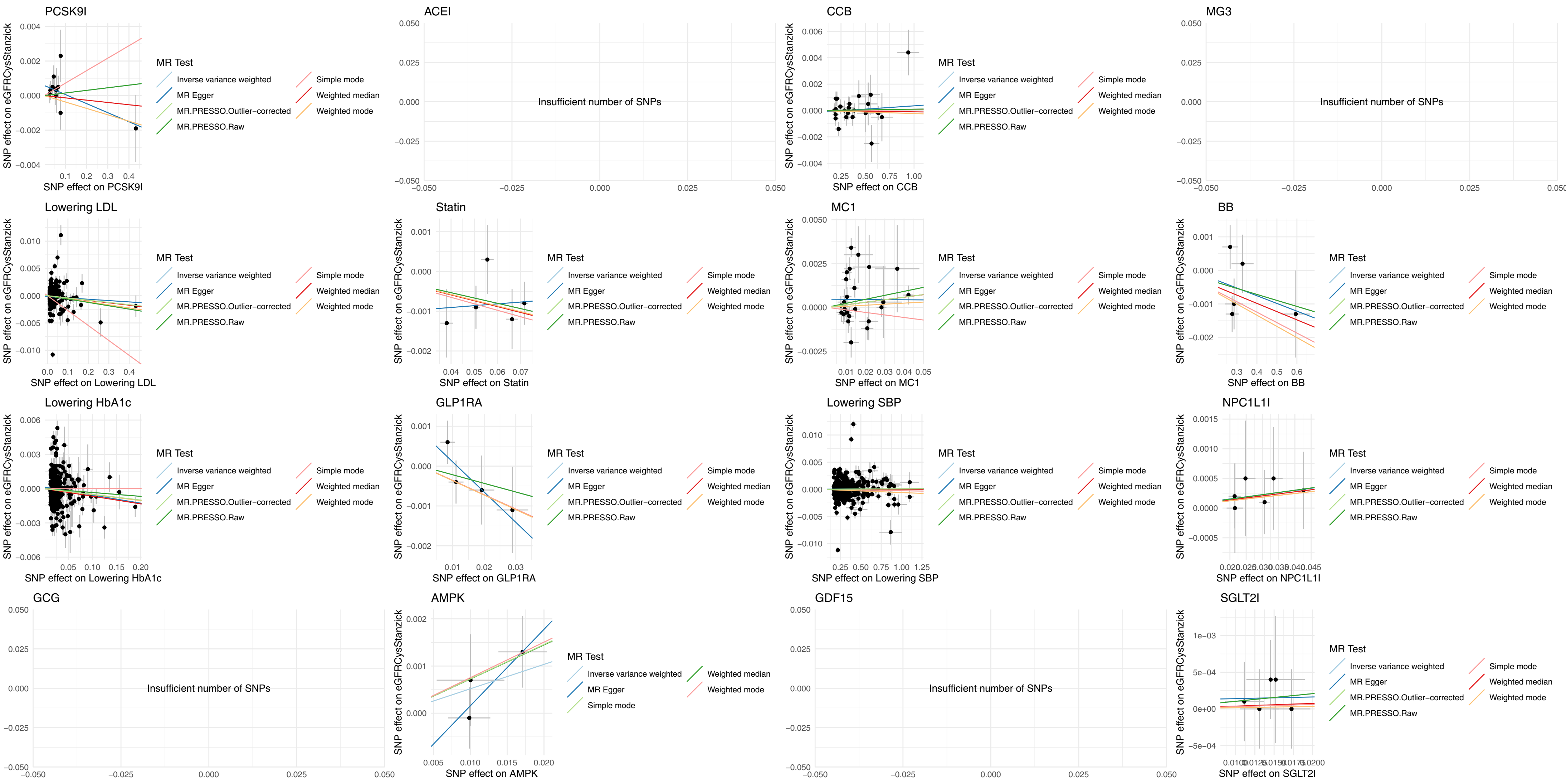

Figure S8. Scatter plots of exposures and outcome cross-sectional BUN (European only).

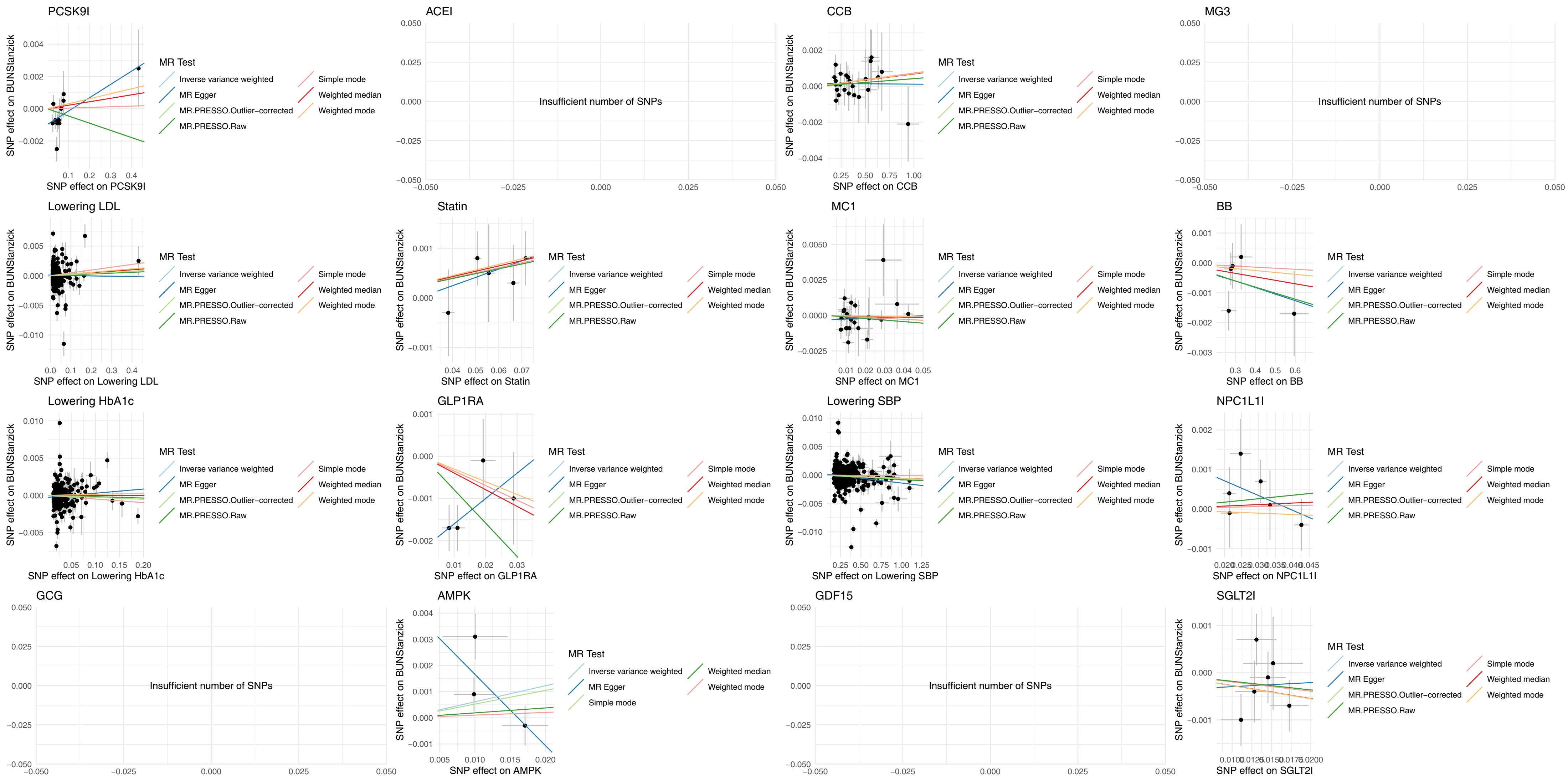

### Table S1. Genetic variants as instrument variables for exposure of SBP, LDL, and HbA1c

| SNP | gene.exposure | chr.exposure | pos.exposure | effect_allele.exposure | other_allele.exposure | eaf.exposure | beta.exposure | se.exposure | pval.exposure | samplesize.exposure | units.exposure | exposure | mr_keep.exposure | pval_origin.exposure | units.exposure_dat | id.exposure | data_source.exposure | IntermediatePheno |
|---|---|---|---|---|---|---|---|---|---|---|---|---|---|---|---|---|---|---|
| rs4291 | ACE | 17 | 61554194 | T | A | 0.3845 | 0.2839 | 0.0312 | 8.65E-20 | 725185 | units | ACEI | TRUE | reported | units | D94SpL | textfile | SBP |
| rs11196549 | ADRB1 | 10 | 115723772 | A | G | 0.0475 | 0.5944 | 0.0721 | 1.70E-16 | 712636 | units | BB | TRUE | reported | units | IBfVDL | textfile | SBP |
| rs460718 | ADRB1 | 10 | 115721364 | G | A | 0.6734 | 0.2764 | 0.0324 | 1.36E-17 | 725026 | units | BB | TRUE | reported | units | IBfVDL | textfile | SBP |
| rs11196597 | ADRB1 | 10 | 115788094 | A | G | 0.133 | 0.2858 | 0.0458 | 4.23E-10 | 701272 | units | BB | TRUE | reported | units | IBfVDL | textfile | SBP |
| rs17875473 | ADRB1 | 10 | 115800294 | T | C | 0.0871 | 0.3283 | 0.0552 | 2.66E-09 | 707302 | units | BB | TRUE | reported | units | IBfVDL | textfile | SBP |
| rs4359161 | ADRB1 | 10 | 115826508 | G | A | 0.8188 | 0.2662 | 0.0391 | 9.46E-12 | 733998 | units | BB | TRUE | reported | units | IBfVDL | textfile | SBP |
| rs3821843 | CACNA1D | 3 | 53558012 | A | G | 0.6808 | 0.3373 | 0.0335 | 6.56E-24 | 684820 | units | CCB | TRUE | reported | units | f3yEFg | textfile | SBP |
| rs114987861 | CACNA1D | 3 | 53605712 | A | G | 0.0284 | 0.5289 | 0.0958 | 3.36E-08 | 680765 | units | CCB | TRUE | reported | units | f3yEFg | textfile | SBP |
| rs113210396 | CACNA1D | 3 | 53612327 | G | T | 0.9549 | 0.4338 | 0.077 | 1.76E-08 | 664156 | units | CCB | TRUE | reported | units | f3yEFg | textfile | SBP |
| rs7340705 | CACNA1D | 3 | 53734443 | C | T | 0.3268 | 0.2425 | 0.0322 | 4.87E-14 | 727632 | units | CCB | TRUE | reported | units | f3yEFg | textfile | SBP |
| rs2488136 | CACNB2 | 10 | 18334521 | A | G | 0.2875 | 0.2261 | 0.0334 | 1.22E-11 | 730374 | units | CCB | TRUE | reported | units | f3yEFg | textfile | SBP |
| rs1888693 | CACNB2 | 10 | 18440444 | A | G | 0.3449 | 0.3858 | 0.0317 | 4.69E-34 | 732344 | units | CCB | TRUE | reported | units | f3yEFg | textfile | SBP |
| rs16916914 | CACNB2 | 10 | 18457722 | C | T | 0.0369 | 0.5636 | 0.0806 | 2.72E-12 | 731283 | units | CCB | TRUE | reported | units | f3yEFg | textfile | SBP |
| rs7076319 | CACNB2 | 10 | 18459450 | G | A | 0.2661 | 0.321 | 0.0341 | 5.07E-21 | 732191 | units | CCB | TRUE | reported | units | f3yEFg | textfile | SBP |
| rs61278674 | CACNB2 | 10 | 18481737 | G | A | 0.0938 | 0.3298 | 0.054 | 1.03E-09 | 676191 | units | CCB | TRUE | reported | units | f3yEFg | textfile | SBP |
| rs1779209 | CACNB2 | 10 | 18516925 | T | C | 0.3145 | 0.2453 | 0.0325 | 4.29E-14 | 730147 | units | CCB | TRUE | reported | units | f3yEFg | textfile | SBP |
| rs10828399 | CACNB2 | 10 | 18553968 | G | A | 0.4782 | 0.1947 | 0.0302 | 1.10E-10 | 732842 | units | CCB | TRUE | reported | units | f3yEFg | textfile | SBP |
| rs10828452 | CACNB2 | 10 | 18592450 | A | T | 0.793 | 0.3046 | 0.0388 | 4.20E-15 | 679250 | units | CCB | TRUE | reported | units | f3yEFg | textfile | SBP |
| rs10828542 | CACNB2 | 10 | 18627285 | A | G | 0.6137 | 0.1817 | 0.0311 | 5.18E-09 | 725325 | units | CCB | TRUE | reported | units | f3yEFg | textfile | SBP |
| rs12780039 | CACNB2 | 10 | 18678987 | C | G | 0.121 | 0.2852 | 0.047 | 1.26E-09 | 712266 | units | CCB | TRUE | reported | units | f3yEFg | textfile | SBP |
| rs112133583 | CACNB2 | 10 | 18695681 | C | T | 0.9701 | 0.5546 | 0.0973 | 1.18E-08 | 625088 | units | CCB | TRUE | reported | units | f3yEFg | textfile | SBP |
| rs11014170 | CACNB2 | 10 | 18710991 | G | A | 0.9794 | 0.6701 | 0.115 | 5.61E-09 | 647186 | units | CCB | TRUE | reported | units | f3yEFg | textfile | SBP |
| rs7923191 | CACNB2 | 10 | 18727901 | G | A | 0.2082 | 0.369 | 0.0376 | 1.09E-22 | 713691 | units | CCB | TRUE | reported | units | f3yEFg | textfile | SBP |
| rs12258967 | CACNB2 | 10 | 18727959 | C | G | 0.7047 | 0.6327 | 0.0337 | 1.08E-78 | 711222 | units | CCB | TRUE | reported | units | f3yEFg | textfile | SBP |
| rs72786098 | CACNB2 | 10 | 18729855 | G | A | 0.9678 | 0.5033 | 0.0883 | 1.18E-08 | 705121 | units | CCB | TRUE | reported | units | f3yEFg | textfile | SBP |
| rs1998822 | CACNB2 | 10 | 18755664 | G | A | 0.2766 | 0.1958 | 0.0343 | 1.15E-08 | 707468 | units | CCB | TRUE | reported | units | f3yEFg | textfile | SBP |
| rs4748474 | CACNB2 | 10 | 18790727 | A | G | 0.5214 | 0.1946 | 0.0304 | 1.61E-10 | 723052 | units | CCB | TRUE | reported | units | f3yEFg | textfile | SBP |
| rs150857355 | CACNB3 | 12 | 49209340 | C | G | 0.0217 | 0.9406 | 0.1122 | 5.20E-17 | 650268 | units | CCB | TRUE | reported | units | f3yEFg | textfile | SBP |
| rs2239046 | CACNA1C | 12 | 2434419 | A | G | 0.6817 | 0.2082 | 0.0322 | 9.58E-11 | 739131 | units | CCB | TRUE | reported | units | f3yEFg | textfile | SBP |
| rs714277 | CACNA1C | 12 | 2514270 | T | C | 0.2834 | 0.1986 | 0.0333 | 2.38E-09 | 738689 | units | CCB | TRUE | reported | units | f3yEFg | textfile | SBP |
| rs217386 | NPC1L1 | 7 | 44600695 | G | A | 0.581 | 0.0306736 | 0.00169123 | 1.53E-57 | 828800 | units | NPC1L1I | TRUE | reported | units | Sr1HqQ | textfile | LDL |
| rs2073547 | NPC1L1 | 7 | 44582331 | G | A | 0.202 | 0.0426732 | 0.0021095 | 3.92E-71 | 810540 | units | NPC1L1I | TRUE | reported | units | Sr1HqQ | textfile | LDL |
| rs17655652 | NPC1L1 | 7 | 44580991 | T | C | 0.691 | 0.0214347 | 0.00186978 | 5.20E-24 | 797660 | units | NPC1L1I | TRUE | reported | units | Sr1HqQ | textfile | LDL |
| rs7791240 | NPC1L1 | 7 | 44602589 | C | T | 0.0976 | 0.033426 | 0.00278528 | 3.71E-26 | 841736 | units | NPC1L1I | TRUE | reported | units | Sr1HqQ | textfile | LDL |
| rs10234070 | NPC1L1 | 7 | 44537696 | T | C | 0.122 | 0.0215346 | 0.00255523 | 1.09E-13 | 841647 | units | NPC1L1I | TRUE | reported | units | Sr1HqQ | textfile | LDL |
| rs2300414 | NPC1L1 | 7 | 44682938 | A | G | 0.0765 | 0.0247834 | 0.00309308 | 1.62E-12 | 841814 | units | NPC1L1I | TRUE | reported | units | Sr1HqQ | textfile | LDL |
| rs2479394 | PCSK9 | 1 | 55486064 | G | A | 0.282 | 0.0288483 | 0.00180404 | 3.97E-45 | 838176 | units | PCSK9I | TRUE | reported | units | 9rRHy9 | textfile | LDL |
| rs11206510 | PCSK9 | 1 | 55496039 | T | C | 0.821 | 0.06576 | 0.0021498 | 3.72E-160 | 815718 | units | PCSK9I | TRUE | reported | units | 9rRHy9 | textfile | LDL |
| rs2479409 | PCSK9 | 1 | 55504650 | G | A | 0.338 | 0.0475249 | 0.00176457 | 1.30E-124 | 805217 | units | PCSK9I | TRUE | reported | units | 9rRHy9 | textfile | LDL |
| rs11591147 | PCSK9 | 1 | 55505647 | G | T | 0.9843 | 0.432401 | 0.00647577 | 1.00E-200 | 835772 | units | PCSK9I | TRUE | reported | units | 9rRHy9 | textfile | LDL |
| rs572512 | PCSK9 | 1 | 55517344 | T | C | 0.345 | 0.0257849 | 0.00187785 | 9.97E-34 | 763132 | units | PCSK9I | TRUE | reported | units | 9rRHy9 | textfile | LDL |
| rs585131 | PCSK9 | 1 | 55524116 | T | C | 0.827 | 0.0449434 | 0.00215947 | 3.48E-75 | 839963 | units | PCSK9I | TRUE | reported | units | 9rRHy9 | textfile | LDL |
| rs12067569 | PCSK9 | 1 | 55528629 | A | G | 0.0341 | 0.0775073 | 0.00458236 | 2.80E-50 | 842606 | units | PCSK9I | TRUE | reported | units | 9rRHy9 | textfile | LDL |
| rs10493176 | PCSK9 | 1 | 55538552 | T | G | 0.9092 | 0.0770077 | 0.00291663 | 7.82E-120 | 842612 | units | PCSK9I | TRUE | reported | units | 9rRHy9 | textfile | LDL |
| rs11583974 | PCSK9 | 1 | 55551718 | A | G | 0.0381 | 0.057305 | 0.0045749 | 2.39E-28 | 836859 | units | PCSK9I | TRUE | reported | units | 9rRHy9 | textfile | LDL |
| rs2495477 | PCSK9 | 1 | 55518467 | A | G | 0.601 | 0.0540576 | 0.00178118 | 1.09E-157 | 789095 | units | PCSK9I | TRUE | reported | units | 9rRHy9 | textfile | LDL |
| rs10888897 | PCSK9 | 1 | 55513061 | C | T | 0.626 | 0.039283 | 0.00175374 | 8.61E-87 | 790721 | units | PCSK9I | TRUE | reported | units | 9rRHy9 | textfile | LDL |
| rs10515198 | HMGCR | 5 | 74641560 | A | G | 0.0989 | 0.0556756 | 0.00272222 | 1.14E-72 | 842630 | units | Statin | TRUE | reported | units | fwCnU5 | textfile | LDL |
| rs12916 | HMGCR | 5 | 74656539 | C | T | 0.403 | 0.0716379 | 0.00166597 | 1.00E-200 | 842634 | units | Statin | TRUE | reported | units | fwCnU5 | textfile | LDL |
| rs12173076 | HMGCR | 5 | 74697050 | G | T | 0.115 | 0.0663316 | 0.0026113 | 4.60E-111 | 842631 | units | Statin | TRUE | reported | units | fwCnU5 | textfile | LDL |
| rs2303152 | HMGCR | 5 | 74641707 | A | G | 0.0958 | 0.0381259 | 0.00276126 | 4.38E-34 | 842621 | units | Statin | TRUE | reported | units | fwCnU5 | textfile | LDL |
| rs10066707 | HMGCR | 5 | 74560579 | A | G | 0.374 | 0.0507496 | 0.00175593 | 3.41E-143 | 838160 | units | Statin | TRUE | reported | units | fwCnU5 | textfile | LDL |
| rs1645060 | PRKAA1 | 5 | 41391349 | A | G | 0.067139 | 0.010031 | 0.0045945 | 0.0290128 | 344182 | units | AMPK | TRUE | reported | units | Vyjxei | textfile | HbA1c |
| rs3793342 | PRKAG2 | 7 | 150695195 | G | A | 0.8568 | 0.017102 | 0.003286 | 1.95E-07 | 344182 | units | AMPK | TRUE | reported | units | Vyjxei | textfile | HbA1c |
| rs10272655 | PRKAG2 | 7 | 151643303 | T | C | 0.20815 | 0.0098639 | 0.002855 | 5.50E-04 | 344182 | units | AMPK | TRUE | reported | units | Vyjxei | textfile | HbA1c |
| rs72866989 | GCG | 2 | 162382170 | G | A | 0.9903772 | 0.023898 | 0.011768 | 0.0422854 | 344182 | units | GCG | TRUE | reported | units | vxLdAw | textfile | HbA1c |
| rs1227732 | GDF15 | 19 | 18498808 | T | G | 0.19168 | 0.010667 | 0.0029207 | 2.60E-04 | 344182 | units | GDF15 | TRUE | reported | units | WbqWq0 | textfile | HbA1c |
| rs10305420 | GLP1R | 6 | 39016636 | C | T | 0.60655 | 0.011086 | 0.0023721 | 2.96E-06 | 344182 | units | GLP1RA | TRUE | reported | units | puxRXK | textfile | HbA1c |
| rs75151020 | GLP1R | 6 | 39031592 | C | A | 0.092777 | 0.01922 | 0.0039679 | 1.27E-06 | 344182 | units | GLP1RA | TRUE | reported | units | puxRXK | textfile | HbA1c |
| rs2268647 | GLP1R | 6 | 39043178 | T | C | 0.52106 | 0.0084163 | 0.0023019 | 2.56E-04 | 344182 | units | GLP1RA | TRUE | reported | units | puxRXK | textfile | HbA1c |
| rs10305518 | GLP1R | 6 | 39055012 | G | T | 0.056804 | 0.028924 | 0.0049979 | 7.16E-09 | 344182 | units | GLP1RA | TRUE | reported | units | puxRXK | textfile | HbA1c |
| rs117877390 | NDUFA13 | 19 | 19378416 | T | C | 0.027086 | 0.02535 | 0.0073311 | 5.44E-04 | 344182 | units | MC1 | TRUE | reported | units | IaocOm | textfile | HbA1c |
| rs2965201 | NDUFA13 | 19 | 19478051 | T | C | 0.83205 | 0.011113 | 0.0030787 | 3.07E-04 | 344182 | units | MC1 | TRUE | reported | units | IaocOm | textfile | HbA1c |

### Table S1. Genetic variants as instrument variables for exposure of SBP, LDL, and HbA1c (continue)

| SNP | gene.exposure | chr.exposure | pos.exposure | effect_allele.exposure | other_allele.exposure | eaf.exposure | beta.exposure | se.exposure | pval.exposure | samplesize.exposure | units.exposure | exposure | mr_keep.exposure | pval_origin.exposure | units.exposure_dat | id.exposure | data_source.exposure | IntermediatePheno |
|---|---|---|---|---|---|---|---|---|---|---|---|---|---|---|---|---|---|---|
| rs62383878 | NDUFA2 | 5 | 139215386 | A | C | 0.76731 | 0.0087409 | 0.0027295 | 0.00136311 | 344182 | units | MC1 | TRUE | reported | units | IaocOm | textfile | HbA1c |
| rs6897346 | NDUFA2 | 5 | 139694387 | C | T | 0.8028 | 0.009195 | 0.0028948 | 0.00149149 | 344182 | units | MC1 | TRUE | reported | units | IaocOm | textfile | HbA1c |
| rs7788702 | NDUFA5 | 7 | 123237421 | A | G | 0.29947 | 0.014302 | 0.0025121 | 1.25E-08 | 344182 | units | MC1 | TRUE | reported | units | IaocOm | textfile | HbA1c |
| rs1043409 | NDUFA7 | 19 | 7536141 | T | A | 0.059431 | 0.021729 | 0.004935 | 1.07E-05 | 344182 | units | MC1 | TRUE | reported | units | IaocOm | textfile | HbA1c |
| rs73497430 | NDUFA7 | 19 | 8420054 | T | G | 0.75489 | 0.010426 | 0.0026894 | 1.06E-04 | 344182 | units | MC1 | TRUE | reported | units | IaocOm | textfile | HbA1c |
| rs4837917 | NDUFA8 | 9 | 124709309 | C | T | 0.29452 | 0.0075748 | 0.0025201 | 0.00264978 | 344182 | units | MC1 | TRUE | reported | units | IaocOm | textfile | HbA1c |
| rs150943293 | NDUFA8 | 9 | 125453400 | A | G | 0.981514 | 0.021926 | 0.0085707 | 0.0105201 | 344182 | units | MC1 | TRUE | reported | units | IaocOm | textfile | HbA1c |
| rs8027626 | NDUFAF1 | 15 | 41711514 | G | T | 0.32388 | 0.012638 | 0.0024553 | 2.65E-07 | 344182 | units | MC1 | TRUE | reported | units | IaocOm | textfile | HbA1c |
| rs9866749 | NDUFAF3 | 3 | 49650935 | T | A | 0.70539 | 0.028301 | 0.0025857 | 7.08E-28 | 344182 | units | MC1 | TRUE | reported | units | IaocOm | textfile | HbA1c |
| rs1354034 | NDUFAF3 | 3 | 56849749 | C | T | 0.60126 | 0.014722 | 0.0023452 | 3.44E-10 | 344182 | units | MC1 | TRUE | reported | units | IaocOm | textfile | HbA1c |
| rs9399137 | NDUFAF3 | 6 | 135419018 | T | C | 0.74104 | 0.042268 | 0.0026336 | 6.04E-58 | 344182 | units | MC1 | TRUE | reported | units | IaocOm | textfile | HbA1c |
| rs147052086 | NDUFAF3 | 3 | 48412551 | A | G | 0.015943 | 0.029285 | 0.0096112 | 0.00231132 | 344182 | units | MC1 | TRUE | reported | units | IaocOm | textfile | HbA1c |
| rs62180557 | NDUFB3 | 2 | 200986595 | T | C | 0.97631 | 0.016336 | 0.0075672 | 0.0308653 | 344182 | units | MC1 | TRUE | reported | units | IaocOm | textfile | HbA1c |
| rs653790 | NDUFB6 | 9 | 32576997 | C | T | 0.75418 | 0.011865 | 0.002807 | 2.37E-05 | 344182 | units | MC1 | TRUE | reported | units | IaocOm | textfile | HbA1c |
| rs792699 | NDUFB8 | 10 | 102942689 | G | C | 0.75305 | 0.0072714 | 0.0026915 | 0.00690129 | 344182 | units | MC1 | TRUE | reported | units | IaocOm | textfile | HbA1c |
| rs77145138 | NDUFC1 | 4 | 139765732 | C | A | 0.010607 | 0.036483 | 0.01146 | 0.00145459 | 344182 | units | MC1 | TRUE | reported | units | IaocOm | textfile | HbA1c |
| rs2450122 | NDUFC2 | 11 | 77930345 | T | C | 0.84513 | 0.020952 | 0.0031757 | 4.19E-11 | 344182 | units | MC1 | TRUE | reported | units | IaocOm | textfile | HbA1c |
| rs4657093 | NDUFS2 | 1 | 161693003 | T | C | 0.87109 | 0.0090254 | 0.0034409 | 0.00871746 | 344182 | units | MC1 | TRUE | reported | units | IaocOm | textfile | HbA1c |
| rs1809084 | NDUFS4 | 5 | 52025487 | T | C | 0.30621 | 0.011632 | 0.0024884 | 2.95E-06 | 344182 | units | MC1 | TRUE | reported | units | IaocOm | textfile | HbA1c |
| rs62372178 | NDUFS4 | 5 | 53503340 | T | C | 0.56501 | 0.010461 | 0.002329 | 7.08E-06 | 344182 | units | MC1 | TRUE | reported | units | IaocOm | textfile | HbA1c |
| rs151128822 | NDUFS8 | 11 | 67440592 | G | A | 0.90734 | 0.012613 | 0.0039633 | 0.00146009 | 344182 | units | MC1 | TRUE | reported | units | IaocOm | textfile | HbA1c |
| rs1532331 | NDUFV1 | 5 | 43116830 | T | G | 0.69674 | 0.010369 | 0.0025112 | 3.64E-05 | 344182 | units | MC1 | TRUE | reported | units | IaocOm | textfile | HbA1c |
| rs12969399 | NDUFV2 | 18 | 9091853 | G | T | 0.3602 | 0.010038 | 0.0023951 | 2.78E-05 | 344182 | units | MC1 | TRUE | reported | units | IaocOm | textfile | HbA1c |
| rs11889246 | GPD2 | 2 | 157468544 | C | A | 0.74323 | 0.0096368 | 0.0026346 | 2.55E-04 | 344182 | units | MG3 | TRUE | reported | units | fhrnCj | textfile | HbA1c |
| rs4488457 | SLC5A2 | 16 | 31659189 | T | G | 0.28797 | 0.013084 | 0.0025505 | 2.90E-07 | 344182 | units | SGLT2I | TRUE | reported | units | Xjf1jJ | textfile | HbA1c |
| rs11865835 | SLC5A2 | 16 | 31509816 | T | C | 0.71642 | 0.011123 | 0.0025546 | 1.34E-05 | 344182 | units | SGLT2I | TRUE | reported | units | Xjf1jJ | textfile | HbA1c |
| rs35445454 | SLC5A2 | 16 | 31699326 | C | T | 0.65563 | 0.012817 | 0.0024242 | 1.24E-07 | 344182 | units | SGLT2I | TRUE | reported | units | Xjf1jJ | textfile | HbA1c |
| rs111510548 | SLC5A2 | 16 | 31476695 | T | C | 0.8966 | 0.015206 | 0.0038138 | 6.69E-05 | 344182 | units | SGLT2I | TRUE | reported | units | Xjf1jJ | textfile | HbA1c |
| rs2070896 | SLC5A2 | 16 | 31384554 | T | C | 0.62503 | 0.017269 | 0.0024469 | 1.70E-12 | 344182 | units | SGLT2I | TRUE | reported | units | Xjf1jJ | textfile | HbA1c |
| rs28692853 | SLC5A2 | 16 | 31573030 | C | A | 0.49258 | 0.014549 | 0.0023054 | 2.78E-10 | 344182 | units | SGLT2I | TRUE | reported | units | Xjf1jJ | textfile | HbA1c |

### Table S2. The effect of genetically proxied antihypertensives, lipid-lowering, and antidiabetic drugs on annual eGFR change

| IntermediatePheno | outcome | exposure | method | nsnp | Beta | 95%CI | b | se | pval | RSSobs | Phet | Q | Q_df | Q_pval | egger_intercept | egger_se | egger_pval | Lower95 | Upper95 | MeanFstat |
|---|---|---|---|---|---|---|---|---|---|---|---|---|---|---|---|---|---|---|---|---|
| SBP | OveralleGFRDeclAdjDM | ACEI | Wald ratio | 1 | -0.074 | (-0.392, 0.244) | -0.073969708 | 0.162028883 | 0.648014842 | | | | | | | | | -0.391546319 | 0.243606904 | 82.79833374 |
| SBP | OveralleGFRDeclAdjDM | CCB | Inverse variance weighted | 24 | 0.012 | (-0.061, 0.084) | 0.011526968 | 0.036797503 | 0.754087639 | | | 21.47717745 | 23 | 0.551971516 | -0.03133263 | 0.029467259 | 0.299177127 | -0.060596139 | 0.083650075 | 66.21360566 |
| SBP | OveralleGFRDeclAdjDM | CCB | MR Egger | 24 | 0.102 | (-0.080, 0.283) | 0.101882898 | 0.092601756 | 0.283127029 | | | 20.34656376 | 22 | 0.561371199 | -0.03133263 | 0.029467259 | 0.299177127 | -0.079616543 | 0.28338234 | 66.21360566 |
| SBP | OveralleGFRDeclAdjDM | CCB | MR.PRESSO.Outlier-corrected | 24 | | (NA, NA) | | | | 24.19038187 | 0.514 | | | | | | | | | 66.21360566 |
| SBP | OveralleGFRDeclAdjDM | CCB | MR.PRESSO.Raw | 24 | 0.012 | (-0.058, 0.081) | 0.011526968 | 0.035558468 | 0.748739413 | 24.19038187 | 0.514 | | | | | | | -0.058167629 | 0.081221565 | 66.21360566 |
| SBP | OveralleGFRDeclAdjDM | BB | Inverse variance weighted | 5 | -0.019 | (-0.217, 0.180) | -0.018566875 | 0.101465868 | 0.854808678 | | | 4.749131378 | 4 | 0.314019857 | 0.061001801 | 0.148577938 | 0.708950172 | -0.217439977 | 0.180306226 | 52.2808789 |
| SBP | OveralleGFRDeclAdjDM | BB | MR Egger | 5 | -0.211 | (-1.158, 0.735) | -0.211272656 | 0.483007208 | 0.691388274 | | | 4.496476475 | 3 | 0.212604793 | 0.061001801 | 0.148577938 | 0.708950172 | -1.157966783 | 0.735421472 | 52.2808789 |
| SBP | OveralleGFRDeclAdjDM | BB | MR.PRESSO.Outlier-corrected | 5 | | (NA, NA) | | | | 6.696074104 | 0.401 | | | | | | | | | 52.2808789 |
| SBP | OveralleGFRDeclAdjDM | BB | MR.PRESSO.Raw | 5 | -0.019 | (-0.217, 0.180) | -0.018566875 | 0.101465868 | 0.863709212 | 6.696074104 | 0.401 | | | | | | | -0.217439977 | 0.180306226 | 52.2808789 |
| SBP | OveralleGFRDeclAdjDM | Lowering SBP | Inverse variance weighted | 440 | 0.045 | (0.025, 0.064) | 0.044655516 | 0.009859275 | 5.91822E-06 | | | 676.5213143 | 439 | 2.16565E-12 | 0.022897982 | 0.007558255 | 0.002594273 | 0.025331337 | 0.063979695 | 75.12872998 |
| SBP | OveralleGFRDeclAdjDM | Lowering SBP | MR Egger | 440 | -0.024 | (-0.072, 0.024) | -0.02401482 | 0.024682366 | 0.331112903 | | | 662.6361077 | 438 | 2.02768E-11 | 0.022897982 | 0.007558255 | 0.002594273 | -0.072392257 | 0.024362617 | 75.12872998 |
| SBP | OveralleGFRDeclAdjDM | Lowering SBP | MR.PRESSO.Outlier-corrected | 440 | 0.044 | (0.027, 0.061) | 0.043750544 | 0.008588329 | 5.22014E-07 | 680.6891113 | <0.001 | | | | | | | 0.026917419 | 0.06058367 | 75.12872998 |
| SBP | OveralleGFRDeclAdjDM | Lowering SBP | MR.PRESSO.Raw | 440 | 0.045 | (0.025, 0.064) | 0.044655516 | 0.009859275 | 7.63628E-06 | 680.6891113 | <0.001 | | | | | | | 0.025331337 | 0.063979695 | 75.12872998 |
| LDL | OveralleGFRDeclAdjDM | PCSK9I | Inverse variance weighted | 11 | 0.066 | (0.008, 0.125) | 0.066484234 | 0.03007978 | 0.027086896 | | | 8.968769153 | 10 | 0.535070052 | 0.000804476 | 0.003063418 | 0.798765606 | 0.007527864 | 0.125440604 | 869.0826758 |
| LDL | OveralleGFRDeclAdjDM | PCSK9I | MR Egger | 11 | 0.055 | (-0.046, 0.157) | 0.05544734 | 0.051683225 | 0.311269461 | | | 8.899806511 | 9 | 0.446573952 | 0.000804476 | 0.003063418 | 0.798765606 | -0.045851782 | 0.156746462 | 869.0826758 |
| LDL | OveralleGFRDeclAdjDM | PCSK9I | MR.PRESSO.Outlier-corrected | 11 | | (NA, NA) | | | | 9.910074284 | 0.624 | | | | | | | | | 869.0826758 |
| LDL | OveralleGFRDeclAdjDM | PCSK9I | MR.PRESSO.Raw | 11 | 0.066 | (0.011, 0.122) | 0.066484234 | 0.028486631 | 0.041773434 | 9.910074284 | 0.624 | | | | | | | 0.010650438 | 0.12231803 | 869.0826758 |
| LDL | OveralleGFRDeclAdjDM | Lowering LDL | Inverse variance weighted | 229 | -0.002 | (-0.025, 0.022) | -0.001841094 | 0.012025596 | 0.878321042 | | | 243.7237586 | 228 | 0.226294379 | -0.001540891 | 0.000564327 | 0.006820352 | -0.025411263 | 0.021729075 | 240.6474171 |
| LDL | OveralleGFRDeclAdjDM | Lowering LDL | MR Egger | 229 | 0.035 | (-0.000, 0.071) | 0.035234763 | 0.018027956 | 0.051875692 | | | 235.9734376 | 227 | 0.327469089 | -0.001540891 | 0.000564327 | 0.006820352 | -0.000100031 | 0.070569556 | 240.6474171 |
| LDL | OveralleGFRDeclAdjDM | Lowering LDL | MR.PRESSO.Outlier-corrected | 229 | | (NA, NA) | | | | 246.0707918 | 0.228 | | | | | | | | | 240.6474171 |
| LDL | OveralleGFRDeclAdjDM | Lowering LDL | MR.PRESSO.Raw | 229 | -0.002 | (-0.025, 0.022) | -0.001841094 | 0.012025596 | 0.87845645 | 246.0707918 | 0.228 | | | | | | | -0.025411263 | 0.021729075 | 240.6474171 |
| LDL | OveralleGFRDeclAdjDM | Statin | Inverse variance weighted | 5 | 0.014 | (-0.072, 0.100) | 0.014152871 | 0.043792785 | 0.746560325 | | | 2.77894134 | 4 | 0.595473254 | -0.018599223 | 0.013681847 | 0.267176709 | -0.071680986 | 0.099986729 | 787.7134762 |
| LDL | OveralleGFRDeclAdjDM | Statin | MR Egger | 5 | 0.322 | (-0.130, 0.775) | 0.322299148 | 0.230868203 | 0.257077105 | | | 0.93094909 | 3 | 0.81795338 | -0.018599223 | 0.013681847 | 0.267176709 | -0.13020253 | 0.774800827 | 787.7134762 |
| LDL | OveralleGFRDeclAdjDM | Statin | MR.PRESSO.Outlier-corrected | 5 | | (NA, NA) | | | | 3.947468486 | 0.665 | | | | | | | | | 787.7134762 |
| LDL | OveralleGFRDeclAdjDM | Statin | MR.PRESSO.Raw | 5 | 0.014 | (-0.057, 0.086) | 0.014152871 | 0.03650163 | 0.71796292 | 3.947468486 | 0.665 | | | | | | | -0.057390323 | 0.085696065 | 787.7134762 |
| LDL | OveralleGFRDeclAdjDM | NPC1L1I | Inverse variance weighted | 6 | 0.052 | (-0.097, 0.200) | 0.051639295 | 0.075584688 | 0.494481838 | | | 0.6295358 | 5 | 0.986611029 | -0.004408996 | 0.009174412 | 0.655920602 | -0.096506692 | 0.199785283 | 191.4711596 |
| LDL | OveralleGFRDeclAdjDM | NPC1L1I | MR Egger | 6 | 0.191 | (-0.398, 0.780) | 0.19142374 | 0.300529196 | 0.558782885 | | | 0.39858316 | 4 | 0.98259274 | -0.004408996 | 0.009174412 | 0.655920602 | -0.397613485 | 0.780460965 | 191.4711596 |
| LDL | OveralleGFRDeclAdjDM | NPC1L1I | MR.PRESSO.Outlier-corrected | 6 | | (NA, NA) | | | | 1.36873682 | 0.973 | | | | | | | | | 191.4711596 |
| LDL | OveralleGFRDeclAdjDM | NPC1L1I | MR.PRESSO.Raw | 6 | 0.052 | (-0.001, 0.104) | 0.051639295 | 0.026820016 | 0.112149378 | 1.36873682 | 0.973 | | | | | | | -0.000927937 | 0.104206527 | 191.4711596 |
| HbA1c | OveralleGFRDeclAdjDM | MG3 | Wald ratio | 1 | 0.654 | (-0.404, 1.711) | 0.653743981 | 0.539598207 | 0.225689103 | | | | | | | | | -0.403868504 | 1.711356467 | 13.3793901 |
| HbA1c | OveralleGFRDeclAdjDM | MC1 | Inverse variance weighted | 25 | -0.102 | (-0.243, 0.039) | -0.101836906 | 0.071926817 | 0.156822185 | | | 24.76439157 | 24 | 0.418653163 | 0.000933115 | 0.002339322 | 0.693658877 | -0.242813467 | 0.039139655 | 30.29781153 |
| HbA1c | OveralleGFRDeclAdjDM | MC1 | MR Egger | 25 | -0.149 | (-0.424, 0.125) | -0.14940298 | 0.139933768 | 0.296748549 | | | 24.59425551 | 23 | 0.371539059 | 0.000933115 | 0.002339322 | 0.693658877 | -0.423673166 | 0.124867206 | 30.29781153 |
| HbA1c | OveralleGFRDeclAdjDM | MC1 | MR.PRESSO.Outlier-corrected | 25 | | (NA, NA) | | | | 28.0084957 | 0.397 | | | | | | | | | 30.29781153 |
| HbA1c | OveralleGFRDeclAdjDM | MC1 | MR.PRESSO.Raw | 25 | -0.102 | (-0.243, 0.039) | -0.101836906 | 0.071926817 | 0.169669704 | 28.0084957 | 0.397 | | | | | | | -0.242813467 | 0.039139655 | 30.29781153 |
| HbA1c | OveralleGFRDeclAdjDM | Lowering HbA1c | Inverse variance weighted | 287 | 0.016 | (-0.005, 0.037) | 0.015986762 | 0.010515984 | 0.128452045 | | | 291.761669 | 286 | 0.394680138 | 0.001373793 | 0.000588269 | 0.02022098 | -0.004624567 | 0.036598091 | 123.2003391 |
| HbA1c | OveralleGFRDeclAdjDM | Lowering HbA1c | MR Egger | 287 | -0.021 | (-0.058, 0.016) | -0.020878801 | 0.018923318 | 0.27081149 | | | 286.2834161 | 285 | 0.467484888 | 0.001373793 | 0.000588269 | 0.02022098 | -0.057968503 | 0.016210902 | 123.2003391 |
| HbA1c | OveralleGFRDeclAdjDM | Lowering HbA1c | MR.PRESSO.Outlier-corrected | 287 | | (NA, NA) | | | | 293.6611879 | 0.399 | | | | | | | | | 123.2003391 |
| HbA1c | OveralleGFRDeclAdjDM | Lowering HbA1c | MR.PRESSO.Raw | 287 | 0.016 | (-0.005, 0.037) | 0.015986762 | 0.010515984 | 0.129556787 | 293.6611879 | 0.399 | | | | | | | -0.004624567 | 0.036598091 | 123.2003391 |
| HbA1c | OveralleGFRDeclAdjDM | GLP1RA | Inverse variance weighted | 4 | 0.063 | (-0.326, 0.452) | 0.063086556 | 0.198647796 | 0.750803589 | | | 0.445189927 | 3 | 0.93075695 | -0.004254938 | 0.006382211 | 0.573587804 | -0.326263125 | 0.452436237 | 23.04119877 |
| HbA1c | OveralleGFRDeclAdjDM | GLP1RA | MR Egger | 4 | 0.333 | (-0.551, 1.217) | 0.33311519 | 0.451121511 | 0.537155591 | | | 0.00071806 | 2 | 0.999641034 | -0.004254938 | 0.006382211 | 0.573587804 | -0.55108297 | 1.217313351 | 23.04119877 |
| HbA1c | OveralleGFRDeclAdjDM | GLP1RA | MR.PRESSO.Outlier-corrected | 4 | | (NA, NA) | | | | 0.864701246 | 0.933 | | | | | | | | | 23.04119877 |
| HbA1c | OveralleGFRDeclAdjDM | GLP1RA | MR.PRESSO.Raw | 4 | 0.063 | (-0.087, 0.213) | 0.063086556 | 0.07652367 | 0.470143012 | 0.864701246 | 0.933 | | | | | | | -0.086899837 | 0.213072949 | 23.04119877 |
| HbA1c | OveralleGFRDeclAdjDM | GCG | Wald ratio | 1 | -1.749 | (-3.299, -0.199) | -1.749100343 | 0.79086116 | 0.026991535 | | | | | | | | | -3.299188217 | -0.19901247 | 4.12399181 |
| HbA1c | OveralleGFRDeclAdjDM | AMPK | Inverse variance weighted | 3 | 0.035 | (-0.773, 0.842) | 0.034530243 | 0.411901932 | 0.933190623 | | | 3.894468731 | 2 | 0.142668094 | 0.022849964 | 0.015964536 | 0.388230697 | -0.772797545 | 0.84185803 | 14.59673982 |
| HbA1c | OveralleGFRDeclAdjDM | AMPK | MR Egger | 3 | -1.704 | (-4.172, 0.765) | -1.703624688 | 1.259387057 | 0.405258711 | | | 1.277458803 | 1 | 0.258372063 | 0.022849964 | 0.015964536 | 0.388230697 | -4.172023319 | 0.764773943 | 14.59673982 |
| HbA1c | OveralleGFRDeclAdjDM | GDF15 | Wald ratio | 1 | -0.534 | (-1.563, 0.495) | -0.534358301 | 0.524983594 | 0.308745851 | | | | | | | | | -1.563326146 | 0.494609543 | 13.33861351 |
| HbA1c | OveralleGFRDeclAdjDM | SGLT2I | Inverse variance weighted | 6 | -0.41 | (-0.698, -0.123) | -0.410067711 | 0.146669192 | 0.005176052 | | | 0.548904197 | 5 | 0.990223023 | 0.000264373 | 0.014503863 | 0.986330146 | -0.697539326 | -0.122596096 | 29.79338183 |
| HbA1c | OveralleGFRDeclAdjDM | SGLT2I | MR Egger | 6 | -0.429 | (-2.440, 1.583) | -0.428580655 | 1.026182661 | 0.697650556 | | | 0.548571946 | 4 | 0.968603464 | 0.000264373 | 0.014503863 | 0.986330146 | -2.439898671 | 1.582737362 | 29.79338183 |
| HbA1c | OveralleGFRDeclAdjDM | SGLT2I | MR.PRESSO.Outlier-corrected | 6 | | (NA, NA) | | | | 0.842850428 | 0.988 | | | | | | | | | 29.79338183 |
| HbA1c | OveralleGFRDeclAdjDM | SGLT2I | MR.PRESSO.Raw | 6 | -0.41 | (-0.505, -0.315) | -0.410067711 | 0.048596184 | 0.000383541 | 0.842850428 | 0.988 | | | | | | | -0.505316233 | -0.31481919 | 29.79338183 |

### Table S3. The effect of genetically proxied antihypertensives, lipid-lowering, and antidiabetic drugs on creatinine-based cross-sectional eGFR, all ancestries

| IntermediatePheno | outcome | exposure | method | nsnp | Beta | 95%CI | b | se | pval | RSSobs | Phet | Q | Q_df | Q_pval | egger_intercept | egger_se | egger_pval | Lower95 | Upper95 | MeanFstat |
|---|---|---|---|---|---|---|---|---|---|---|---|---|---|---|---|---|---|---|---|---|
| SBP | eGFRCreatStanzick | ACEI | Wald ratio | 1 | 0.021 | (-0.002, 0.045) | 0.021134202 | 0.011976048 | 0.077613209 | | | | | | | | | -0.002338852 | 0.044607256 | 82.79833374 |
| SBP | eGFRCreatStanzick | CCB | Inverse variance weighted | 24 | 0.005 | (-0.000, 0.010) | 0.005166443 | 0.002666013 | 0.052636476 | | | 25.49405889 | 23 | 0.325324937 | 7.65523E-05 | 0.002227194 | 0.972890777 | -5.89419E-05 | 0.010391827 | 66.21360566 |
| SBP | eGFRCreatStanzick | CCB | MR Egger | 24 | 0.005 | (-0.008, 0.018) | 0.004951884 | 0.006811518 | 0.4748994 | | | 25.49268992 | 22 | 0.274098966 | 7.65523E-05 | 0.002227194 | 0.972890777 | -0.00839869 | 0.018302459 | 66.21360566 |
| SBP | eGFRCreatStanzick | CCB | MR.PRESSO.Outlier-corrected | 24 | | (NA, NA) | | | | 27.6231051 | 0.328 | | | | | | | | | 66.21360566 |
| SBP | eGFRCreatStanzick | CCB | MR.PRESSO.Raw | 24 | 0.005 | (-0.000, 0.010) | 0.005166443 | 0.002666013 | 0.065008658 | 27.6231051 | 0.328 | | | | | | | -5.89419E-05 | 0.010391827 | 66.21360566 |
| SBP | eGFRCreatStanzick | BB | Inverse variance weighted | 5 | -0.006 | (-0.022, 0.011) | -0.005720095 | 0.008541414 | 0.503055713 | | | 7.494793148 | 4 | 0.111939112 | 0.002999493 | 0.012255934 | 0.822444259 | -0.022461266 | 0.011021075 | 52.2808789 |
| SBP | eGFRCreatStanzick | BB | MR Egger | 5 | -0.015 | (-0.093, 0.063) | -0.015123121 | 0.039642492 | 0.728257341 | | | 7.34808453 | 3 | 0.061593014 | 0.002999493 | 0.012255934 | 0.822444259 | -0.092822406 | 0.062576163 | 52.2808789 |
| SBP | eGFRCreatStanzick | BB | MR.PRESSO.Outlier-corrected | 5 | | (NA, NA) | | | | 12.72075492 | 0.172 | | | | | | | | | 52.2808789 |
| SBP | eGFRCreatStanzick | BB | MR.PRESSO.Raw | 5 | -0.006 | (-0.022, 0.011) | -0.005720095 | 0.008541414 | 0.53972955 | 12.72075492 | 0.172 | | | | | | | -0.022461266 | 0.011021075 | 52.2808789 |
| SBP | eGFRCreatStanzick | Lowering SBP | Inverse variance weighted | 436 | 0.003 | (-0.001, 0.006) | 0.002694223 | 0.001899444 | 0.156066073 | | | 5015.097084 | 435 | 0 | -0.001180112 | 0.001463513 | 0.420479536 | -0.001028687 | 0.006417134 | 75.12872998 |
| SBP | eGFRCreatStanzick | Lowering SBP | MR Egger | 436 | 0.006 | (-0.003, 0.015) | 0.006129019 | 0.004664273 | 0.189528491 | | | 5007.594816 | 434 | 0 | -0.001180112 | 0.001463513 | 0.420479536 | -0.003012957 | 0.015270994 | 75.12872998 |
| SBP | eGFRCreatStanzick | Lowering SBP | MR.PRESSO.Outlier-corrected | 436 | 0.003 | (0.002, 0.005) | 0.003372568 | 0.000882563 | 0.000155462 | 5047.019042 | <0.001 | | | | | | | 0.001642745 | 0.005102392 | 75.12872998 |
| SBP | eGFRCreatStanzick | Lowering SBP | MR.PRESSO.Raw | 436 | 0.003 | (-0.001, 0.006) | 0.002694223 | 0.001899444 | 0.156782089 | 5047.019042 | <0.001 | | | | | | | -0.001028687 | 0.006417134 | 75.12872998 |
| LDL | eGFRCreatStanzick | PCSK9I | Inverse variance weighted | 11 | 0 | (-0.008, 0.009) | 0.00034537 | 0.004222157 | 0.934806223 | | | 51.67145502 | 10 | 1.31255E-07 | 0.001022091 | 0.00031591 | 0.01023396 | -0.007930058 | 0.008620798 | 869.0826758 |
| LDL | eGFRCreatStanzick | PCSK9I | MR Egger | 11 | -0.012 | (-0.022, -0.002) | -0.012052096 | 0.004882617 | 0.035665636 | | | 23.88789213 | 9 | 0.004482114 | 0.001022091 | 0.00031591 | 0.01023396 | -0.021622026 | -0.002482167 | 869.0826758 |
| LDL | eGFRCreatStanzick | PCSK9I | MR.PRESSO.Outlier-corrected | 11 | 0.003 | (-0.004, 0.009) | 0.002820699 | 0.003350547 | 0.424314666 | 75.51541352 | <0.001 | | | | | | | -0.003746372 | 0.009387771 | 869.0826758 |
| LDL | eGFRCreatStanzick | PCSK9I | MR.PRESSO.Raw | 11 | 0 | (-0.008, 0.009) | 0.00034537 | 0.004222157 | 0.936420313 | 75.51541352 | <0.001 | | | | | | | -0.007930058 | 0.008620798 | 869.0826758 |
| LDL | eGFRCreatStanzick | Lowering LDL | Inverse variance weighted | 224 | -0.006 | (-0.010, -0.002) | -0.005839386 | 0.002030503 | 0.004029629 | | | 1451.12242 | 223 | 7.0112E-179 | 6.08278E-06 | 9.85036E-05 | 0.950815985 | -0.009819172 | -0.001859601 | 243.5409148 |
| LDL | eGFRCreatStanzick | Lowering LDL | MR Egger | 224 | -0.006 | (-0.012, -0.000) | -0.005977088 | 0.003018939 | 0.048954096 | | | 1451.097494 | 222 | 2.7661E-179 | 6.08278E-06 | 9.85036E-05 | 0.950815985 | -0.01189421 | -5.9967E-05 | 243.5409148 |
| LDL | eGFRCreatStanzick | Lowering LDL | MR.PRESSO.Outlier-corrected | 224 | -0.006 | (-0.008, -0.003) | -0.005673567 | 0.001364276 | 4.89351E-05 | 1464.201733 | <0.001 | | | | | | | -0.008347548 | -0.002999587 | 243.5409148 |
| LDL | eGFRCreatStanzick | Lowering LDL | MR.PRESSO.Raw | 224 | -0.006 | (-0.010, -0.002) | -0.005839386 | 0.002030503 | 0.004420157 | 1464.201733 | <0.001 | | | | | | | -0.009819172 | -0.001859601 | 243.5409148 |
| LDL | eGFRCreatStanzick | Statin | Inverse variance weighted | 5 | -0.012 | (-0.018, -0.006) | -0.01196108 | 0.003088822 | 0.00010778 | | | 1.295516748 | 4 | 0.862135722 | 0.000572941 | 0.000895191 | 0.567685704 | -0.018015172 | -0.005906989 | 787.7134762 |
| LDL | eGFRCreatStanzick | Statin | MR Egger | 5 | -0.021 | (-0.051, 0.008) | -0.021474871 | 0.015182336 | 0.252149175 | | | 0.88588974 | 3 | 0.828831535 | 0.000572941 | 0.000895191 | 0.567685704 | -0.051232249 | 0.008282508 | 787.7134762 |
| LDL | eGFRCreatStanzick | Statin | MR.PRESSO.Outlier-corrected | 5 | | (NA, NA) | | | | 2.291346897 | 0.83 | | | | | | | | | 787.7134762 |
| LDL | eGFRCreatStanzick | Statin | MR.PRESSO.Raw | 5 | -0.012 | (-0.015, -0.009) | -0.01196108 | 0.001757861 | 0.002437384 | 2.291346897 | 0.83 | | | | | | | -0.015406487 | -0.008515674 | 787.7134762 |
| LDL | eGFRCreatStanzick | NPC1L1I | Inverse variance weighted | 6 | -0.015 | (-0.029, -0.002) | -0.01539985 | 0.006905953 | 0.025751529 | | | 8.93583618 | 5 | 0.111650969 | -0.000762012 | 0.000843526 | 0.417416381 | -0.028935517 | -0.001864182 | 191.4711596 |
| LDL | eGFRCreatStanzick | NPC1L1I | MR Egger | 6 | 0.008 | (-0.045, 0.061) | 0.008281312 | 0.02714236 | 0.775502136 | | | 7.421685295 | 4 | 0.115212636 | -0.000762012 | 0.000843526 | 0.417416381 | -0.044917713 | 0.061480337 | 191.4711596 |
| LDL | eGFRCreatStanzick | NPC1L1I | MR.PRESSO.Outlier-corrected | 6 | | (NA, NA) | | | | 11.28973193 | 0.208 | | | | | | | | | 191.4711596 |
| LDL | eGFRCreatStanzick | NPC1L1I | MR.PRESSO.Raw | 6 | -0.015 | (-0.029, -0.002) | -0.01539985 | 0.006905953 | 0.076170918 | 11.28973193 | 0.208 | | | | | | | -0.028935517 | -0.001864182 | 191.4711596 |
| HbA1c | eGFRCreatStanzick | MG3 | Wald ratio | 1 | 0 | (-0.069, 0.069) | 0 | 0.035281421 | 1 | | | | | | | | | -0.069151586 | 0.069151586 | 13.3793901 |
| HbA1c | eGFRCreatStanzick | MC1 | Inverse variance weighted | 24 | 0.022 | (0.001, 0.043) | 0.021924575 | 0.010581672 | 0.038271163 | | | 114.3554923 | 23 | 4.26097E-14 | 7.74338E-05 | 0.000353511 | 0.828636593 | 0.001184498 | 0.042664653 | 31.06201727 |
| HbA1c | eGFRCreatStanzick | MC1 | MR Egger | 24 | 0.018 | (-0.023, 0.059) | 0.017986677 | 0.020976372 | 0.400428402 | | | 114.1066388 | 22 | 2.02736E-14 | 7.74338E-05 | 0.000353511 | 0.828636593 | -0.023127012 | 0.059100367 | 31.06201727 |
| HbA1c | eGFRCreatStanzick | MC1 | MR.PRESSO.Outlier-corrected | 24 | 0.018 | (0.005, 0.031) | 0.017905024 | 0.006453735 | 0.011701106 | 134.9884139 | <0.001 | | | | | | | 0.005255703 | 0.030554344 | 31.06201727 |
| HbA1c | eGFRCreatStanzick | MC1 | MR.PRESSO.Raw | 24 | 0.022 | (0.001, 0.043) | 0.021924575 | 0.010581672 | 0.049666832 | 134.9884139 | <0.001 | | | | | | | 0.001184498 | 0.042664653 | 31.06201727 |
| HbA1c | eGFRCreatStanzick | Lowering HbA1c | Inverse variance weighted | 276 | 0 | (-0.003, 0.004) | 0.000456766 | 0.001838023 | 0.803740255 | | | 1819.194232 | 275 | 1.9457E-225 | 3.26242E-05 | 0.000106052 | 0.758601493 | -0.003145758 | 0.00405929 | 123.8837706 |
| HbA1c | eGFRCreatStanzick | Lowering HbA1c | MR Egger | 276 | 0 | (-0.007, 0.006) | -0.000405241 | 0.003352828 | 0.903886229 | | | 1818.566143 | 274 | 9.8481E-226 | 3.26242E-05 | 0.000106052 | 0.758601493 | -0.006976784 | 0.006166302 | 123.8837706 |
| HbA1c | eGFRCreatStanzick | Lowering HbA1c | MR.PRESSO.Outlier-corrected | 276 | 0 | (-0.002, 0.002) | -2.79192E-05 | 0.001200842 | 0.981470019 | 1832.949486 | <0.001 | | | | | | | -0.00238157 | 0.002325732 | 123.8837706 |
| HbA1c | eGFRCreatStanzick | Lowering HbA1c | MR.PRESSO.Raw | 276 | 0 | (-0.003, 0.004) | 0.000456766 | 0.001838023 | 0.803925735 | 1832.949486 | <0.001 | | | | | | | -0.003145758 | 0.00405929 | 123.8837706 |
| HbA1c | eGFRCreatStanzick | GLP1RA | Inverse variance weighted | 4 | 0.001 | (-0.026, 0.027) | 0.000567131 | 0.013569155 | 0.966661648 | | | 0.776708255 | 3 | 0.855029611 | -0.000101728 | 0.000454676 | 0.843737856 | -0.026028414 | 0.027162676 | 23.04119877 |
| HbA1c | eGFRCreatStanzick | GLP1RA | MR Egger | 4 | 0.007 | (-0.052, 0.065) | 0.00653491 | 0.029926311 | 0.847399849 | | | 0.72665023 | 2 | 0.695360325 | -0.000101728 | 0.000454676 | 0.843737856 | -0.05212066 | 0.065190481 | 23.04119877 |
| HbA1c | eGFRCreatStanzick | GLP1RA | MR.PRESSO.Outlier-corrected | 4 | | (NA, NA) | | | | 1.77188338 | 0.825 | | | | | | | | | 23.04119877 |
| HbA1c | eGFRCreatStanzick | GLP1RA | MR.PRESSO.Raw | 4 | 0.001 | (-0.013, 0.014) | 0.000567131 | 0.006904324 | 0.939707775 | 1.77188338 | 0.825 | | | | | | | -0.012965344 | 0.014099606 | 23.04119877 |
| HbA1c | eGFRCreatStanzick | GCG | Wald ratio | 1 | -0.092 | (-0.222, 0.038) | -0.092057913 | 0.066407231 | 0.165666491 | | | | | | | | | -0.222216085 | 0.038100259 | 4.12399181 |
| HbA1c | eGFRCreatStanzick | AMPK | Inverse variance weighted | 3 | 0.02 | (-0.033, 0.072) | 0.019523291 | 0.02693345 | 0.468530849 | | | 3.740748729 | 2 | 0.154065974 | -0.001091634 | 0.001436105 | 0.586224956 | -0.033266271 | 0.072312854 | 14.59673982 |
| HbA1c | eGFRCreatStanzick | AMPK | MR Egger | 3 | 0.104 | (-0.122, 0.330) | 0.104111549 | 0.115338063 | 0.532539567 | | | 2.37085476 | 1 | 0.123619348 | -0.001091634 | 0.001436105 | 0.586224956 | -0.121951053 | 0.330174152 | 14.59673982 |
| HbA1c | eGFRCreatStanzick | GDF15 | Wald ratio | 1 | 0.094 | (0.031, 0.156) | 0.09374707 | 0.031874004 | 0.003269682 | | | | | | | | | 0.031274023 | 0.156220118 | 13.33861351 |
| HbA1c | eGFRCreatStanzick | SGLT2I | Inverse variance weighted | 6 | 0.024 | (0.004, 0.045) | 0.02438242 | 0.010481106 | 0.020001419 | | | 0.9895219 | 5 | 0.963406469 | 0.000608672 | 0.001017322 | 0.581865048 | 0.003839452 | 0.044925389 | 29.79338183 |
| HbA1c | eGFRCreatStanzick | SGLT2I | MR Egger | 6 | -0.019 | (-0.161, 0.124) | -0.018602785 | 0.072605156 | 0.810419771 | | | 0.631549927 | 4 | 0.959494068 | 0.000608672 | 0.001017322 | 0.581865048 | -0.160908891 | 0.123703321 | 29.79338183 |
| HbA1c | eGFRCreatStanzick | SGLT2I | MR.PRESSO.Outlier-corrected | 6 | | (NA, NA) | | | | 1.62259003 | 0.955 | | | | | | | | | 29.79338183 |
| HbA1c | eGFRCreatStanzick | SGLT2I | MR.PRESSO.Raw | 6 | 0.024 | (0.015, 0.034) | 0.02438242 | 0.004662672 | 0.003383749 | 1.62259003 | 0.955 | | | | | | | 0.015243584 | 0.033521257 | 29.79338183 |

### Table S4. The effect of genetically proxied antihypertensives, lipid-lowering, and antidiabetic drugs on creatinine-based cross-sectional eGFR, European only

| IntermediatePheno | outcome | exposure | method | nsnp | Beta | 95%CI | b | se | pval | RSSobs | Phet | Q | Q_df | Q_pval | egger_intercept | egger_se | egger_pval | Lower95 | Upper95 | MeanFstat |
|---|---|---|---|---|---|---|---|---|---|---|---|---|---|---|---|---|---|---|---|---|
| SBP | eGFREurStanzick | ACEI | Wald ratio | 1 | 0.018 | (-0.003, 0.040) | 0.018316309 | 0.01111659 | 0.099423434 | | | | | | | | | -0.003472209 | 0.040104826 | 82.79833374 |
| SBP | eGFREurStanzick | CCB | Inverse variance weighted | 24 | 0.006 | (0.001, 0.011) | 0.005856034 | 0.002529076 | 0.020586497 | | | 23.39792963 | 23 | 0.43774507 | 0.000273381 | 0.002056238 | 0.895439946 | 0.000899044 | 0.010813024 | 66.21360566 |
| SBP | eGFREurStanzick | CCB | MR Egger | 24 | 0.005 | (-0.007, 0.018) | 0.00508134 | 0.006374484 | 0.433890614 | | | 23.37914531 | 22 | 0.380592172 | 0.000273381 | 0.002056238 | 0.895439946 | -0.007412649 | 0.017575328 | 66.21360566 |
| SBP | eGFREurStanzick | CCB | MR.PRESSO.Outlier-corrected | 24 | | (NA, NA) | | | | 25.45822244 | 0.448 | | | | | | | | | 66.21360566 |
| SBP | eGFREurStanzick | CCB | MR.PRESSO.Raw | 24 | 0.006 | (0.001, 0.011) | 0.005856034 | 0.002529076 | 0.029855854 | 25.45822244 | 0.448 | | | | | | | 0.000899044 | 0.010813024 | 66.21360566 |
| SBP | eGFREurStanzick | BB | Inverse variance weighted | 5 | -0.011 | (-0.030, 0.008) | -0.011378825 | 0.009664386 | 0.239036838 | | | 9.21721525 | 4 | 0.055893615 | -0.000393591 | 0.013986217 | 0.979316814 | -0.030321022 | 0.007563372 | 52.2808789 |
| SBP | eGFREurStanzick | BB | MR Egger | 5 | -0.01 | (-0.098, 0.078) | -0.010156267 | 0.044853576 | 0.835416735 | | | 9.214782751 | 3 | 0.026567421 | -0.000393591 | 0.013986217 | 0.979316814 | -0.098069276 | 0.077756743 | 52.2808789 |
| SBP | eGFREurStanzick | BB | MR.PRESSO.Outlier-corrected | 5 | | (NA, NA) | | | | 14.60004001 | 0.095 | | | | | | | | | 52.2808789 |
| SBP | eGFREurStanzick | BB | MR.PRESSO.Raw | 5 | -0.011 | (-0.030, 0.008) | -0.011378825 | 0.009664386 | 0.304308888 | 14.60004001 | 0.095 | | | | | | | -0.030321022 | 0.007563372 | 52.2808789 |
| SBP | eGFREurStanzick | Lowering SBP | Inverse variance weighted | 441 | 0.001 | (-0.003, 0.005) | 0.000809874 | 0.001917366 | 0.672741217 | | | 5342.303533 | 440 | 0 | -0.001204011 | 0.001482254 | 0.417068827 | -0.002948163 | 0.004567911 | 75.12872998 |
| SBP | eGFREurStanzick | Lowering SBP | MR Egger | 441 | 0.004 | (-0.005, 0.014) | 0.00441845 | 0.004838903 | 0.36168595 | | | 5334.286239 | 439 | 0 | -0.001204011 | 0.001482254 | 0.417068827 | -0.0050658 | 0.013902699 | 75.12872998 |
| SBP | eGFREurStanzick | Lowering SBP | MR.PRESSO.Outlier-corrected | 441 | 0.002 | (0.000, 0.004) | 0.002150514 | 0.000890738 | 0.016256567 | 5372.356893 | <0.001 | | | | | | | 0.000404667 | 0.003896361 | 75.12872998 |
| SBP | eGFREurStanzick | Lowering SBP | MR.PRESSO.Raw | 441 | 0.001 | (-0.003, 0.005) | 0.000809874 | 0.001917366 | 0.672947543 | 5372.356893 | <0.001 | | | | | | | -0.002948163 | 0.004567911 | 75.12872998 |
| LDL | eGFREurStanzick | PCSK9I | Inverse variance weighted | 11 | 0.001 | (-0.008, 0.009) | 0.000556127 | 0.004247437 | 0.895828714 | | | 51.30076424 | 10 | 1.53683E-07 | 0.001041224 | 0.000287975 | 0.00560947 | -0.007768848 | 0.008881103 | 869.0826758 |
| LDL | eGFREurStanzick | PCSK9I | MR Egger | 11 | -0.012 | (-0.020, -0.003) | -0.011605396 | 0.004414374 | 0.027403475 | | | 20.91718781 | 9 | 0.013023237 | 0.001041224 | 0.000287975 | 0.00560947 | -0.020257569 | -0.002953223 | 869.0826758 |
| LDL | eGFREurStanzick | PCSK9I | MR.PRESSO.Outlier-corrected | 11 | 0.005 | (-0.002, 0.013) | 0.005463315 | 0.003705445 | 0.178603844 | 83.0393874 | 0.001 | | | | | | | -0.001799358 | 0.012725988 | 869.0826758 |
| LDL | eGFREurStanzick | PCSK9I | MR.PRESSO.Raw | 11 | 0.001 | (-0.008, 0.009) | 0.000556127 | 0.004247437 | 0.898425322 | 83.0393874 | 0.001 | | | | | | | -0.007768848 | 0.008881103 | 869.0826758 |
| LDL | eGFREurStanzick | Lowering LDL | Inverse variance weighted | 228 | -0.007 | (-0.011, -0.003) | -0.006826961 | 0.002120283 | 0.001282647 | | | 1650.246051 | 227 | 3.1917E-214 | -1.21103E-05 | 0.000101294 | 0.904940683 | -0.010982716 | -0.002671206 | 241.4800402 |
| LDL | eGFREurStanzick | Lowering LDL | MR Egger | 228 | -0.007 | (-0.013, -0.000) | -0.006546949 | 0.003162373 | 0.039563844 | | | 1650.141685 | 226 | 1.2334E-214 | -1.21103E-05 | 0.000101294 | 0.904940683 | -0.0127452 | -0.000348697 | 241.4800402 |
| LDL | eGFREurStanzick | Lowering LDL | MR.PRESSO.Outlier-corrected | 228 | -0.007 | (-0.010, -0.004) | -0.006988138 | 0.001399979 | 1.36011E-06 | 1663.399703 | <0.001 | | | | | | | -0.009732097 | -0.00424418 | 241.4800402 |
| LDL | eGFREurStanzick | Lowering LDL | MR.PRESSO.Raw | 228 | -0.007 | (-0.011, -0.003) | -0.006826961 | 0.002120283 | 0.001470404 | 1663.399703 | <0.001 | | | | | | | -0.010982716 | -0.002671206 | 241.4800402 |
| LDL | eGFREurStanzick | Statin | Inverse variance weighted | 5 | -0.013 | (-0.019, -0.007) | -0.012933599 | 0.002960399 | 1.24891E-05 | | | 1.070012511 | 4 | 0.8990004 | 0.000543645 | 0.000911428 | 0.592869028 | -0.018735982 | -0.007131216 | 787.7134762 |
| LDL | eGFREurStanzick | Statin | MR Egger | 5 | -0.022 | (-0.052, 0.008) | -0.02189599 | 0.015314431 | 0.248136817 | | | 0.714229062 | 3 | 0.869852136 | 0.000543645 | 0.000911428 | 0.592869028 | -0.051912275 | 0.008120294 | 787.7134762 |
| LDL | eGFREurStanzick | Statin | MR.PRESSO.Outlier-corrected | 5 | | (NA, NA) | | | | 1.832279852 | 0.886 | | | | | | | | | 787.7134762 |
| LDL | eGFREurStanzick | Statin | MR.PRESSO.Raw | 5 | -0.013 | (-0.016, -0.010) | -0.012933599 | 0.001531139 | 0.001075987 | 1.832279852 | 0.886 | | | | | | | -0.015934632 | -0.009932566 | 787.7134762 |
| LDL | eGFREurStanzick | NPC1L1I | Inverse variance weighted | 6 | -0.02 | (-0.033, -0.008) | -0.02020049 | 0.006362486 | 0.001498693 | | | 6.950275836 | 5 | 0.224364704 | -0.000706666 | 0.00078437 | 0.418563899 | -0.032670963 | -0.007730017 | 191.4711596 |
| LDL | eGFREurStanzick | NPC1L1I | MR Egger | 6 | 0.002 | (-0.048, 0.053) | 0.002394982 | 0.025905105 | 0.930784112 | | | 5.777834438 | 4 | 0.21636544 | -0.000706666 | 0.00078437 | 0.418563899 | -0.048379024 | 0.053168988 | 191.4711596 |
| LDL | eGFREurStanzick | NPC1L1I | MR.PRESSO.Outlier-corrected | 6 | | (NA, NA) | | | | 9.014568156 | 0.335 | | | | | | | | | 191.4711596 |
| LDL | eGFREurStanzick | NPC1L1I | MR.PRESSO.Raw | 6 | -0.02 | (-0.033, -0.008) | -0.02020049 | 0.006362486 | 0.024677894 | 9.014568156 | 0.335 | | | | | | | -0.032670963 | -0.007730017 | 191.4711596 |
| HbA1c | eGFREurStanzick | MG3 | Wald ratio | 1 | -0.015 | (-0.086, 0.057) | -0.014631413 | 0.036464387 | 0.688234501 | | | | | | | | | -0.08610161 | 0.056838785 | 13.3793901 |
| HbA1c | eGFREurStanzick | MC1 | Inverse variance weighted | 25 | 0.022 | (0.001, 0.043) | 0.022143679 | 0.010575737 | 0.036276103 | | | 110.5193823 | 24 | 4.57564E-13 | 4.54176E-05 | 0.000347675 | 0.897202074 | 0.001415235 | 0.042872123 | 30.29781153 |
| HbA1c | eGFREurStanzick | MC1 | MR Egger | 25 | 0.02 | (-0.022, 0.061) | 0.019759194 | 0.021208715 | 0.361192242 | | | 110.4374435 | 23 | 2.1117E-13 | 4.54176E-05 | 0.000347675 | 0.897202074 | -0.021809887 | 0.061328276 | 30.29781153 |
| HbA1c | eGFREurStanzick | MC1 | MR.PRESSO.Outlier-corrected | 25 | 0.021 | (0.006, 0.035) | 0.02069979 | 0.007399572 | 0.010791629 | 127.0761544 | <0.001 | | | | | | | 0.00619663 | 0.035202951 | 30.29781153 |
| HbA1c | eGFREurStanzick | MC1 | MR.PRESSO.Raw | 25 | 0.022 | (0.001, 0.043) | 0.022143679 | 0.010575737 | 0.047017445 | 127.0761544 | <0.001 | | | | | | | 0.001415235 | 0.042872123 | 30.29781153 |
| HbA1c | eGFREurStanzick | Lowering HbA1c | Inverse variance weighted | 281 | 0 | (-0.003, 0.004) | 0.000338697 | 0.001920001 | 0.859975913 | | | 2010.881833 | 280 | 5.6451E-259 | -2.67023E-06 | 0.000108237 | 0.980335699 | -0.003424504 | 0.004101899 | 124.6028832 |
| HbA1c | eGFREurStanzick | Lowering HbA1c | MR Egger | 281 | 0 | (-0.006, 0.007) | 0.000410362 | 0.003483985 | 0.906322544 | | | 2010.877447 | 279 | 2.1036E-259 | -2.67023E-06 | 0.000108237 | 0.980335699 | -0.006418248 | 0.007238973 | 124.6028832 |
| HbA1c | eGFREurStanzick | Lowering HbA1c | MR.PRESSO.Outlier-corrected | 281 | 0 | (-0.002, 0.003) | 0.000235533 | 0.001187455 | 0.842938534 | 2025.002812 | <0.001 | | | | | | | -0.002091879 | 0.002562945 | 124.6028832 |
| HbA1c | eGFREurStanzick | Lowering HbA1c | MR.PRESSO.Raw | 281 | 0 | (-0.003, 0.004) | 0.000338697 | 0.001920001 | 0.860103434 | 2025.002812 | <0.001 | | | | | | | -0.003424504 | 0.004101899 | 124.6028832 |
| HbA1c | eGFREurStanzick | GLP1RA | Inverse variance weighted | 4 | -0.007 | (-0.034, 0.020) | -0.006604113 | 0.01374435 | 0.630874346 | | | 0.418102896 | 3 | 0.936477988 | -2.17752E-05 | 0.000445601 | 0.965466434 | -0.033543038 | 0.020334813 | 23.04119877 |
| HbA1c | eGFREurStanzick | GLP1RA | MR Egger | 4 | -0.005 | (-0.068, 0.057) | -0.005198666 | 0.031876049 | 0.885437203 | | | 0.415714914 | 2 | 0.812322819 | -2.17752E-05 | 0.000445601 | 0.965466434 | -0.067675722 | 0.05727839 | 23.04119877 |
| HbA1c | eGFREurStanzick | GLP1RA | MR.PRESSO.Outlier-corrected | 4 | | (NA, NA) | | | | 0.824417602 | 0.937 | | | | | | | | | 23.04119877 |
| HbA1c | eGFREurStanzick | GLP1RA | MR.PRESSO.Raw | 4 | -0.007 | (-0.017, 0.003) | -0.006604113 | 0.005131037 | 0.288378083 | 0.824417602 | 0.937 | | | | | | | -0.016660946 | 0.00345272 | 23.04119877 |
| HbA1c | eGFREurStanzick | GCG | Wald ratio | 1 | -0.095 | (-0.225, 0.035) | -0.095238095 | 0.066281697 | 0.150755309 | | | | | | | | | -0.225150222 | 0.034674031 | 4.12399181 |
| HbA1c | eGFREurStanzick | AMPK | Inverse variance weighted | 3 | 0.024 | (-0.036, 0.085) | 0.024297152 | 0.030938278 | 0.432252639 | | | 4.784793423 | 2 | 0.091410337 | -0.001512394 | 0.001481002 | 0.493324092 | -0.036341873 | 0.084936178 | 14.59673982 |
| HbA1c | eGFREurStanzick | AMPK | MR Egger | 3 | 0.138 | (-0.088, 0.363) | 0.137575562 | 0.115073654 | 0.443449414 | | | 2.342224692 | 1 | 0.125909624 | -0.001512394 | 0.001481002 | 0.493324092 | -0.0879688 | 0.363119925 | 14.59673982 |
| HbA1c | eGFREurStanzick | GDF15 | Wald ratio | 1 | 0.1 | (0.029, 0.172) | 0.100403112 | 0.03632699 | 0.005712012 | | | | | | | | | 0.029202212 | 0.171604012 | 13.33861351 |
| HbA1c | eGFREurStanzick | SGLT2I | Inverse variance weighted | 6 | 0.028 | (0.008, 0.048) | 0.028074709 | 0.009996771 | 0.004979181 | | | 1.247819432 | 5 | 0.94020831 | 0.000679361 | 0.000990689 | 0.530552635 | 0.008481037 | 0.04766838 | 29.79338183 |
| HbA1c | eGFREurStanzick | SGLT2I | MR Egger | 6 | -0.02 | (-0.157, 0.118) | -0.019589006 | 0.070221567 | 0.79410407 | | | 0.777571624 | 4 | 0.94142936 | 0.000679361 | 0.000990689 | 0.530552635 | -0.157223277 | 0.118045265 | 29.79338183 |
| HbA1c | eGFREurStanzick | SGLT2I | MR.PRESSO.Outlier-corrected | 6 | | (NA, NA) | | | | 2.070126299 | 0.946 | | | | | | | | | 29.79338183 |
| HbA1c | eGFREurStanzick | SGLT2I | MR.PRESSO.Raw | 6 | 0.028 | (0.018, 0.038) | 0.028074709 | 0.004994024 | 0.002465869 | 2.070126299 | 0.946 | | | | | | | 0.018286422 | 0.037862996 | 29.79338183 |

### Table S5. The effect of genetically proxied antihypertensives, lipid-lowering, and antidiabetic drugs on cystatin-based cross-sectional eGFR

| IntermediatePheno | outcome | exposure | method | nsnp | Beta | 95%CI | b | se | pval | RSSobs | Phet | Q | Q_df | Q_pval | egger_intercept | egger_se | egger_pval | Lower95 | Upper95 | MeanFstat |
|---|---|---|---|---|---|---|---|---|---|---|---|---|---|---|---|---|---|---|---|---|
| SBP | eGFRCysStanzick | ACEI | Wald ratio | 1 | 0.032 | (-0.006, 0.069) | 0.031701303 | 0.019052483 | 0.096133712 | | | | | | | | | -0.005641564 | 0.06904417 | 82.79833374 |
| SBP | eGFRCysStanzick | CCB | Inverse variance weighted | 24 | 0.001 | (-0.008, 0.010) | 0.001010451 | 0.004542302 | 0.823960827 | | | 28.02521559 | 23 | 0.21482959 | -0.001417588 | 0.003706179 | 0.7057659 | -0.00789246 | 0.009913362 | 66.21360566 |
| SBP | eGFRCysStanzick | CCB | MR Egger | 24 | 0.005 | (-0.017, 0.027) | 0.004975774 | 0.011353564 | 0.665473542 | | | 27.8400778 | 22 | 0.181041893 | -0.001417588 | 0.003706179 | 0.7057659 | -0.017277211 | 0.027228758 | 66.21360566 |
| SBP | eGFRCysStanzick | CCB | MR.PRESSO.Outlier-corrected | 24 | | (NA, NA) | | | | 30.1933562 | 0.225 | | | | | | | | | 66.21360566 |
| SBP | eGFRCysStanzick | CCB | MR.PRESSO.Raw | 24 | 0.001 | (-0.008, 0.010) | 0.001010451 | 0.004542302 | 0.825924396 | 30.1933562 | 0.225 | | | | | | | -0.00789246 | 0.009913362 | 66.21360566 |
| SBP | eGFRCysStanzick | BB | Inverse variance weighted | 5 | -0.018 | (-0.044, 0.009) | -0.017887002 | 0.013533007 | 0.186257587 | | | 6.778861431 | 4 | 0.148046158 | 0.001640767 | 0.019595861 | 0.938545136 | -0.044411696 | 0.008637693 | 52.2808789 |
| SBP | eGFRCysStanzick | BB | MR Egger | 5 | -0.023 | (-0.146, 0.100) | -0.022968836 | 0.062667785 | 0.738303287 | | | 6.763056713 | 3 | 0.079845936 | 0.001640767 | 0.019595861 | 0.938545136 | -0.145797696 | 0.099860023 | 52.2808789 |
| SBP | eGFRCysStanzick | BB | MR.PRESSO.Outlier-corrected | 5 | | (NA, NA) | | | | 11.0068961 | 0.193 | | | | | | | | | 52.2808789 |
| SBP | eGFRCysStanzick | BB | MR.PRESSO.Raw | 5 | -0.018 | (-0.044, 0.009) | -0.017887002 | 0.013533007 | 0.256781562 | 11.0068961 | 0.193 | | | | | | | -0.044411696 | 0.008637693 | 52.2808789 |
| SBP | eGFRCysStanzick | Lowering SBP | Inverse variance weighted | 439 | 0.001 | (-0.004, 0.006) | 0.000924477 | 0.002386536 | 0.698481013 | | | 2925.729903 | 438 | 0 | 0.000184344 | 0.001844436 | 0.92043291 | -0.003753134 | 0.005602087 | 74.77613976 |
| SBP | eGFRCysStanzick | Lowering SBP | MR Egger | 439 | 0 | (-0.011, 0.012) | 0.000376135 | 0.005984042 | 0.949909681 | | | 2925.663027 | 437 | 0 | 0.000184344 | 0.001844436 | 0.92043291 | -0.011352587 | 0.012104856 | 74.77613976 |
| SBP | eGFRCysStanzick | Lowering SBP | MR.PRESSO.Outlier-corrected | 439 | 0.001 | (-0.002, 0.004) | 0.000883778 | 0.001413395 | 0.532144604 | 2941.320798 | <0.001 | | | | | | | -0.001886477 | 0.003654032 | 74.77613976 |
| SBP | eGFRCysStanzick | Lowering SBP | MR.PRESSO.Raw | 439 | 0.001 | (-0.004, 0.006) | 0.000924477 | 0.002386536 | 0.698669174 | 2941.320798 | <0.001 | | | | | | | -0.003753134 | 0.005602087 | 74.77613976 |
| LDL | eGFRCysStanzick | PCSK9I | Inverse variance weighted | 11 | 0.002 | (-0.005, 0.008) | 0.001508062 | 0.003111332 | 0.627889268 | | | 9.338906426 | 10 | 0.500270366 | 0.000584043 | 0.000310341 | 0.092519406 | -0.004590148 | 0.007606272 | 869.0826758 |
| LDL | eGFRCysStanzick | PCSK9I | MR Egger | 11 | -0.005 | (-0.015, 0.004) | -0.005242754 | 0.004748483 | 0.298197024 | | | 5.797208755 | 9 | 0.760030436 | 0.000584043 | 0.000310341 | 0.092519406 | -0.01454978 | 0.004064272 | 869.0826758 |
| LDL | eGFRCysStanzick | PCSK9I | MR.PRESSO.Outlier-corrected | 11 | | (NA, NA) | | | | 14.69328858 | 0.435 | | | | | | | | | 869.0826758 |
| LDL | eGFRCysStanzick | PCSK9I | MR.PRESSO.Raw | 11 | 0.002 | (-0.004, 0.007) | 0.001508062 | 0.003006729 | 0.626832644 | 14.69328858 | 0.435 | | | | | | | -0.004385127 | 0.007401251 | 869.0826758 |
| LDL | eGFRCysStanzick | Lowering LDL | Inverse variance weighted | 221 | -0.006 | (-0.013, 0.001) | -0.006146751 | 0.003408402 | 0.071323651 | | | 1481.241315 | 220 | 1.1168E-185 | -0.000163019 | 0.000162944 | 0.318193442 | -0.01282722 | 0.000533717 | 242.8748359 |
| LDL | eGFRCysStanzick | Lowering LDL | MR Egger | 221 | -0.002 | (-0.012, 0.007) | -0.00243323 | 0.005039326 | 0.629685791 | | | 1474.502271 | 219 | 7.6012E-185 | -0.000163019 | 0.000162944 | 0.318193442 | -0.012310308 | 0.007443848 | 242.8748359 |
| LDL | eGFRCysStanzick | Lowering LDL | MR.PRESSO.Outlier-corrected | 221 | -0.004 | (-0.008, -0.000) | -0.004114931 | 0.002019606 | 0.04295173 | 1492.562867 | <0.001 | | | | | | | -0.008073359 | -0.000156503 | 242.8748359 |
| LDL | eGFRCysStanzick | Lowering LDL | MR.PRESSO.Raw | 221 | -0.006 | (-0.013, 0.001) | -0.006146751 | 0.003408402 | 0.072691799 | 1492.562867 | <0.001 | | | | | | | -0.01282722 | 0.000533717 | 242.8748359 |
| LDL | eGFRCysStanzick | Statin | Inverse variance weighted | 5 | -0.013 | (-0.023, -0.004) | -0.013334779 | 0.004994026 | 0.007581818 | | | 2.713997929 | 4 | 0.606767386 | -0.001087337 | 0.001558661 | 0.535626559 | -0.023123069 | -0.003546489 | 787.7134762 |
| LDL | eGFRCysStanzick | Statin | MR Egger | 5 | 0.005 | (-0.047, 0.056) | 0.004542406 | 0.026108421 | 0.872957181 | | | 2.227338533 | 3 | 0.526583626 | -0.001087337 | 0.001558661 | 0.535626559 | -0.046630098 | 0.055714911 | 787.7134762 |
| LDL | eGFRCysStanzick | Statin | MR.PRESSO.Outlier-corrected | 5 | | (NA, NA) | | | | 3.533759086 | 0.672 | | | | | | | | | 787.7134762 |
| LDL | eGFRCysStanzick | Statin | MR.PRESSO.Raw | 5 | -0.013 | (-0.021, -0.005) | -0.013334779 | 0.004113633 | 0.031625822 | 3.533759086 | 0.672 | | | | | | | -0.021397499 | -0.005272059 | 787.7134762 |
| LDL | eGFRCysStanzick | NPC1L1I | Inverse variance weighted | 6 | 0.008 | (-0.010, 0.025) | 0.007540224 | 0.009107216 | 0.407704726 | | | 0.296579617 | 5 | 0.99770692 | 1.03804E-05 | 0.001068816 | 0.992716085 | -0.010309919 | 0.025390367 | 191.4711596 |
| LDL | eGFRCysStanzick | NPC1L1I | MR Egger | 6 | 0.007 | (-0.062, 0.077) | 0.007206416 | 0.035556503 | 0.849280665 | | | 0.296485292 | 4 | 0.990039927 | 1.03804E-05 | 0.001068816 | 0.992716085 | -0.06248433 | 0.076897163 | 191.4711596 |
| LDL | eGFRCysStanzick | NPC1L1I | MR.PRESSO.Outlier-corrected | 6 | | (NA, NA) | | | | 0.394215383 | 0.997 | | | | | | | | | 191.4711596 |
| LDL | eGFRCysStanzick | NPC1L1I | MR.PRESSO.Raw | 6 | 0.008 | (0.003, 0.012) | 0.007540224 | 0.00221805 | 0.019261832 | 0.394215383 | 0.997 | | | | | | | 0.003192847 | 0.011887602 | 191.4711596 |
| HbA1c | eGFRCysStanzick | MG3 | Wald ratio | 1 | 0.083 | (-0.027, 0.193) | 0.083015109 | 0.05612859 | 0.139135917 | | | | | | | | | -0.026996928 | 0.193027146 | 13.3793901 |
| HbA1c | eGFRCysStanzick | MC1 | Inverse variance weighted | 24 | 0.023 | (-0.007, 0.053) | 0.022780778 | 0.015375758 | 0.138445814 | | | 90.68113032 | 23 | 5.46183E-10 | 0.000463654 | 0.000500637 | 0.364427626 | -0.007355708 | 0.052917264 | 31.06201727 |
| HbA1c | eGFRCysStanzick | MC1 | MR Egger | 24 | -0.001 | (-0.059, 0.058) | -0.00079153 | 0.029760973 | 0.979021558 | | | 87.27840188 | 22 | 9.91314E-10 | 0.000463654 | 0.000500637 | 0.364427626 | -0.059123037 | 0.057539978 | 31.06201727 |
| HbA1c | eGFRCysStanzick | MC1 | MR.PRESSO.Outlier-corrected | 24 | 0.014 | (-0.010, 0.039) | 0.014447118 | 0.012613004 | 0.264345225 | 96.86382397 | <0.001 | | | | | | | -0.010274369 | 0.039168605 | 31.06201727 |
| HbA1c | eGFRCysStanzick | MC1 | MR.PRESSO.Raw | 24 | 0.023 | (-0.007, 0.053) | 0.022780778 | 0.015375758 | 0.152017362 | 96.86382397 | <0.001 | | | | | | | -0.007355708 | 0.052917264 | 31.06201727 |
| HbA1c | eGFRCysStanzick | Lowering HbA1c | Inverse variance weighted | 273 | -0.003 | (-0.008, 0.002) | -0.003380206 | 0.002554746 | 0.185799358 | | | 1270.604164 | 272 | 1.4718E-128 | 0.000139475 | 0.000145289 | 0.337920328 | -0.008387509 | 0.001627097 | 124.7585211 |
| HbA1c | eGFRCysStanzick | Lowering HbA1c | MR Egger | 273 | -0.007 | (-0.016, 0.002) | -0.00706379 | 0.004610018 | 0.126622452 | | | 1266.297984 | 271 | 3.7038E-128 | 0.000139475 | 0.000145289 | 0.337920328 | -0.016099426 | 0.001971845 | 124.7585211 |
| HbA1c | eGFRCysStanzick | Lowering HbA1c | MR.PRESSO.Outlier-corrected | 273 | -0.005 | (-0.009, -0.001) | -0.005006828 | 0.001874775 | 0.008071714 | 1278.094731 | <0.001 | | | | | | | -0.008681387 | -0.001332269 | 124.7585211 |
| HbA1c | eGFRCysStanzick | Lowering HbA1c | MR.PRESSO.Raw | 273 | -0.003 | (-0.008, 0.002) | -0.003380206 | 0.002554746 | 0.186910529 | 1278.094731 | <0.001 | | | | | | | -0.008387509 | 0.001627097 | 124.7585211 |
| HbA1c | eGFRCysStanzick | GLP1RA | Inverse variance weighted | 4 | -0.022 | (-0.067, 0.024) | -0.02164224 | 0.023124696 | 0.349328298 | | | 2.415682202 | 3 | 0.490722062 | 0.000867149 | 0.000766592 | 0.375371446 | -0.066966644 | 0.023682164 | 23.04119877 |
| HbA1c | eGFRCysStanzick | GLP1RA | MR Egger | 4 | -0.076 | (-0.180, 0.028) | -0.075909074 | 0.053256452 | 0.290125532 | | | 1.136129004 | 2 | 0.566621072 | 0.000867149 | 0.000766592 | 0.375371446 | -0.18029172 | 0.028473571 | 23.04119877 |
| HbA1c | eGFRCysStanzick | GLP1RA | MR.PRESSO.Outlier-corrected | 4 | | (NA, NA) | | | | 3.490994538 | 0.59 | | | | | | | | | 23.04119877 |
| HbA1c | eGFRCysStanzick | GLP1RA | MR.PRESSO.Raw | 4 | -0.022 | (-0.062, 0.019) | -0.02164224 | 0.020750822 | 0.373617856 | 3.490994538 | 0.59 | | | | | | | -0.062313851 | 0.019029371 | 23.04119877 |
| HbA1c | eGFRCysStanzick | GCG | Wald ratio | 1 | 0.08 | (-0.125, 0.284) | 0.079504561 | 0.10410913 | 0.445066549 | | | | | | | | | -0.124549335 | 0.283558457 | 4.12399181 |
| HbA1c | eGFRCysStanzick | AMPK | Inverse variance weighted | 3 | 0.052 | (-0.016, 0.119) | 0.051746774 | 0.034359108 | 0.13205255 | | | 1.219261434 | 2 | 0.543551556 | -0.001465238 | 0.001656067 | 0.538872938 | -0.015597078 | 0.119090625 | 14.59673982 |
| HbA1c | eGFRCysStanzick | AMPK | MR Egger | 3 | 0.162 | (-0.092, 0.416) | 0.162131063 | 0.129405241 | 0.428835438 | | | 0.436443597 | 1 | 0.508843993 | -0.001465238 | 0.001656067 | 0.538872938 | -0.09150321 | 0.415765335 | 14.59673982 |
| HbA1c | eGFRCysStanzick | GDF15 | Wald ratio | 1 | 0.103 | (-0.016, 0.222) | 0.103121777 | 0.060851223 | 0.090141051 | | | | | | | | | -0.01614662 | 0.222390175 | 13.33861351 |
| HbA1c | eGFRCysStanzick | SGLT2I | Inverse variance weighted | 5 | 0.01 | (-0.025, 0.046) | 0.010398512 | 0.018078944 | 0.565174616 | | | 0.463870286 | 4 | 0.976921554 | 0.000117843 | 0.001700955 | 0.949125927 | -0.025036218 | 0.045833242 | 30.16136229 |
| HbA1c | eGFRCysStanzick | SGLT2I | MR Egger | 5 | 0.002 | (-0.231, 0.236) | 0.00224088 | 0.119128035 | 0.986173257 | | | 0.459070525 | 3 | 0.927786899 | 0.000117843 | 0.001700955 | 0.949125927 | -0.231250069 | 0.235731829 | 30.16136229 |
| HbA1c | eGFRCysStanzick | SGLT2I | MR.PRESSO.Outlier-corrected | 5 | | (NA, NA) | | | | 0.805019534 | 0.982 | | | | | | | | | 30.16136229 |
| HbA1c | eGFRCysStanzick | SGLT2I | MR.PRESSO.Raw | 5 | 0.01 | (-0.002, 0.022) | 0.010398512 | 0.006156606 | 0.166487852 | 0.805019534 | 0.982 | | | | | | | -0.001668435 | 0.022465459 | 30.16136229 |

### Table S6. The effect of genetically proxied antihypertensives, lipid-lowering, and antidiabetic drugs on BUN

| IntermediatePheno | outcome | exposure | method | nsnp | Beta | 95%CI | b | se | pval | RSSobs | Phet | Q | Q_df | Q_pval | egger_intercept | egger_se | egger_pval | Lower95 | Upper95 | MeanFstat |
|---|---|---|---|---|---|---|---|---|---|---|---|---|---|---|---|---|---|---|---|---|
| SBP | BUNStanzick | ACEI | Wald ratio | 1 | -0.025 | (-0.062, 0.013) | -0.024656569 | 0.019263825 | 0.200565715 | | | | | | | | | -0.062413667 | 0.013100528 | 82.79833374 |
| SBP | BUNStanzick | CCB | Inverse variance weighted | 24 | 0.004 | (-0.005, 0.013) | 0.004218691 | 0.004544967 | 0.353297795 | | | 16.15963816 | 23 | 0.848350066 | 0.0015927 | 0.00361312 | 0.663651772 | -0.004689444 | 0.013126827 | 66.21360566 |
| SBP | BUNStanzick | CCB | MR Egger | 24 | 0 | (-0.023, 0.022) | -0.000442108 | 0.011508715 | 0.96970322 | | | 15.96532457 | 22 | 0.817603023 | 0.0015927 | 0.00361312 | 0.663651772 | -0.022999188 | 0.022114973 | 66.21360566 |
| SBP | BUNStanzick | CCB | MR.PRESSO.Outlier-corrected | 24 | | (NA, NA) | | | | 17.52099569 | 0.844 | | | | | | | | | 66.21360566 |
| SBP | BUNStanzick | CCB | MR.PRESSO.Raw | 24 | 0.004 | (-0.003, 0.012) | 0.004218691 | 0.003809629 | 0.279578611 | 17.52099569 | 0.844 | | | | | | | -0.003248181 | 0.011685564 | 66.21360566 |
| SBP | BUNStanzick | BB | Inverse variance weighted | 5 | -0.02 | (-0.042, 0.002) | -0.020234484 | 0.011266512 | 0.072496946 | | | 4.184394569 | 4 | 0.381625564 | 0.00056026 | 0.01652193 | 0.975078881 | -0.042316848 | 0.00184788 | 52.2808789 |
| SBP | BUNStanzick | BB | MR Egger | 5 | -0.022 | (-0.127, 0.083) | -0.022000782 | 0.053687108 | 0.709460772 | | | 4.182791313 | 3 | 0.242390458 | 0.00056026 | 0.01652193 | 0.975078881 | -0.127227513 | 0.08322595 | 52.2808789 |
| SBP | BUNStanzick | BB | MR.PRESSO.Outlier-corrected | 5 | | (NA, NA) | | | | 6.539678622 | 0.435 | | | | | | | | | 52.2808789 |
| SBP | BUNStanzick | BB | MR.PRESSO.Raw | 5 | -0.02 | (-0.042, 0.002) | -0.020234484 | 0.011266512 | 0.146923339 | 6.539678622 | 0.435 | | | | | | | -0.042316848 | 0.00184788 | 52.2808789 |
| SBP | BUNStanzick | Lowering SBP | Inverse variance weighted | 436 | -0.008 | (-0.013, -0.003) | -0.007792159 | 0.002546794 | 0.002216358 | | | 2916.012233 | 435 | 0 | 0.0031243 | 0.001959424 | 0.111552181 | -0.012783875 | -0.002800444 | 75.12872998 |
| SBP | BUNStanzick | Lowering SBP | MR Egger | 436 | -0.017 | (-0.030, -0.005) | -0.017171117 | 0.006407963 | 0.007650148 | | | 2899.029344 | 434 | 0 | 0.0031243 | 0.001959424 | 0.111552181 | -0.029730726 | -0.004611509 | 75.12872998 |
| SBP | BUNStanzick | Lowering SBP | MR.PRESSO.Outlier-corrected | 436 | -0.007 | (-0.009, -0.004) | -0.006586152 | 0.001463512 | 8.9177E-06 | 2932.657666 | <0.001 | | | | | | | -0.009454636 | -0.003717668 | 75.12872998 |
| SBP | BUNStanzick | Lowering SBP | MR.PRESSO.Raw | 436 | -0.008 | (-0.013, -0.003) | -0.007792159 | 0.002546794 | 0.002353407 | 2932.657666 | <0.001 | | | | | | | -0.012783875 | -0.002800444 | 75.12872998 |
| LDL | BUNStanzick | PCSK9I | Inverse variance weighted | 11 | -0.004 | (-0.014, 0.005) | -0.004473706 | 0.004980479 | 0.369053288 | | | 20.11726758 | 10 | 0.028162842 | -0.000975887 | 0.000399757 | 0.037291287 | -0.014235445 | 0.005288033 | 869.0826758 |
| LDL | BUNStanzick | PCSK9I | MR Egger | 11 | 0.008 | (-0.005, 0.021) | 0.008274616 | 0.006622126 | 0.24298436 | | | 12.10308131 | 9 | 0.207560273 | -0.000975887 | 0.000399757 | 0.037291287 | -0.00470475 | 0.021253983 | 869.0826758 |
| LDL | BUNStanzick | PCSK9I | MR.PRESSO.Outlier-corrected | 11 | | (NA, NA) | | | | 27.97913219 | 0.062 | | | | | | | | | 869.0826758 |
| LDL | BUNStanzick | PCSK9I | MR.PRESSO.Raw | 11 | -0.004 | (-0.014, 0.005) | -0.004473706 | 0.004980479 | 0.390168286 | 27.97913219 | 0.062 | | | | | | | -0.014235445 | 0.005288033 | 869.0826758 |
| LDL | BUNStanzick | Lowering LDL | Inverse variance weighted | 220 | 0.001 | (-0.005, 0.008) | 0.001435421 | 0.003410944 | 0.673880682 | | | 1242.918203 | 219 | 1.3547E-142 | 8.54798E-05 | 0.00016287 | 0.600230589 | -0.005250029 | 0.00812087 | 241.12835 |
| LDL | BUNStanzick | Lowering LDL | MR Egger | 220 | -0.001 | (-0.011, 0.010) | -0.00062105 | 0.005198685 | 0.905018632 | | | 1241.349703 | 218 | 1.0825E-142 | 8.54798E-05 | 0.00016287 | 0.600230589 | -0.010810472 | 0.009568372 | 241.12835 |
| LDL | BUNStanzick | Lowering LDL | MR.PRESSO.Outlier-corrected | 220 | 0.002 | (-0.003, 0.006) | 0.001863111 | 0.002318186 | 0.422542695 | 1255.157961 | <0.001 | | | | | | | -0.002680533 | 0.006406755 | 241.12835 |
| LDL | BUNStanzick | Lowering LDL | MR.PRESSO.Raw | 220 | 0.001 | (-0.005, 0.008) | 0.001435421 | 0.003410944 | 0.674293357 | 1255.157961 | <0.001 | | | | | | | -0.005250029 | 0.00812087 | 241.12835 |
| LDL | BUNStanzick | Statin | Inverse variance weighted | 5 | 0.01 | (-0.000, 0.020) | 0.009774777 | 0.005104863 | 0.055518373 | | | 1.142198374 | 4 | 0.88752047 | -0.000421584 | 0.001579884 | 0.806879773 | -0.000230754 | 0.019780307 | 787.7134762 |
| LDL | BUNStanzick | Statin | MR Egger | 5 | 0.017 | (-0.035, 0.068) | 0.016691942 | 0.026419881 | 0.572381659 | | | 1.070991978 | 3 | 0.784080943 | -0.000421584 | 0.001579884 | 0.806879773 | -0.035091025 | 0.068474909 | 787.7134762 |
| LDL | BUNStanzick | Statin | MR.PRESSO.Outlier-corrected | 5 | | (NA, NA) | | | | 1.59880246 | 0.918 | | | | | | | | | 787.7134762 |
| LDL | BUNStanzick | Statin | MR.PRESSO.Raw | 5 | 0.01 | (0.004, 0.015) | 0.009774777 | 0.002727877 | 0.023099703 | 1.59880246 | 0.918 | | | | | | | 0.004428137 | 0.015121416 | 787.7134762 |
| LDL | BUNStanzick | NPC1L1I | Inverse variance weighted | 6 | 0.009 | (-0.010, 0.027) | 0.008747882 | 0.009403489 | 0.352225909 | | | 4.102102774 | 5 | 0.534811364 | 0.00145857 | 0.00117399 | 0.281948676 | -0.009682957 | 0.027178721 | 191.4711596 |
| LDL | BUNStanzick | NPC1L1I | MR Egger | 6 | -0.037 | (-0.112, 0.038) | -0.037114242 | 0.038092933 | 0.385073444 | | | 2.558536467 | 4 | 0.634185493 | 0.00145857 | 0.00117399 | 0.281948676 | -0.111776391 | 0.037547908 | 191.4711596 |
| LDL | BUNStanzick | NPC1L1I | MR.PRESSO.Outlier-corrected | 6 | | (NA, NA) | | | | 7.170223812 | 0.507 | | | | | | | | | 191.4711596 |
| LDL | BUNStanzick | NPC1L1I | MR.PRESSO.Raw | 6 | 0.009 | (-0.008, 0.025) | 0.008747882 | 0.008517405 | 0.351488955 | 7.170223812 | 0.507 | | | | | | | -0.007946232 | 0.025441996 | 191.4711596 |
| HbA1c | BUNStanzick | MG3 | Wald ratio | 1 | -0.125 | (-0.258, 0.009) | -0.124522663 | 0.068093143 | 0.06744295 | | | | | | | | | -0.257985223 | 0.008939897 | 13.3793901 |
| HbA1c | BUNStanzick | MC1 | Inverse variance weighted | 24 | -0.011 | (-0.032, 0.010) | -0.010898995 | 0.010604314 | 0.304049163 | | | 34.125684 | 23 | 0.063362576 | -0.00031 | 0.00034262 | 0.375377751 | -0.03168345 | 0.009885459 | 31.06201727 |
| HbA1c | BUNStanzick | MC1 | MR Egger | 24 | 0.006 | (-0.036, 0.047) | 0.005725609 | 0.021235516 | 0.789962205 | | | 32.90138051 | 22 | 0.063274613 | -0.00031 | 0.00034262 | 0.375377751 | -0.035896003 | 0.047347221 | 31.06201727 |
| HbA1c | BUNStanzick | MC1 | MR.PRESSO.Outlier-corrected | 24 | | (NA, NA) | | | | 37.30667474 | 0.088 | | | | | | | | | 31.06201727 |
| HbA1c | BUNStanzick | MC1 | MR.PRESSO.Raw | 24 | -0.011 | (-0.032, 0.010) | -0.010898995 | 0.010604314 | 0.314737455 | 37.30667474 | 0.088 | | | | | | | -0.03168345 | 0.009885459 | 31.06201727 |
| HbA1c | BUNStanzick | Lowering HbA1c | Inverse variance weighted | 274 | -0.002 | (-0.007, 0.004) | -0.001781631 | 0.002854121 | 0.532475715 | | | 1356.613211 | 273 | 4.7184E-143 | -0.000273604 | 0.000160443 | 0.089280485 | -0.007375708 | 0.003812445 | 124.4214904 |
| HbA1c | BUNStanzick | Lowering HbA1c | MR Egger | 274 | 0.006 | (-0.005, 0.016) | 0.005622706 | 0.005190576 | 0.279655593 | | | 1342.262636 | 272 | 6.5749E-141 | -0.000273604 | 0.000160443 | 0.089280485 | -0.004550824 | 0.015796235 | 124.4214904 |
| HbA1c | BUNStanzick | Lowering HbA1c | MR.PRESSO.Outlier-corrected | 274 | -0.003 | (-0.006, 0.001) | -0.002622199 | 0.001781856 | 0.14236869 | 1367.65536 | <0.001 | | | | | | | -0.006114636 | 0.000870239 | 124.4214904 |
| HbA1c | BUNStanzick | Lowering HbA1c | MR.PRESSO.Raw | 274 | -0.002 | (-0.007, 0.004) | -0.001781631 | 0.002854121 | 0.532997005 | 1367.65536 | <0.001 | | | | | | | -0.007375708 | 0.003812445 | 124.4214904 |
| HbA1c | BUNStanzick | GLP1RA | Inverse variance weighted | 4 | -0.079 | (-0.162, 0.004) | -0.079279641 | 0.042376802 | 0.061369145 | | | 9.309484705 | 3 | 0.025446928 | -0.002207556 | 0.000777887 | 0.104976959 | -0.162338174 | 0.003778892 | 23.04119877 |
| HbA1c | BUNStanzick | GLP1RA | MR Egger | 4 | 0.06 | (-0.047, 0.168) | 0.060235039 | 0.054731609 | 0.385847599 | | | 1.255886807 | 2 | 0.533688254 | -0.002207556 | 0.000777887 | 0.104976959 | -0.047038914 | 0.167508992 | 23.04119877 |
| HbA1c | BUNStanzick | GLP1RA | MR.PRESSO.Outlier-corrected | 4 | | (NA, NA) | | | | 16.05310432 | 0.126 | | | | | | | | | 23.04119877 |
| HbA1c | BUNStanzick | GLP1RA | MR.PRESSO.Raw | 4 | -0.079 | (-0.162, 0.004) | -0.079279641 | 0.042376802 | 0.158124026 | 16.05310432 | 0.126 | | | | | | | -0.162338174 | 0.003778892 | 23.04119877 |
| HbA1c | BUNStanzick | GCG | Wald ratio | 1 | 0.046 | (-0.196, 0.288) | 0.046028956 | 0.123566826 | 0.709518721 | | | | | | | | | -0.196162022 | 0.288219935 | 4.12399181 |
| HbA1c | BUNStanzick | AMPK | Inverse variance weighted | 3 | 0.061 | (-0.098, 0.221) | 0.061280742 | 0.081478961 | 0.451987855 | | | 11.37054316 | 2 | 0.003395611 | 0.004381796 | 0.003364152 | 0.416838674 | -0.098418021 | 0.220979506 | 14.59673982 |
| HbA1c | BUNStanzick | AMPK | MR Egger | 3 | -0.272 | (-0.793, 0.248) | -0.272259515 | 0.265517978 | 0.492019802 | | | 4.216782749 | 1 | 0.040025985 | 0.004381796 | 0.003364152 | 0.416838674 | -0.792674753 | 0.248155722 | 14.59673982 |
| HbA1c | BUNStanzick | GDF15 | Wald ratio | 1 | 0.009 | (-0.111, 0.130) | 0.009374707 | 0.061516828 | 0.878877322 | | | | | | | | | -0.111198275 | 0.129947689 | 13.33861351 |
| HbA1c | BUNStanzick | SGLT2I | Inverse variance weighted | 6 | -0.018 | (-0.055, 0.019) | -0.018307361 | 0.018968613 | 0.33447486 | | | 5.956478501 | 5 | 0.310476815 | -0.000382267 | 0.002016388 | 0.858869529 | -0.055485841 | 0.01887112 | 29.79338183 |
| HbA1c | BUNStanzick | SGLT2I | MR Egger | 6 | 0.009 | (-0.272, 0.289) | 0.008567778 | 0.143324893 | 0.955199309 | | | 5.903435171 | 4 | 0.20647679 | -0.000382267 | 0.002016388 | 0.858869529 | -0.272349012 | 0.289484568 | 29.79338183 |
| HbA1c | BUNStanzick | SGLT2I | MR.PRESSO.Outlier-corrected | 6 | | (NA, NA) | | | | 8.596572683 | 0.357 | | | | | | | | | 29.79338183 |
| HbA1c | BUNStanzick | SGLT2I | MR.PRESSO.Raw | 6 | -0.018 | (-0.055, 0.019) | -0.018307361 | 0.018968613 | 0.378801637 | 8.596572683 | 0.357 | | | | | | | -0.055485841 | 0.01887112 | 29.79338183 |

#### Table S7. Steiger test of directionality

| id.exposure | id.outcome | exposure | outcome | snp_r2.exposure | snp_r2.outcome | correct_causal_direction | steiger_pval | IntermediatePheno |
|---|---|---|---|---|---|---|---|---|
| 9rRHy9 | LUmXh9 | PCSK9I | OveralleGFRDeclAdjDM | 0.005963165 | 4.56E-05 | TRUE | 1.93E-244 | LDL |
| D94SpL | LUmXh9 | ACEI | OveralleGFRDeclAdjDM | 1.14E-04 | 7.10E-07 | TRUE | 4.79E-06 | SBP |
| f3yEFg | LUmXh9 | CCB | OveralleGFRDeclAdjDM | 0.002247725 | 7.29E-05 | TRUE | 6.48E-73 | SBP |
| fhrnCj | LUmXh9 | MG3 | OveralleGFRDeclAdjDM | 3.89E-05 | 4.77E-06 | TRUE | 0.102730004 | HbA1c |
| Fq9tX7 | LUmXh9 | Lowering LDL | OveralleGFRDeclAdjDM | 0.042086443 | 8.00E-04 | TRUE | 0 | LDL |
| fwCnU5 | LUmXh9 | Statin | OveralleGFRDeclAdjDM | 0.00301332 | 9.04E-06 | TRUE | 2.43E-135 | LDL |
| IaocOm | LUmXh9 | MC1 | OveralleGFRDeclAdjDM | 0.002199959 | 9.90E-05 | TRUE | 5.31E-49 | HbA1c |
| IBfVDL | LUmXh9 | BB | OveralleGFRDeclAdjDM | 3.65E-04 | 1.84E-05 | TRUE | 2.36E-11 | SBP |
| JWMMYI | LUmXh9 | Lowering HbA1c | OveralleGFRDeclAdjDM | 0.100508917 | 0.001160243 | TRUE | 0 | HbA1c |
| puxRXK | LUmXh9 | GLP1RA | OveralleGFRDeclAdjDM | 2.68E-04 | 1.66E-06 | TRUE | 9.72E-10 | HbA1c |
| rXdiei | LUmXh9 | Lowering SBP | OveralleGFRDeclAdjDM | 0.047501093 | 0.002302333 | TRUE | 0 | SBP |
| Sr1HqQ | LUmXh9 | NPC1L1I | OveralleGFRDeclAdjDM | 0.001086372 | 3.24E-06 | TRUE | 4.73E-52 | LDL |
| vxLdAw | LUmXh9 | GCG | OveralleGFRDeclAdjDM | 1.20E-05 | 2.00E-05 | FALSE | 0.701965626 | HbA1c |
| Vyjxei | LUmXh9 | AMPK | OveralleGFRDeclAdjDM | 1.27E-04 | 1.18E-05 | TRUE | 0.001286691 | HbA1c |
| WbqWq0 | LUmXh9 | GDF15 | OveralleGFRDeclAdjDM | 3.88E-05 | 3.13E-06 | TRUE | 0.066969906 | HbA1c |
| Xjf1jJ | LUmXh9 | SGLT2I | OveralleGFRDeclAdjDM | 5.19E-04 | 2.72E-05 | TRUE | 1.51E-12 | HbA1c |
| 9rRHy9 | bhH1Up | PCSK9I | eGFRCreatStanzick | 0.005963165 | 4.26E-05 | TRUE | 0 | LDL |
| D94SpL | bhH1Up | ACEI | eGFRCreatStanzick | 1.14E-04 | 3.65E-06 | TRUE | 4.48E-09 | SBP |
| f3yEFg | bhH1Up | CCB | eGFRCreatStanzick | 0.002247725 | 3.25E-05 | TRUE | 3.53E-164 | SBP |
| fhrnCj | bhH1Up | MG3 | eGFRCreatStanzick | 3.89E-05 | 5.22E-09 | TRUE | 0.001499736 | HbA1c |
| Fq9tX7 | bhH1Up | Lowering LDL | eGFRCreatStanzick | 0.041773377 | 0.001602779 | TRUE | 0 | LDL |
| fwCnU5 | bhH1Up | Statin | eGFRCreatStanzick | 0.00301332 | 1.88E-05 | TRUE | 4.44E-273 | LDL |
| IaocOm | bhH1Up | MC1 | eGFRCreatStanzick | 0.002165219 | 1.17E-04 | TRUE | 1.96E-74 | HbA1c |
| IBfVDL | bhH1Up | BB | eGFRCreatStanzick | 3.65E-04 | 7.86E-06 | TRUE | 1.82E-26 | SBP |
| JWMMYI | bhH1Up | Lowering HbA1c | eGFRCreatStanzick | 0.097507451 | 0.001843754 | TRUE | 0 | HbA1c |
| puxRXK | bhH1Up | GLP1RA | eGFRCreatStanzick | 2.68E-04 | 7.82E-07 | TRUE | 1.86E-15 | HbA1c |
| rXdiei | bhH1Up | Lowering SBP | eGFRCreatStanzick | 0.047501093 | 0.004994802 | TRUE | 0 | SBP |
| Sr1HqQ | bhH1Up | NPC1L1I | eGFRCreatStanzick | 0.001086372 | 1.75E-05 | TRUE | 7.88E-89 | LDL |
| vxLdAw | bhH1Up | GCG | eGFRCreatStanzick | 1.20E-05 | 2.16E-06 | TRUE | 0.323309054 | HbA1c |
| Vyjxei | bhH1Up | AMPK | eGFRCreatStanzick | 1.27E-04 | 4.31E-06 | TRUE | 2.00E-06 | HbA1c |
| WbqWq0 | bhH1Up | GDF15 | eGFRCreatStanzick | 3.88E-05 | 6.54E-06 | TRUE | 0.058413318 | HbA1c |
| Xjf1jJ | bhH1Up | SGLT2I | eGFRCreatStanzick | 5.19E-04 | 6.52E-06 | TRUE | 3.22E-25 | HbA1c |
| 9rRHy9 | YFZ69l | PCSK9I | eGFREurStanzick | 0.005963165 | 5.30E-05 | TRUE | 0 | LDL |
| D94SpL | YFZ69l | ACEI | eGFREurStanzick | 1.14E-04 | 2.82E-06 | TRUE | 6.83E-09 | SBP |
| f3yEFg | YFZ69l | CCB | eGFREurStanzick | 0.002247725 | 3.07E-05 | TRUE | 1.68E-157 | SBP |
| fhrnCj | YFZ69l | MG3 | eGFREurStanzick | 3.89E-05 | 1.67E-07 | TRUE | 0.003352414 | HbA1c |
| Fq9tX7 | YFZ69l | Lowering LDL | eGFREurStanzick | 0.042030245 | 0.001877254 | TRUE | 0 | LDL |
| fwCnU5 | YFZ69l | Statin | eGFREurStanzick | 0.00301332 | 2.27E-05 | TRUE | 3.92E-247 | LDL |
| IaocOm | YFZ69l | MC1 | eGFREurStanzick | 0.002199959 | 1.50E-04 | TRUE | 8.15E-68 | HbA1c |
| IBfVDL | YFZ69l | BB | eGFREurStanzick | 3.65E-04 | 1.29E-05 | TRUE | 6.87E-23 | SBP |
| JWMMYI | YFZ69l | Lowering HbA1c | eGFREurStanzick | 0.099197532 | 0.002208108 | TRUE | 0 | HbA1c |
| puxRXK | YFZ69l | GLP1RA | eGFREurStanzick | 2.68E-04 | 6.75E-07 | TRUE | 4.69E-15 | HbA1c |
| rXdiei | YFZ69l | Lowering SBP | eGFREurStanzick | 0.047501093 | 0.005636153 | TRUE | 0 | SBP |
| Sr1HqQ | YFZ69l | NPC1L1I | eGFREurStanzick | 0.001086372 | 2.11E-05 | TRUE | 1.05E-80 | LDL |
| vxLdAw | YFZ69l | GCG | eGFREurStanzick | 1.20E-05 | 2.44E-06 | TRUE | 0.347994789 | HbA1c |
| Vyjxei | YFZ69l | AMPK | eGFREurStanzick | 1.27E-04 | 6.42E-06 | TRUE | 9.93E-06 | HbA1c |
| WbqWq0 | YFZ69l | GDF15 | eGFREurStanzick | 3.88E-05 | 7.74E-06 | TRUE | 0.081972108 | HbA1c |
| Xjf1jJ | YFZ69l | SGLT2I | eGFREurStanzick | 5.19E-04 | 9.84E-06 | TRUE | 5.53E-23 | HbA1c |
| 9rRHy9 | 3lhKLG | PCSK9I | eGFRCysStanzick | 0.005963165 | 2.12E-05 | TRUE | 0 | LDL |
| D94SpL | 3lhKLG | ACEI | eGFRCysStanzick | 1.14E-04 | 7.29E-06 | TRUE | 2.21E-05 | SBP |
| f3yEFg | 3lhKLG | CCB | eGFRCysStanzick | 0.002247725 | 6.17E-05 | TRUE | 6.49E-97 | SBP |
| fhrnCj | 3lhKLG | MG3 | eGFRCysStanzick | 3.89E-05 | 4.47E-06 | TRUE | 0.067344746 | HbA1c |
| Fq9tX7 | 3lhKLG | Lowering LDL | eGFRCysStanzick | 0.040932001 | 0.003605136 | TRUE | 0 | LDL |
| fwCnU5 | 3lhKLG | Statin | eGFRCysStanzick | 0.00301332 | 2.37E-05 | TRUE | 1.83E-164 | LDL |
| IaocOm | 3lhKLG | MC1 | eGFRCysStanzick | 0.002165219 | 2.20E-04 | TRUE | 5.15E-45 | HbA1c |
| IBfVDL | 3lhKLG | BB | eGFRCysStanzick | 3.65E-04 | 2.10E-05 | TRUE | 1.50E-14 | SBP |
| JWMMYI | 3lhKLG | Lowering HbA1c | eGFRCysStanzick | 0.096794725 | 0.004016463 | TRUE | 0 | HbA1c |
| puxRXK | 3lhKLG | GLP1RA | eGFRCysStanzick | 2.68E-04 | 7.96E-06 | TRUE | 1.84E-09 | HbA1c |
| rXdiei | 3lhKLG | Lowering SBP | eGFRCysStanzick | 0.047071007 | 0.007743151 | TRUE | 0 | SBP |
| Sr1HqQ | 3lhKLG | NPC1L1I | eGFRCysStanzick | 0.001086372 | 2.05E-06 | TRUE | 6.47E-66 | LDL |
| vxLdAw | 3lhKLG | GCG | eGFRCysStanzick | 1.20E-05 | 1.29E-06 | TRUE | 0.302773622 | HbA1c |
| Vyjxei | 3lhKLG | AMPK | eGFRCysStanzick | 1.27E-04 | 8.26E-06 | TRUE | 1.90E-04 | HbA1c |
| WbqWq0 | 3lhKLG | GDF15 | eGFRCysStanzick | 3.88E-05 | 6.37E-06 | TRUE | 0.100611268 | HbA1c |
| Xjf1jJ | 3lhKLG | SGLT2I | eGFRCysStanzick | 4.38E-04 | 1.74E-06 | TRUE | 3.21E-18 | HbA1c |
| 9rRHy9 | S0yNct | PCSK9I | BUNStanzick | 0.005963165 | 2.74E-05 | TRUE | 0 | LDL |
| D94SpL | S0yNct | ACEI | BUNStanzick | 1.14E-04 | 2.16E-06 | TRUE | 1.17E-08 | SBP |

Table S7. Steiger test of directionality (continue)

| id.exposure | id.outcome | exposure | outcome | snp_r2.exposure | snp_r2.outcome | correct_causal_direction | steiger_pval | IntermediatePheno |
|---|---|---|---|---|---|---|---|---|
| f3yEFg | S0yNct | CCB | BUNStanzick | 0.002247725 | 2.05E-05 | TRUE | 6.15E-148 | SBP |
| fhrnCj | S0yNct | MG3 | BUNStanzick | 3.89E-05 | 5.15E-06 | TRUE | 0.051290459 | HbA1c |
| Fq9tX7 | S0yNct | Lowering LDL | BUNStanzick | 0.04034702 | 0.001552472 | TRUE | 0 | LDL |
| fwCnU5 | S0yNct | Statin | BUNStanzick | 0.00301332 | 6.33E-06 | TRUE | 2.40E-252 | LDL |
| IaocOm | S0yNct | MC1 | BUNStanzick | 0.002165219 | 4.42E-05 | TRUE | 5.26E-84 | HbA1c |
| IBfVDL | S0yNct | BB | BUNStanzick | 3.65E-04 | 9.26E-06 | TRUE | 3.16E-22 | SBP |
| JWMMYI | S0yNct | Lowering HbA1c | BUNStanzick | 0.097237543 | 0.001783135 | TRUE | 0 | HbA1c |
| puxRXK | S0yNct | GLP1RA | BUNStanzick | 2.68E-04 | 2.48E-05 | TRUE | 2.57E-08 | HbA1c |
| rXdiei | S0yNct | Lowering SBP | BUNStanzick | 0.047501093 | 0.003792423 | TRUE | 0 | SBP |
| Sr1HqQ | S0yNct | NPC1L1I | BUNStanzick | 0.001086372 | 5.82E-06 | TRUE | 2.65E-85 | LDL |
| vxLdAw | S0yNct | GCG | BUNStanzick | 1.20E-05 | 2.07E-07 | TRUE | 0.15831582 | HbA1c |
| Vyjxei | S0yNct | AMPK | BUNStanzick | 1.27E-04 | 1.59E-05 | TRUE | 3.17E-04 | HbA1c |
| WbqWq0 | S0yNct | GDF15 | BUNStanzick | 3.88E-05 | 8.69E-09 | TRUE | 0.00244539 | HbA1c |
| Xjf1jJ | S0yNct | SGLT2I | BUNStanzick | 5.19E-04 | 6.97E-06 | TRUE | 6.82E-23 | HbA1c |
